# Mapping Global Dietary Risk Factors for Cancer: A Structured Network-Based Analysis

**DOI:** 10.1101/2025.10.11.25337797

**Authors:** Hongyue Ma

## Abstract

Understanding the complex relationship between diet and cancer remains a critical challenge in global health research. Traditional studies have often examined isolated dietary components or focused on region-specific dietary patterns, limiting the ability to identify broader, population-level associations. In this study, a comprehensive global-scale analysis was conducted to investigate the associations between 16 dietary components and the incidence of total and subtype-specific cancers across 155 countries or regions. By integrating harmonized dietary exposure data and epidemiological records, and applying a network-based analytical framework, the study reveals both established and underexplored associations, such as the positive relationships between red meat and alcohol intake with overall cancer risk, and the inverse correlations between pulses and multiple cancer subtypes. Beyond individual associations, this study identifies interdependent dietary interaction patterns, highlighting synergistic and antagonistic effects that are often masked in conventional univariate models. The work also exemplifies how macro association (MA) networks can be constructed and dissected at a small scale to extract interpretable modules, offering a methodological foundation for broader, multi-domain MA network development. The resulting findings provide a translational basis for targeted dietary guidance and illustrate the potential of network-based approaches in advancing nutritional oncology research.

**Graphical abstract:** 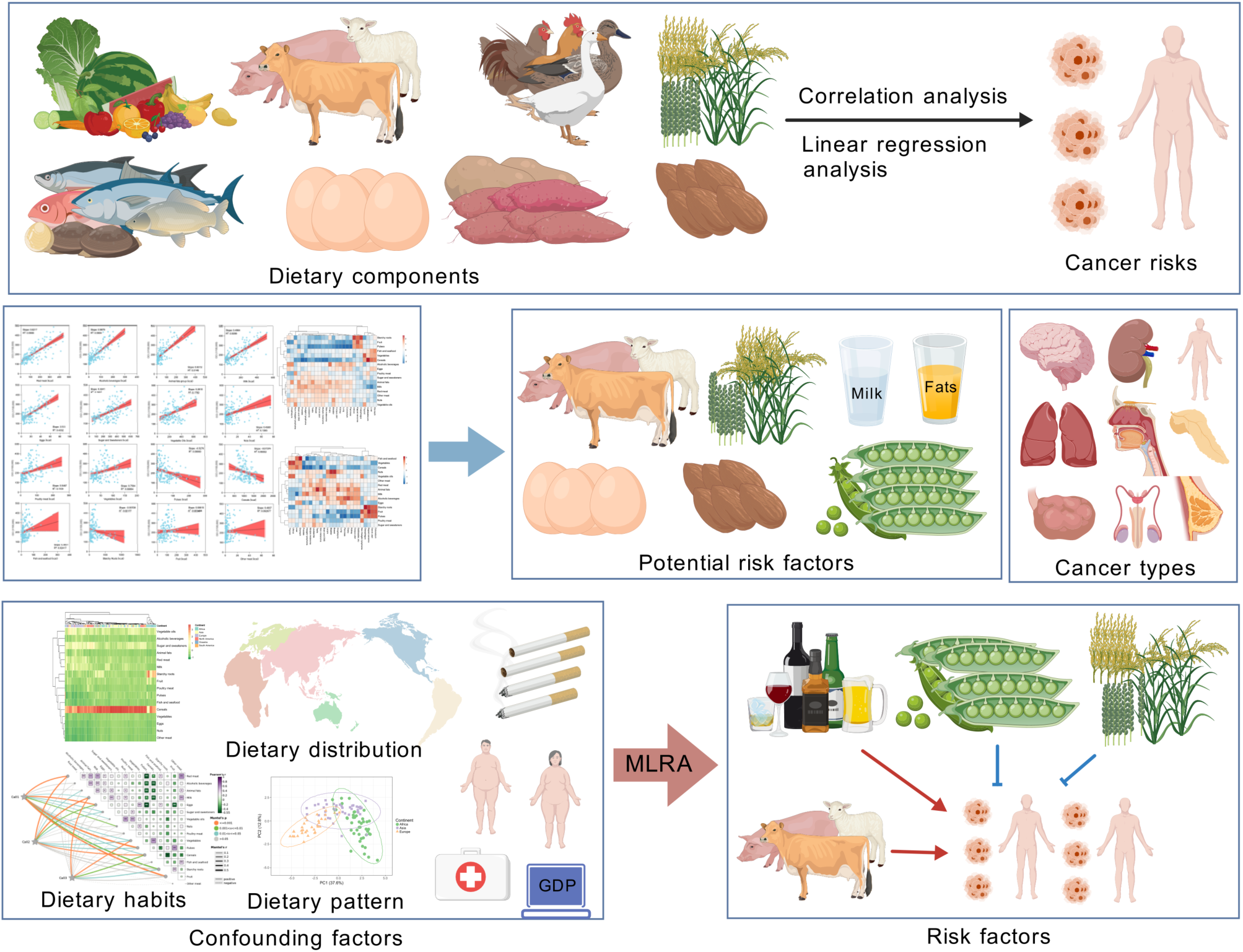

## 1. Introduction

Cancer remains one of the leading global causes of morbidity and mortality, posing a substantial threat to human health^1^. Its development is influenced by a complex interplay of genetic, environmental, and lifestyle factors, among which diet plays a particularly pivotal role^2,3^. As one of the most intimate and modifiable exposures in daily life, dietary intake exerts profound impacts on health, yet its relationship with cancer risk remains controversial and poorly resolved^4,5^.

Historically, epidemiological studies have predominantly focused on isolated dietary components to explore their associations with cancer. Alcohol consumption, red meat intake, and fruit and vegetable consumption have been separately linked to various cancers^6–9^. For instance, red meat has been associated with colorectal cancer, while high fruit and vegetable intake has often been reported as protective^8,9^. However, these findings have been inconsistent. Some large-scale studies have failed to establish a clear link between alcohol or red meat consumption and increased cancer incidence, suggesting potential variability across populations, methodological limitations, or underlying confounders^10–12^.

A major limitation of single-nutrient or single-food studies lies in their inability to account for dietary complexity. In real-world settings, individuals consume diverse food components simultaneously, which may interact synergistically or antagonistically^6,13^. The reductionist approach, by examining one food item in isolation, overlooks these dietary interdependencies and thus may misrepresent true risk estimates^13,14^. There is a growing need to shift toward analyzing dietary exposures as composite dietary groups, or dietary patterns, to better reflect the biological reality of diet-cancer interactions.

In response, an increasing number of studies have examined predefined dietary patterns such as the Mediterranean diet and the Western dietary pattern to evaluate their collective influence on cancer. The Mediterranean diet, characterized by high intake of vegetables, fruits, whole grains, legumes, fish, and olive oil, has been associated with a reduced cancer risk^15,16^. In contrast, the Western dietary pattern, rich in red and processed meats, refined grains, sugar-sweetened beverages, and high-fat dairy products, has been consistently linked to elevated cancer incidence^17–19^. While such pattern-based approaches capture the overall effect of dietary profiles, they often obscure the individual contribution of specific food components^20,21^. This limitation hinders the ability to formulate targeted dietary recommendations and complicates public understanding of how specific dietary modifications may reduce cancer risk.

Moreover, most existing studies have been confined to specific populations and geographic regions, often relying on self-reported food frequency questionnaires (FFQs), which may introduce recall and selection biases^5,22,23^. Such regional constraints limit the generalizability of findings and fail to reflect the diversity of global dietary habits. Furthermore, many studies suffer from relatively small sample sizes, homogeneous cohorts, and conventional analytic frameworks, limiting their capacity to uncover novel associations or capture the global heterogeneity of diet-cancer relationships^5,24,25^.

From a methodological standpoint, existing research rarely addresses the intercorrelation between dietary components or examines the structure of dietary networks. It also tends to overlook dose-response relationships and fails to integrate dietary exposures with confounding factors such as socioeconomic status, caloric intake, or metabolic indicators^5,26,27^. Consequently, despite a large body of literature, our understanding of diet-cancer links remains fragmented.

At the molecular level, mechanistic studies have elucidated some potential pathways through which diet modulates carcinogenesis. For example, red and processed meats can generate carcinogenic compounds such as heterocyclic amines (HCAs) and polycyclic aromatic hydrocarbons (PAHs) during high-temperature cooking^28^. These compounds promote DNA damage and inflammation, contributing to tumor initiation and progression^29,30^. Diets high in added sugars may activate the IGF-1 signaling pathway, enhancing cellular proliferation and inhibiting apoptosis^31–33^. Conversely, omega-3 polyunsaturated fatty acids abundant in fish, such as EPA and DHA, exert anti-inflammatory effects and may disrupt tumor cell membrane architecture^34,35^. Soy isoflavones, acting as phytoestrogens, modulate gene expression in hormone-sensitive cancers^36,37^, while dairy intake has been linked to elevated IGF-1 levels and potential risk of prostate cancer^38,39^. However, these represent only a narrow subset of the diverse mechanistic pathways through which diet may influence cancer risk. Such findings further underscore the complexity of whole-food matrices, wherein the health effects of individual nutrients may be modulated, amplified, or attenuated by interactions within broader dietary contexts.

Taken together, there is an urgent need for globally scalable, integrative approaches to dissect the multi-layered relationships between dietary components and cancer risk. Such approaches should move beyond simplistic one-to-one associations or binary dietary pattern models, and instead adopt multi-factorial analytical frameworks capable of capturing the interactions within and between dietary groups.

This study presents a comprehensive global analysis of dietary exposures and cancer incidence across multiple countries and regions. Both individual food components and composite dietary structures are systematically evaluated, with network level insights integrated to explore cooperative and antagonistic relationships among dietary variables. The work also serves as a case example demonstrating how small-scale macro association (MA) networks can be extracted and analyzed to yield interpretable, domain specific insights^40^. By moving beyond univariate correlations toward small-scale dietary interaction networks, this study contributes a methodological foundation for advancing population-level nutritional oncology research with improved resolution and contextual relevance.

## 2. Methods

### 2.1 Data sources and selection

Data on overall cancer incidence (OCI) for 155 countries or regions spanning 1992-2017 were obtained from the Our World in Data database (https://ourworldindata.org/grapher/per-capita-meat-consumption-by-type-kilograms-per-year, accessed on 1 October 2025). Corresponding dietary composition data, which included 16 dietary components for the same countries or regions and time period, were obtained from the same source (https://ourworldindata.org/grapher/dietary-composition-by-country?country=~OWID_WRL, accessed on 1 October 2025), using standardized preprocessing protocols as applied in previous analyses^9,40^. Age-standardized incidence data for 26 cancer types between 1999 and 2010, covering 40 countries or regions, were obtained from the Global Cancer Observatory (https://gco.iarc.fr/overtime/, accessed on 1 October 2025). National-level (gross domestic product) GDP per capita data were sourced from Our World in Data (https://ourworldindata.org/grapher/gdp-per-capita-maddison-project-database, accessed on 1 October 2025), while gender ratio statistics were extracted from the World Bank database (https://data.worldbank.org/indicator/SP.POP.TOTL.MA.ZS?end=2021&start=2021&view=map&year=2021, accessed on 1 October 2025). For each variable, national-level values were averaged across the available years. Cancer subtype incidence rates for each country or regions were adjusted for gender distribution using the corresponding gender ratios. Data on the proportion of adults with a body mass index (BMI) greater than 25 (classified as overweight or obese), smoking prevalence, and daily per capita caloric supply were also obtained from Our World in Data (https://ourworldindata.org).

### 2.2 Statistical analysis

#### 2.2.1 Association between dietary components and cancer incidence

Pairwise Pearson correlation coefficients were computed between the 16 dietary components and both the OCI (155 countries or regions) and 26 individual cancer types (40 countries or regions), using SPSS (IBM SPSS Statistics). Linear regression models evaluating the association between OCI and dietary components were constructed using GraphPad Prism 9. Radar plots summarizing correlation patterns were generated using Python (v3.11). The graphical abstract was created by BioGDP (https://biogdp.com, accessed on 1 October 2025).

#### 2.2.2 The key confounding

To account for economic confounding, partial correlation analyses between cancer incidence and dietary components were conducted while controlling for GDP per capita (using SPSS). A correlation heatmap of six representative cancer types and their association with dietary components was visualized using ClustVis (https://biit.cs.ut.ee/clustvis/, accessed on 1 October 2025).

#### 2.2.3 Mapping dietary component distributions and global dietary patterns

Heatmaps representing the distribution of 16 dietary components across the 155 countries or regions (OCI dataset) and the 40 countries or regions (cancer subtype dataset) were generated using ClustVis (https://biit.cs.ut.ee/clustvis/, accessed on 1 October 2025). Corresponding dietary association networks were constructed and visualized using Chiplot (https://www.chiplot.online, accessed on 1 October 2025).

#### 2.2.4 Regional dietary patterns and their relationship with cancer

Principal component analysis (PCA) of dietary patterns among the 155 countries or regions was performed using ClustVis (https://biit.cs.ut.ee/clustvis/, accessed on 1 October 2025). Comparative analyses of dietary intake, GDP per capita, and cancer incidence across Africa, Asia, and Europe were conducted using GraphPad Prism 9. Linear regressions stratified by continent were implemented using Chiplot (https://www.chiplot.online, accessed on 1 October 2025), and choropleth maps representing geographic trends in cancer incidence and dietary exposure were generated using Tableau Desktop (v2023.2).

#### 2.2.5 Association between different dietary components

Pearson correlation matrices among the 16 dietary components were calculated for both the 155-country or regions (overall cancer) and 40-country or regions (cancer subtype) datasets. Heatmaps and Mantel regression analyses were conducted in Chiplot (https://www.chiplot.online, accessed on 1 October 2025) to assess structural coherence and pairwise distances among dietary intake profiles.

#### 2.2.6 Multiple linear regression analysis of dietary components and cancer incidence

Multivariable linear regression models were constructed to evaluate the joint effects of multiple dietary components on cancer incidence, using SPSS. In selected models, adjustments were made for potential confounders including GDP per capita, mean daily caloric intake, BMI, and smoking prevalence where data permitted. Variance inflation factors (VIFs) were calculated to assess multicollinearity.

## 3. Results

### 3.1 Association between dietary components and OCI

The consumption of 12 dietary components, including red meat (CC = 0.769, *p* < 0.001), alcoholic beverages (CC = 0.768, *p* < 0.001), pulses (CC = -0.298, *p* < 0.001), and cereals (CC = -0.289, *p* < 0.001), was found to be significantly associated with OCI (Figure 1a and Table S1). However, dietary patterns are influenced by a complex interplay of factors such as GDP, lifestyle habits, and geographic location, making it challenging to accurately identify potential cancer risk factors through simple Pearson correlation analysis^41^.

**Figure 1.**
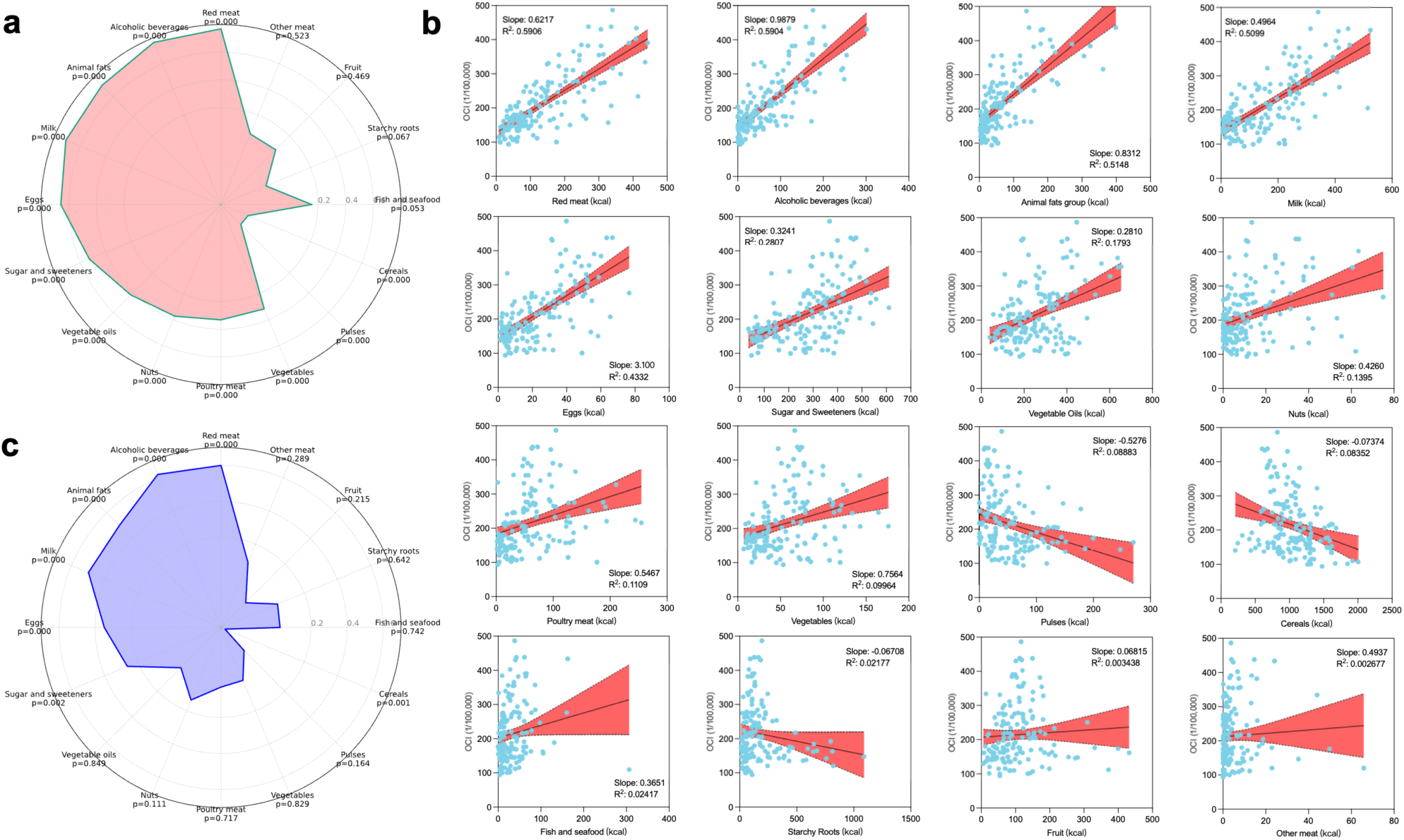
Radar and scatter plots of association between dietary components and OCI. (a) Radar plot shows Pearson correlation coefficients between the intake of 16 dietary components and OCI across 155 countries or regions. (b) Scatter plots displaying linear regression analyses of the association between dietary components and OCI. (c) Radar plot shows partial correlation coefficients between the intake of 16 dietary components and OCI after adjusting for GDP per capita, based on partial correlation coefficients.

Given that dietary habits are largely influenced by economic capacity, GDP per capita was considered a major factor affecting dietary choices as before^42,43^. Additionally, regions with higher GDP tend to have higher cancer detection rates, suggesting that GDP also impacts cancer incidence^44–46^. Pearson correlation analysis between GDP per capita and 16 dietary components, along with OCI, revealed a strong correlation between GDP per capita and OCI, as well as with 10 dietary components including red meat consumption and alcoholic beverage consumption (Figure S1 and S2). This suggests that GDP per capita may be a confounding variable in the relationship between cancer incidence and dietary components. Therefore, GDP per capita was used as a control variable in further correlation analyses. After controlling for GDP per capita, the consumption of red meat (CC = 0.600, *p* < 0.001), alcoholic beverages (CC = 0.620, *p* < 0.001), animal fats (CC = 0.500, *p* < 0.001), milk (CC=0.498, *p* < 0.001), eggs (CC = 0.349, *p* < 0.001), sugar and sweeteners (CC = 0.264, *p* < 0.002) remained positively associated with OCI, while cereal consumption (CC = -0.276, *p* < 0.001) was negatively associated with OCI (Figure 1b). However, the consumption of vegetable oils (CC = 0.016, *p* = 0.849), poultry meat (CC = 0.031, *p* = 0.717), nuts (CC = 0.136, *p* = 0.111), and pulses (CC = -0.119, *p* = 0.164) became non-significant.

Furthermore, to enhance the reliability of the results, linear regression analyses were conducted between the 16 dietary components and OCI (Figure 1c). Red meat (R² = 0.5906), alcoholic beverages (R² = 0.5904), animal fats (R² = 0.5148), milk (R² = 0.5099), and sugar and sweeteners (R² = 0.2807) all exhibited relatively high R² values, whereas cereals showed a low R² (0.0835, < 0.1). Consequently, these five factors may be potential cancer risk factors. Although cereals demonstrated a potential for reducing cancer risk, further consideration is warranted due to its low R² value.

In summary, for overall cancer, the consumption of red meat, alcoholic beverages, animal fats, milk, sugar, and sweeteners may be considered potential risk factors.

### 3.2 Association between dietary components and cancer incidences

#### 3.2.1 Association between dietary components and cancer incidences

Given the diverse etiological factors and pathogenic patterns associated with different types of cancer (Figure S3), it is imperative to investigate the relationship between cancer subtypes and various dietary components. Cluster analysis was employed to explore the similarities and differences in Pearson’s correlation coefficients between 26 types of cancer and 16 dietary components. As illustrated in Figure 2a, cancers were categorized into three clusters (the corresponding network visualization is presented in Figure S4a). The first cluster included stomach, liver, gallbladder, and thyroid cancers, which exhibited correlations with high consumption of fish and seafood, vegetables, and cereals. The second cluster, comprising esophagus cancer, Kaposi sarcoma, and uterus cancer, demonstrated associations with high intake of starchy roots, fruit, and pulses. It is important to note that these correlations do not imply causation but rather reflect geographical similarities. The third cluster encompassed 19 types of cancer, including leukemia, breast cancer, and pancreatic cancer. This large group of cancers showed correlations with high consumption of alcoholic beverages, eggs, poultry meat, sugar and sweeteners, animal fats, milk, red meat, other meats, nuts, and vegetable oils. Evidently, the relationship between this cluster of cancers and dietary components is akin to that of the OCI. These cancers may be closely related to dietary components, with the associated high-incidence dietary components potentially being high-risk factors for cancer.

**Figure 2.**
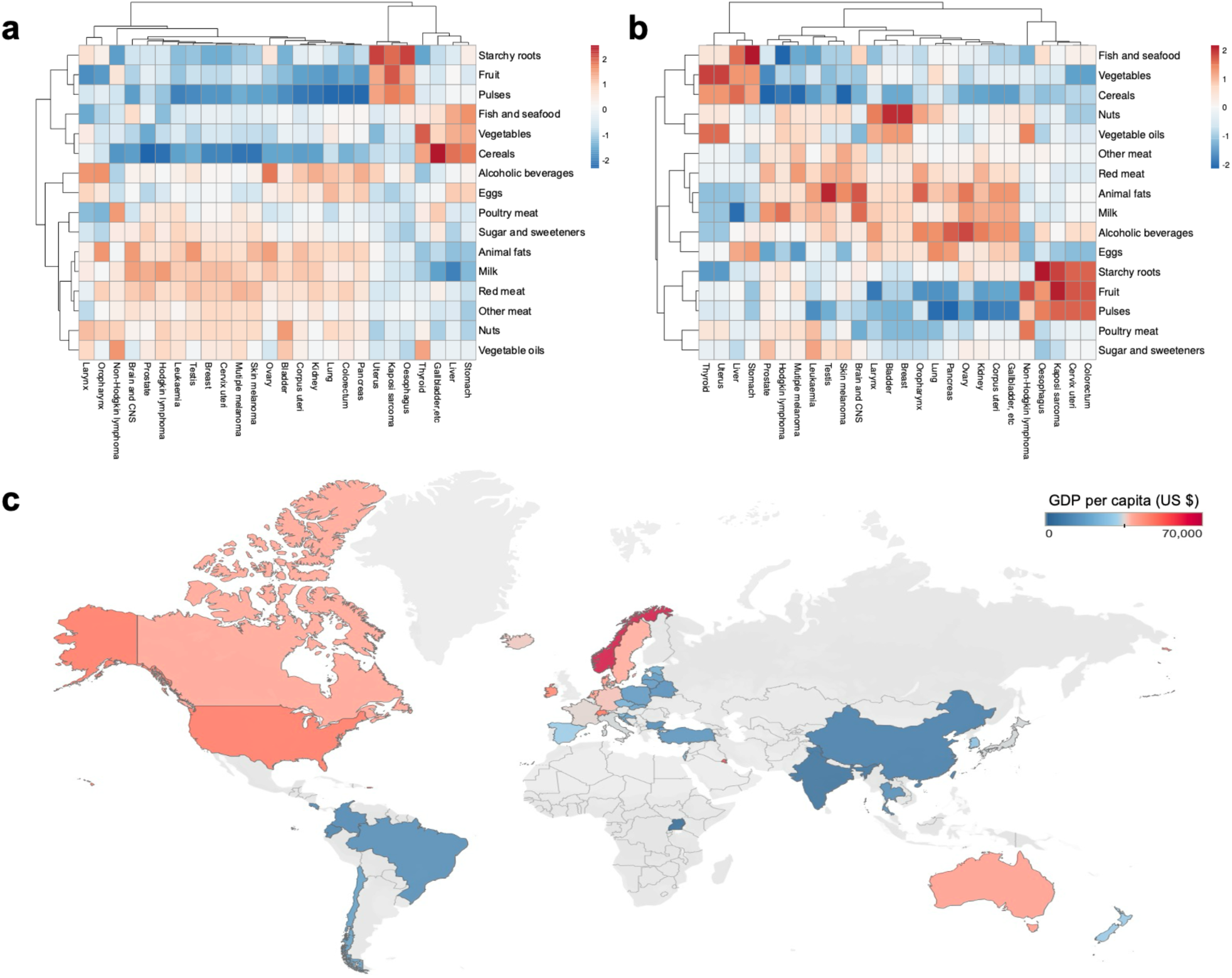
Heatmap of the relationship between dietary components and cancer incidences. (a) Heatmap shows Pearson correlation coefficients between the intake levels of 16 dietary components and the incidences of 26 cancer subtypes across 40 countries or regions (Table S5-S30). (b) Heatmap shows partial correlation coefficients between the intake levels of 16 dietary components and the incidences of 26 cancer subtypes across 40 countries or regions after adjusting for GDP per capita (Table S5-S30). (c) Geographic heatmap illustrating the spatial distribution of average GDP per capita (1999-2010) across the 40 countries or regions included in the analysis of dietary and cancer subtype associations.

#### 3.2.2 The confounding of GDP per capita

Similar to the OCI, cancer subtypes are also subject to confounding by GDP per capita. Among the 10 dietary components associated with 19 types of cancer, all but “other meat” showed significant correlations with GDP per capita (Figure S2 and Table S3). The incidence rates of 17 types of cancer were significantly correlated with GDP per capita (Figure S5 and Table S4), with all but stomach and uterus cancer falling within the cluster of cancers related to dietary components (as shown in Figure 2a among the 19 types of cancer).

Additionally, the regularity in the incidence rates of various cancer types across countries or regions warrants further exploration as a reference. In Figure 2b, the incidence rates of 26 types of cancer were clustered into two groups, with all but cervix uteri cancer from the 19 types of cancer in Figure 2a being clustered into one group (the corresponding network visualization is presented in Figure S4b). This group of cancers predominantly occurs in regions with higher GDP per capita, primarily in Europe and America, while the other group has lower incidence rates in these regions and relatively higher rates in Asia, Africa, and South America.

The strong regularity observed reflects the global distribution pattern of cancer incidence rates but also serves as a reminder that differences in GDP per capita could potentially confound the correlations between cancer and dietary components.

In Figure 2c, partial correlation coefficients between cancer incidences and dietary components, controlling for GDP per capita, were clustered. Cancers were still clustered into three groups, with pancreas cancer, leukemia, breast cancer, and others forming a major group. Compared to Figure 2a, oropharynx cancer was included in this group, while non-Hodgkin lymphoma, colorectal cancer, cervix uteri cancer, Kaposi sarcoma, and esophagus cancer formed another group. Regarding dietary components, the two clusters from Figure 2a changed to three, with red meat, animal fats, milk, alcoholic beverages, eggs, and other meat remaining in the same cluster, associated with the high incidence of the major group of cancers such as pancreas cancer, leukemia, and breast cancer. The relationship between cancer and dietary components undergoes some changes after controlling for GDP per capita, but the regularity remains similar. Therefore, GDP per capita may introduce some confusion into the correlation coefficients between cancer and dietary components, but after eliminating this confounding factor, strong regularity still persists.

#### 3.2.3 Potential dietary risk factors

After controlling for GDP per capita, dietary components that remained associated with the 15 types of cancer from Figure 2c were considered potentially related to cancer. Additionally, linear regression analyses between all 15 types of cancer and each dietary component were conducted as a reference standard (Figure S6-S31 and Table S5-S30), with a low R² (< 0.1) being interpreted as a lack of correlation. In Figure 4c, the relationships between the 15 potentially diet-related cancers and 16 dietary components are presented in a binary format, where dietary components associated with four or more cancer subtypes are considered potential high-risk dietary components. Consequently, alcoholic beverages, animal fats, red meat, milk, and nuts may be considered potential high-risk dietary components, while pulses and cereals may be regarded as low-risk dietary components for cancer.

### 3.3 Dietary components distribution and global dietary patterns

#### 3.3.1 Distribution of potential high cancer risk dietary components and global dietary patterns

Nearly all dietary components associated with high cancer risk demonstrate relatively elevated consumption levels in Europe, North America, Australia, and Russia, whereas their intake is markedly lower in Asia and Africa. For red meat, vegetable oils, and sugar and sweeteners, higher consumption levels are also noted in certain South American countries or regions. Additionally, red meat consumption is particularly high in China and Mongolia.

In Figure 3a, red meat, milk, animal fats, sugar and sweeteners, alcoholic beverages, and vegetable oils consistently cluster together, with the highest consumption observed in Europe and parts of North America, moderate consumption in Asia, and the lowest consumption in Africa. In contrast, the other ten dietary components form a separate cluster, exhibiting an inverse distribution pattern: highest in Africa, moderate in Asia, and lowest in Europe and parts of North America. Globally, the dietary patterns of 155 countries or regions can be broadly categorized into two contrasting clusters. One cluster, represented predominantly by Europe and North America, is characterized by higher consumption of red meat, alcoholic beverages, sugars and sweeteners, and animal and vegetable oils, coupled with lower intake of fruits, vegetables, pulses, and starchy roots. The opposite pattern, marked by lower intake of red meat, alcoholic beverages, sugars and sweeteners, and animal and vegetable oils but higher consumption of plant-based foods, is prevalent across Asia, Africa, and other developing regions. Notably, in the dataset encompassing 26 cancer subtypes from 40 countries or regions, a similar dichotomous trend in dietary structure was observed, reinforcing the global consistency of these dietary patterns. These results are highly consistent with the findings from the exploration of the relationship between dietary components and GDP per capita presented in section 3.2.2. They underscore the distinct patterns of dietary habits and preferences shaped by the substantial differences or similarities in GDP per capita across different countries or regions. Furthermore, the strong regularity observed suggests that the robust correlations between these high cancer risk factors may lead to confounding in the results.

**Figure 3.**
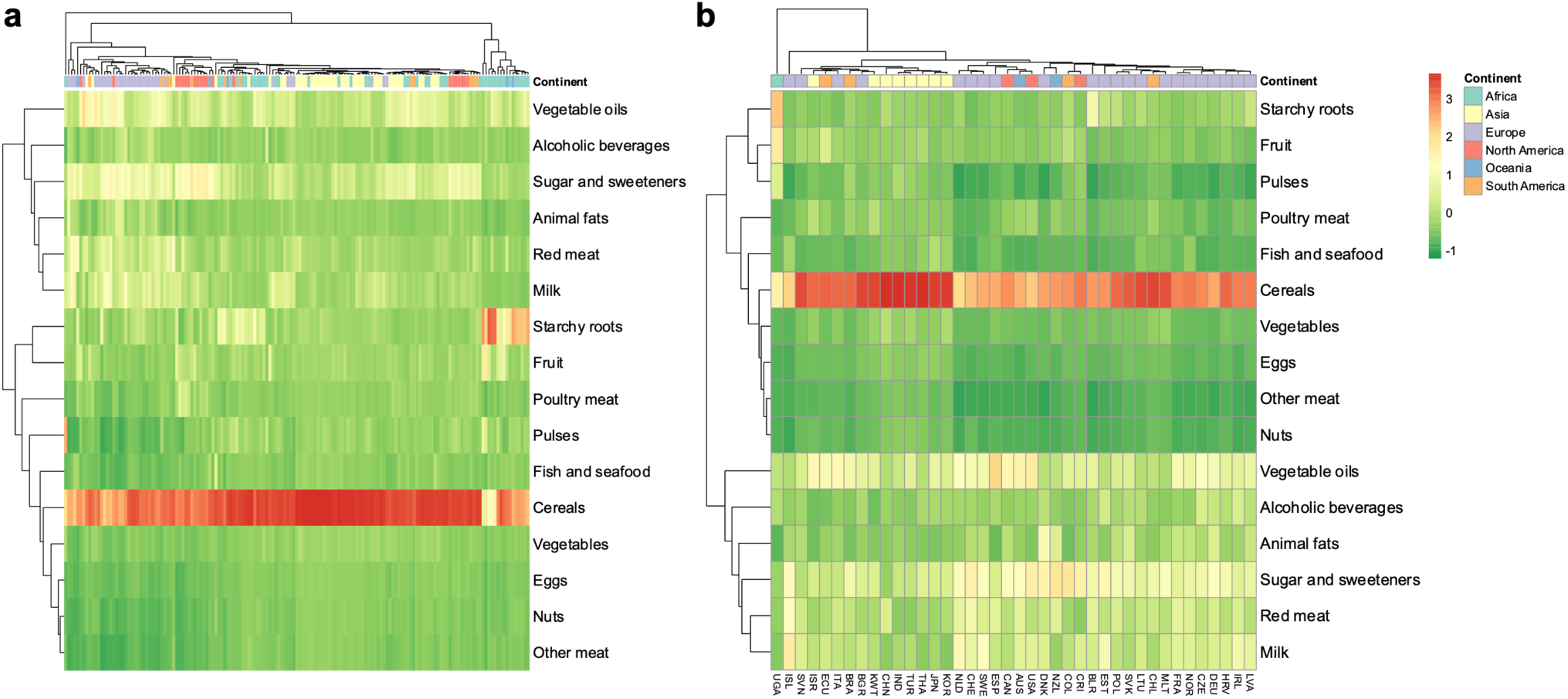
Heatmaps of dietary components. (a) Heatmap depicting the relationships among dietary components across 155 countries or regions included in the OCI dataset. (b) Heatmap depicting the relationships among dietary components across 40 countries or regions included in the dataset of 26 cancer subtypes.

#### 3.3.2 Regional variations in dietary patterns and their associations with cancer incidence

Figures 3b and 3c demonstrate a pronounced clustering of dietary profiles at the continental level, prompting a PCA to further characterize regional dietary structures and evaluate whether the observed associations with OCI remain consistent across distinct dietary contexts. Globally, dietary patterns exhibit considerable overlap (Figure 4a); however, exclusion of South America, North America, and Oceania reveals that Asian and African countries or regions possess more distinct dietary compositions, whereas European countries or regions appear partially aligned with both Asian and African clusters (Figure 4b).

**Figure 4.**
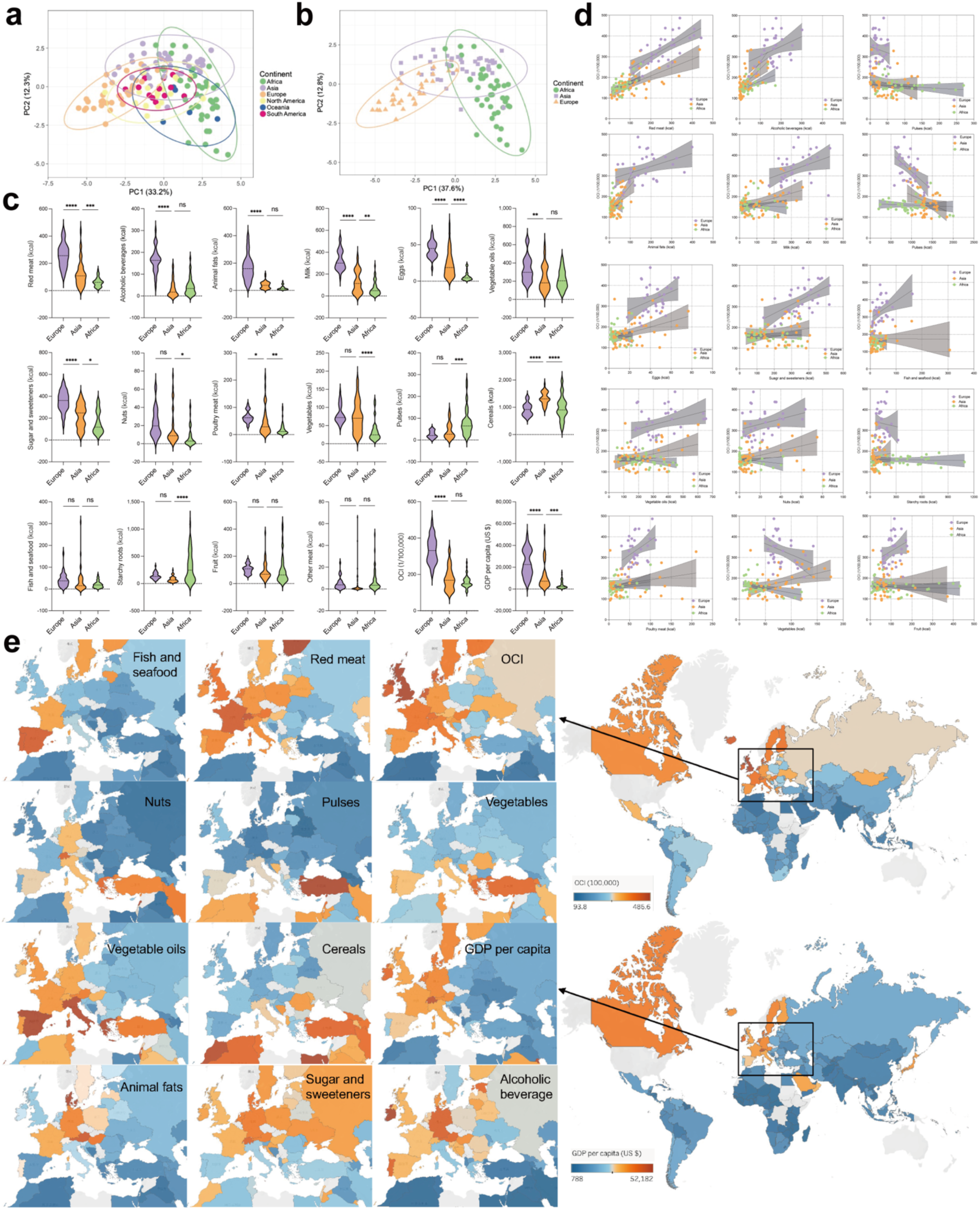
Regional variation in dietary components and their associations with OCI. (a) PCA of 16 dietary components across 155 countries or regions. (b) PCA of the same 16 dietary components within Asia, Africa, and Europe. (c) Comparative distribution of the 16 dietary components, OCI, and GDP per capita across Asia, Africa, and Europe. (d) Region-specific associations between the 16 dietary components and OCI in Asia, Africa, and Europe. (e) Distribution of 10 dietary components, OCI, and GDP per capita in Mediterranean countries or regions.

Comparative analysis of the 16 dietary components, GDP per capita, and OCI across Asia, Africa, and Europe (Figure 4c) reveals that European countries or regions have significantly higher cancer incidence rates and economic development levels than the other two regions. These elevated OCI levels coincide with greater consumption of dietary components associated with cancer risk, including red meat, alcoholic beverages, and sugar and sweeteners, etc. These factors follow consistent regional gradients and align with previously identified risk profiles.

Importantly, the associations between dietary variables and OCI observed within each continent mirror the trends identified at the global level (Figure 4d), suggesting the presence of robust and geographically consistent diet-cancer relationships.

The Mediterranean region exhibits a particularly distinct dietary signature, characterized by increased intake of fish, legumes, and vegetables, and lower levels of red meat, animal fats, and alcohol. As illustrated in Figure 4e, this regional pattern contrasts sharply with the higher-risk dietary profiles of Northern Europe. Notably, countries or regions within the Mediterranean basin also report markedly lower OCI, reinforcing the potential protective role of this dietary structure. The comparatively lower GDP per capita in Mediterranean countries or regions further suggests that differences in dietary patterns arise not only from cultural and geographical determinants but also from underlying economic structures that may exert a substantial influence on dietary choices and nutritional health outcomes. These findings underscore the relevance of continental and subregional dietary patterns in modulating cancer risk. Furthermore, they highlight the importance of spatially resolved analyses in disentangling global versus local nutritional influences on cancer epidemiology.

#### 3.3.3 Association between different dietary components

Quantitatively discerning the associations among various dietary component consumptions significantly aids in elucidating dietary patterns and crafting enhanced dietary strategies. As delineated in section 3.3.1, the high correlation among different dietary components can obfuscate the precision of outcomes, particularly the interconnection of dietary elements associated with elevated cancer risks may exhibit correlations with variables ostensibly unlinked to cancer risk. However, in section 3.3.1, a pronounced uniformity in the distribution of red meat, milk, animal fats, sugar and sweeteners, alcoholic beverages, and vegetable oils is also observed, thus prompting a quantitative analysis of the relationships among 16 dietary component consumptions (Figure 5a and 5b).

**Figure 5.**
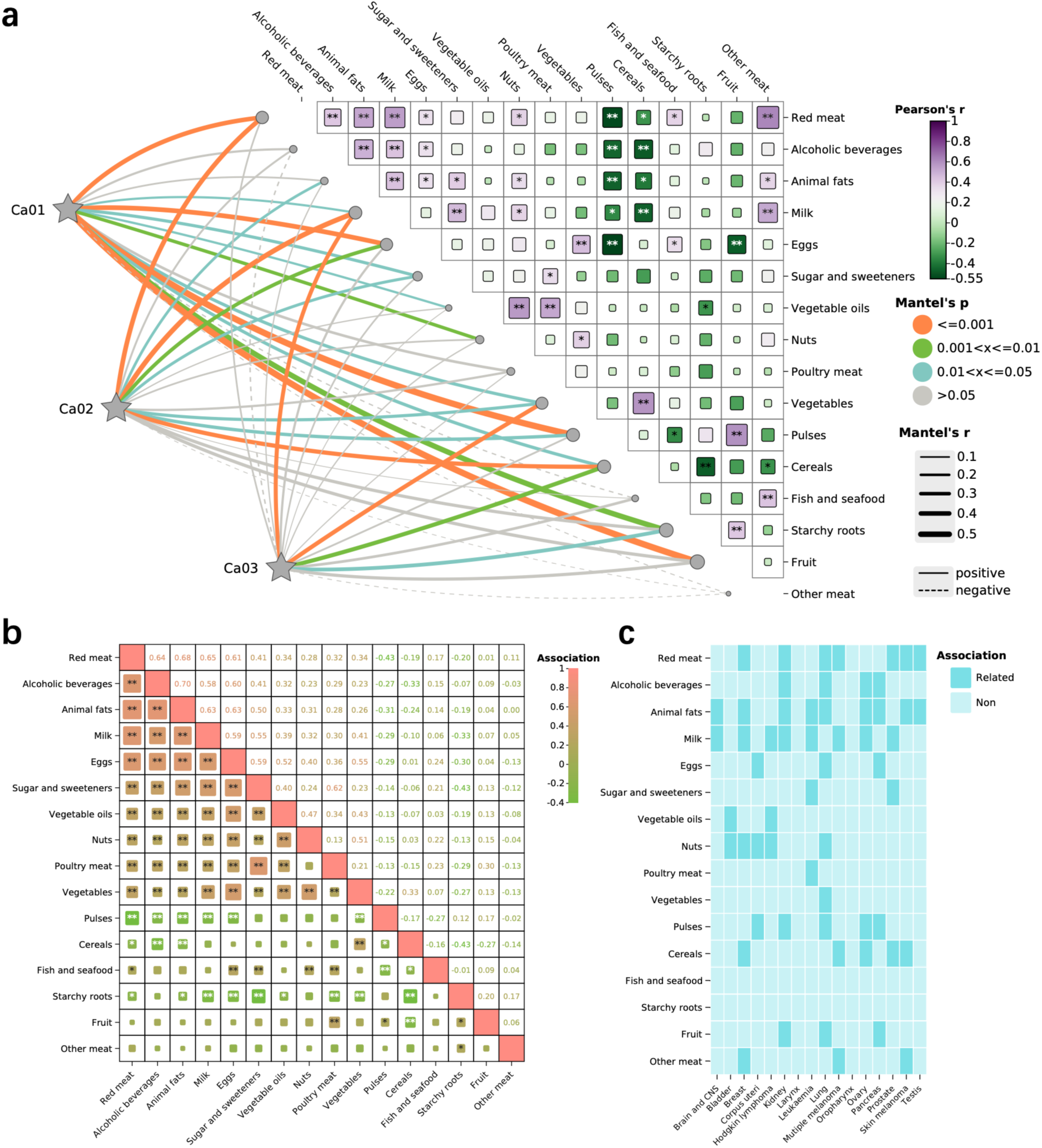
Heatmaps depicting interrelationships among dietary components. (a) Associations among 16 dietary components across 155 countries or regions in the dataset for OCI. (b) Corresponding associations among the same 16 dietary components across 40 countries or regions in the dataset for 26 cancer subtypes. (c) Binary map summarizing the presence or absence of statistically significant associations between the 16 dietary components and cancer incidence. The significance of each association was determined based on the criteria established in Section 3.2.3, whereby relationships that remained significant after adjustment for GDP per capita and demonstrated a regression coefficient of determination (R^2^) greater than 0.1 were considered robustly associated.

In Figure 5b, the relational matrix of 16 dietary components across 155 nations is showcased, revealing a robust correlation among them. The consumptions of red meat, alcoholic beverages, animal fats, milk, eggs, sugar and sweeteners, vegetable oils, nuts, poultry meat, and vegetables are significantly positively correlated, with red meat, alcoholic beverages, animal fats, milk, and eggs nearly exhibiting a highly positive correlation (CC > 0.6). Pulses display a notable negative correlation with the consumption of red meat, alcoholic beverages, animal fats, milk, eggs, and vegetables, while cereals show a significant negative correlation with red meat, milk, and pulses but a positive correlation with vegetables. Starchy roots almost universally demonstrate a significant negative correlation with the consumption of all dietary components, whereas fish and seafood, fruit, and other meats do not exhibit discernible regularity in their correlations with other dietary components.

In Figure 5a, the relational matrix of 16 dietary components across 40 nations is displayed, albeit with a weaker overall correlation compared to Figure 5b. The five dietary components, red meat, alcoholic beverages, animal fats, milk, and eggs, that showed a high positive correlation in Figure 5b remain significantly correlated, while pulses and cereals consumption exhibit a notable negative correlation with these four dietary components. Furthermore, the Mantel regression analysis of three cancer types categorized into three groups with 16 dietary components, as shown in Figure 2b, failed to reveal patterns akin to those significant in Section 3.2 (“Ca01” includes 5 types of cancer, “Ca02” includes 17 types of cancer, “Ca03” includes 4 types of cancer).

### 3.4 Multiple linear regression analysis of dietary components and cancer incidence

To minimize confounding effects, including high correlations between different dietary components that confound the accuracy of the results as mentioned in Section 3.3.3, multivariable linear regression models were employed to more precisely assess the association between cancer-related dietary components between cancer incidences. For overall cancer, six high-risk dietary components were selected, namely red meat, alcoholic beverages, animal fats, milk, eggs, and sugar and sweeteners, as described in Section 3.1, while controlling for GDP per capita, BMI, smoking prevalence, and total calories as confounding factors (Table 1 and S31-S33).

**Table 1.**
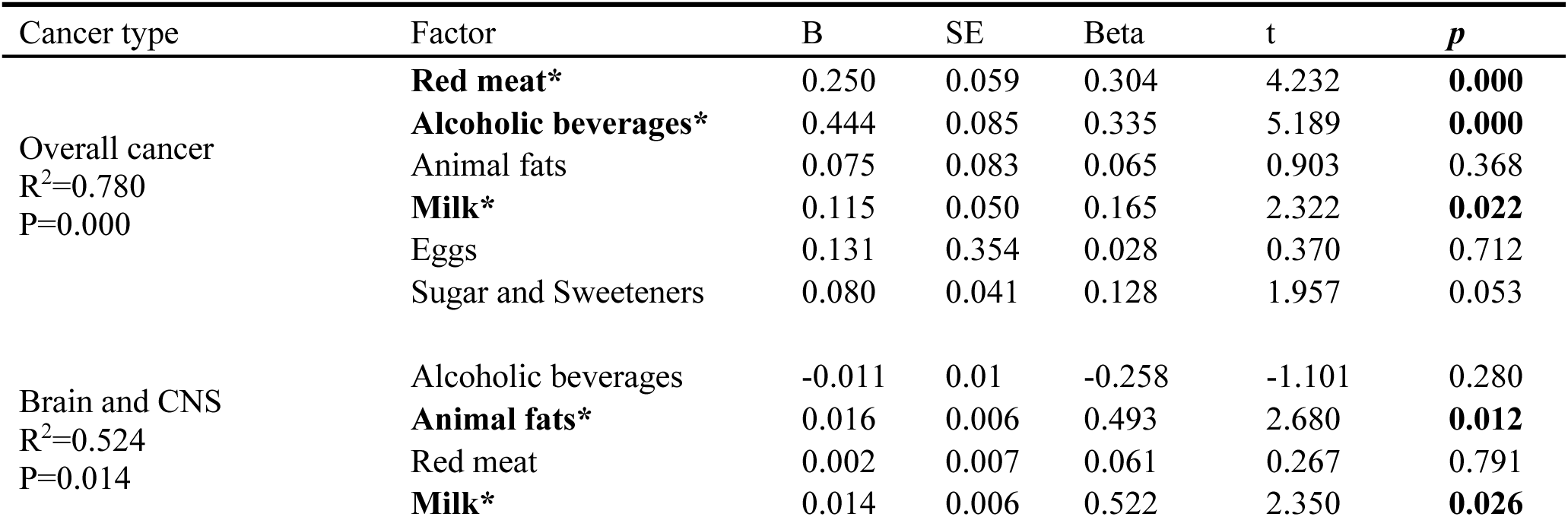
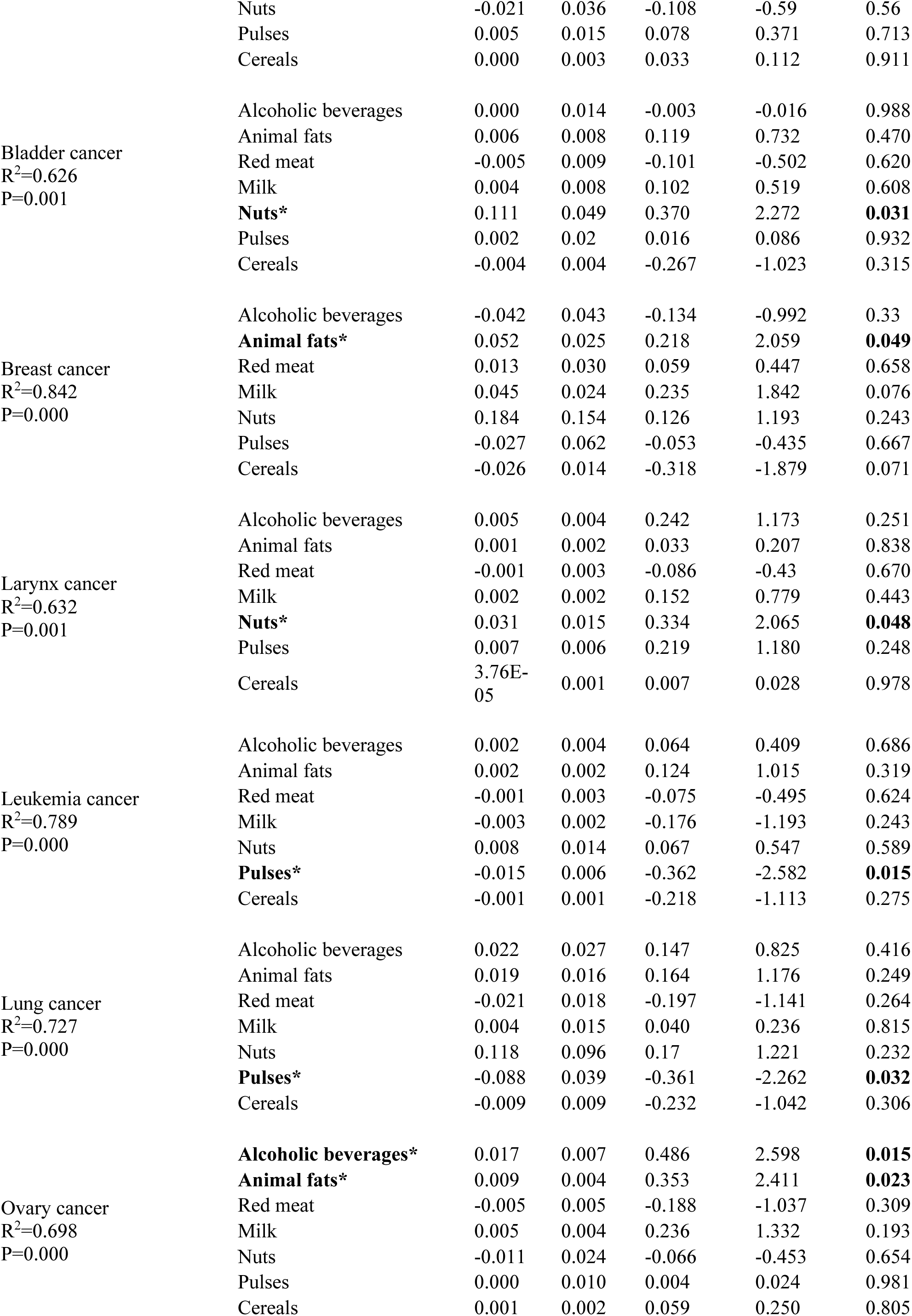
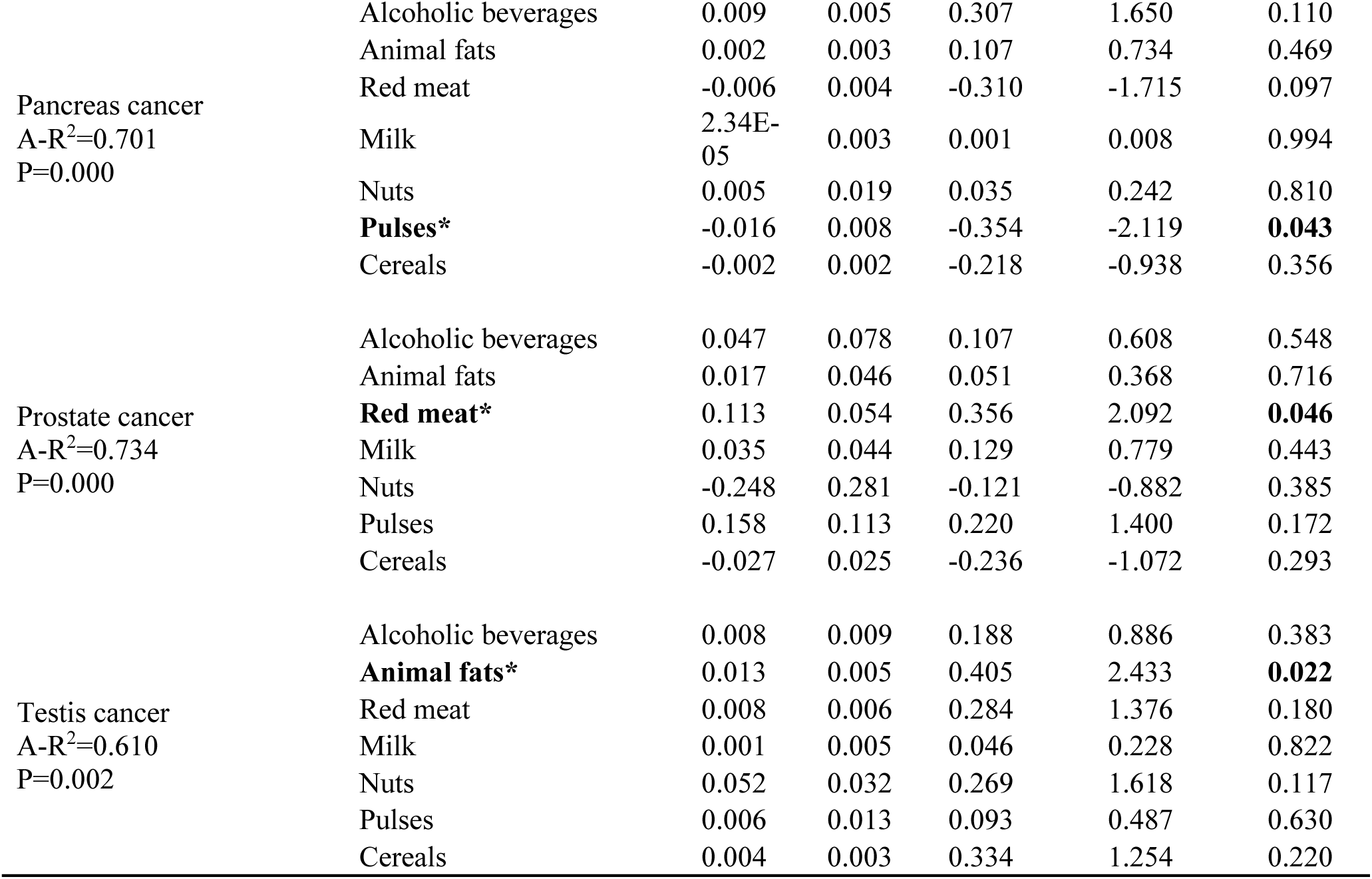
Multiple linear regression analysis of dietary components and cancer incidence.

The model exhibited good overall significance (F-statistic = 45.444, *p* < 0.001), explaining 76.3% of the variance in the data (R² = 0.780). Consumption of red meat (SE = 0.059, t = 4.232, *p* < 0.001), alcoholic beverages (SE = 0.085, t = 5.189, *p* < 0.001), and milk (SE = 0.050, t = 2.322, *p* < 0.05) were found to be significantly positively associated with overall cancer risk, with sugar and sweeteners approaching significant positive association (SE = 0.080, t = 1.957, *p* = 0.053). Animal fats (*p* = 0.368) and eggs (*p* = 0.712) were found to be non-significant. Additionally, the variance inflation factor (VIF) was less than 5, indicating no multicollinearity issues.

For cancer subtypes, five high-risk dietary components described in Section 3.2.3 (red meat, alcoholic beverages, animal fats, milk, nuts) and two low-risk dietary components (cereals and pulses) were selected as described in Section 3.2.3 (Figure 5c). Multivariable linear regression models were established between 7 dietary components and 17 diet-related cancers (as described in Section 3.2.3), also controlling for GDP per capita, BMI, smoking prevalence, and total calories as confounding factors (Table 1 and S34-S81).

Ten cancers exhibited significant associations with dietary components, with all ten models showing an R² greater than 0.5 and being significant (*p* < 0.05). Moreover, the VIF for all variables in all models was less than 10, indicating no multicollinearity issues.

Alcoholic beverages were found to be significantly positively associated with ovary cancer (SE = 0.007, t = 2.598, *p* = 0.015), and red meat with prostate cancer (SE = 0.054, t = 2.092, *p* = 0.046). Animal fats were significantly positively associated with brain and CNS (SE = 0.006, t = 2.680, *p* = 0.012), breast (SE = 0.025, t = 2.059, *p* = 0.049), ovary (SE = 0.004, t = 2.411, *p* = 0.023), and testis cancer (SE = 0.005, t = 2.433, *p* = 0.022). Milk was significantly positively associated with brain and CNS cancer (SE = 0.006, t = 2.350, *p* = 0.026), and nuts with bladder (SE = 0.049, t = 2.272, *p* = 0.031) and larynx cancer (SE = 0.015, t = 2.065, *p* = 0.048). Pulses were found to be significantly negatively associated with leukemia (SE = 0.006, t = -2.582, *p*= 0.015), lung (SE = 0.039, t = -2.262, *p* = 0.032), and pancreas cancer (SE = 0.008, t = -2.119, *p* = 0.043). However, no significant association was found between cereals and any cancer type.

## 4. Discussion

This study presents a global-scale, quantitative investigation of the associations between dietary components and cancer incidence across multiple countries or regions and cancer subtypes. By integrating harmonized, population-level dietary and epidemiological data, the analysis demonstrates that cancer risk is not governed by single dietary components in isolation, but rather by complex, interdependent dietary patterns that vary across regions and cancer types.

This analysis reveals several robust and biologically plausible associations, such as the positive correlation between red meat or alcoholic beverage consumption and overall cancer incidence, that are consistent with prior epidemiological and mechanistic studies^7,47–49^. However, the network-based approach also uncovers non-trivial, less-discussed relationships (for example, the inverse association between pulse intake and several cancer subtypes), including potential antagonistic effects between certain food groups and synergistic effects among others. These findings suggest that conventional one-to-one statistical frameworks may underestimate or mischaracterize dietary effects when ignoring the broader dietary context. Moreover, these results provide a translational foundation for more targeted dietary guidance. While broad dietary patterns remain useful for public communication, the identification of key dietary components and their interaction signatures may inform more personalized or population-specific prevention strategies.

Importantly, this study serves as a conceptual microcosm of the broader MA network framework. By progressing from conventional pairwise associations to one-to-many and many-to-many relationships between dietary factors and cancer subtypes, the analysis demonstrates a foundational strategy for building MA networks across domains^40^. Both one-star and two-star dietary-cancer association subnetworks were constructed, and further efforts were made to analyze intra-network structure, uncover hidden interactions, and identify potential confounding pathways. These subnetworks, although limited in scale, exemplify how interpretable modules can be extracted from high-dimensional global datasets, representing a methodological advance toward the construction of comprehensive, domain-wide MA networks.

Nevertheless, several limitations must be acknowledged. First, although the Three Stars Global Macro Association-analysis (TSGMA) framework was applied to construct one-star and two-star dietary-cancer association networks, higher-order (three-star) temporal associations between dietary patterns and cancer incidence remain unexplored. Second, the magnitude of association strength at each MA level has not yet been quantitatively linked to real-world dose-response effects, limiting interpretability for clinical or policy translation. Third, despite constructing multi-factorial networks, the number of variables remains orders of magnitude below what is necessary for full-scale MA networks, rendering this study a conceptual demonstration rather than a comprehensive implementation.

Future research should expand this framework by incorporating a broader range of variables, temporal dynamics, and systematic TSGMA analyses across domains^9,40^. Prospective or interventional designs will be essential for validating observed associations^50,51^. Moreover, within the dietary-cancer network domain, integrating molecular and omics-level mediators, such as metabolomics or microbiome-derived pathways, may yield deeper mechanistic insights and enhance translational relevance^52–56^.

## 5. Conclusion

This study offers not only empirical insights into the associations between dietary exposures and cancer incidence across diverse regions and subtypes, but also a methodological prototype for scalable MA network construction. By transitioning from conventional pairwise analyses to structured one-to-many and many-to-many dietary-cancer subnetwork models, the analysis exemplifies how interpretable, domain-specific modules can be extracted from complex, high-dimensional global datasets. The network-based approach enables the detection of both known and underexplored interactions, including antagonistic and synergistic patterns that are often masked in linear analytical frameworks. Although modest in scope, this work serves as a conceptual contribution to the broader development of MA networks, illustrating a generalizable strategy for transforming fragmented global data into coherent, actionable knowledge systems. As such, it marks an incremental but meaningful step toward the systematic, cross-domain deployment of interpretable network analytics in public health and beyond.

## RESOURCE AVAILABILITY

### Lead contact

Further information and reasonable requests for resources and reagents should be directed to and will be fulfilled by the lead contact, Hongyue Ma.

### Materials availability

This study did not generate new samples or unique reagents.

### Data and code availability

***• Data***: This study is based exclusively on publicly available datasets, all of which are detailed in the Methods section.
***• Code***: This paper does not report original code.
***• Additional Information***: Any additional information required to reanalyze the data reported in this article is available from the lead contact upon request.

## Data Availability

This study is based exclusively on publicly available datasets, all of which are detailed in the Methods section.

## ACKNOWLEDGMENTS

None.

## AUTHOR CONTRIBUTIONS

Hongyue Ma performed all analyses and wrote the manuscript.

## DECLARATION OF INTERESTS

The author declares no competing interests.

## Notes

### Competing Interest Statement

The authors have declared no competing interest.

