## Supplementary Materials for "Mapping Global Dietary Risk Factors for Cancer: A Structured Network-Based Analysis"

**Table S1.** Association between dietary factors and overall cancer incidence.

| Items | Correlation Coefficient | | | | | | Linear Regression | |
|---|---|---|---|---|---|---|---|---|
| | CC | p-value | N | Partial CC | p-value | N | $R^2$ | N |
| **Red meat** | **0.769** | **0.000** | **155** | **0.600** | **0.000** | **140** | **0.5906** | **155** |
| **Alcoholic beverages** | **0.768** | **0.000** | **155** | **0.620** | **0.000** | **140** | **0.5904** | **155** |
| **Animal fats** | **0.717** | **0.000** | **155** | **0.500** | **0.000** | **140** | **0.5148** | **155** |
| **Milk** | **0.714** | **0.000** | **155** | **0.498** | **0.000** | **140** | **0.5099** | **155** |
| **Eggs** | **0.658** | **0.000** | **155** | **0.349** | **0.000** | **140** | **0.4332** | **155** |
| **Sugar and sweeteners** | **0.530** | **0.000** | **155** | **0.264** | **0.002** | **140** | **0.2807** | **155** |
| Vegetable oils | 0.423 | 0.000 | 155 | 0.016 | 0.849 | 140 | 0.1793 | 155 |
| Nuts | 0.373 | 0.000 | 155 | 0.136 | 0.111 | 140 | 0.1395 | 155 |
| Poultry meat | 0.333 | 0.000 | 155 | 0.031 | 0.717 | 140 | 0.1109 | 155 |
| Vegetables | 0.316 | 0.000 | 155 | 0.018 | 0.829 | 140 | 0.0996 | 155 |
| Pulses | -0.298 | 0.000 | 155 | -0.119 | 0.164 | 140 | 0.0888 | 155 |
| Cereals | -0.289 | 0.000 | 155 | -0.276 | 0.001 | 140 | 0.0835 | 155 |
| Fish and seafood | 0.155 | 0.053 | 155 | 0.028 | 0.742 | 140 | 0.0242 | 155 |
| Starchy roots | -0.148 | 0.067 | 155 | 0.040 | 0.642 | 140 | 0.0218 | 155 |
| Fruit | 0.059 | 0.469 | 155 | -0.106 | 0.215 | 140 | 0.0034 | 155 |
| Other meat | 0.052 | 0.523 | 155 | 0.091 | 0.289 | 140 | 0.0027 | 155 |

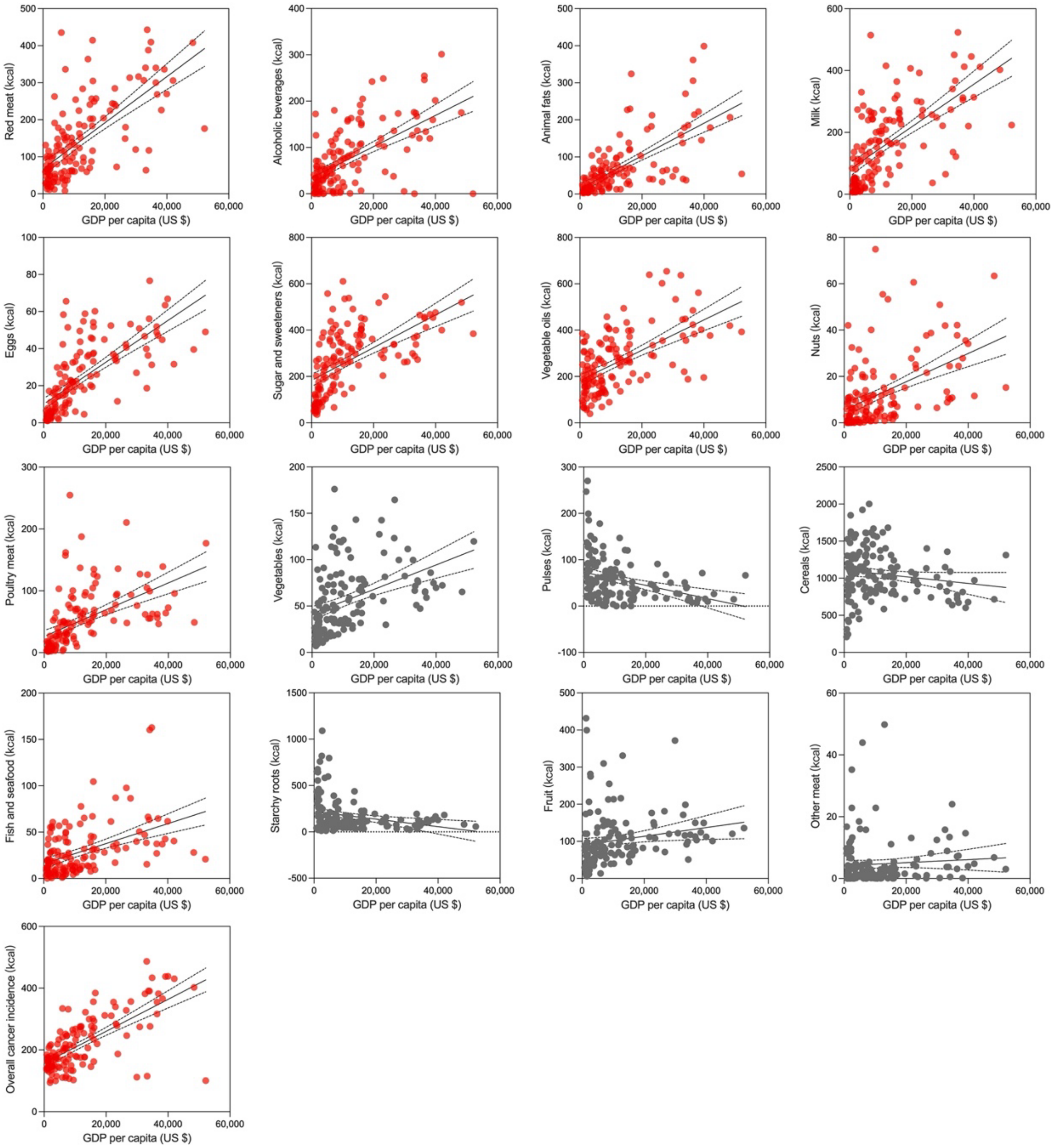


**Figure S1.** Regression analysis of association between dietary factors or OCI and GDP per capita.

**Table S2.** Association between dietary factors or OCI and GDP per capita.

| Food Type | Correlation Coefficient | | | Linear Regression | |
|---|---|---|---|---|---|
| | CC | p-value | N | $R^2$ | N |
| Red meat | 0.676 | 0.000 | 140 | 0.4575 | 140 |
| Alcoholic beverages | 0.595 | 0.000 | 140 | 0.3538 | 140 |
| Animal fats | 0.680 | 0.000 | 140 | 0.4629 | 140 |
| Milk | 0.639 | 0.000 | 140 | 0.4087 | 140 |
| Eggs | 0.707 | 0.000 | 140 | 0.5001 | 140 |
| Sugar and sweeteners | 0.584 | 0.000 | 140 | 0.3414 | 140 |
| Vegetable oils | 0.586 | 0.000 | 140 | 0.3438 | 140 |
| Nuts | 0.483 | 0.000 | 140 | 0.2336 | 140 |
| Poultry meat | 0.536 | 0.000 | 140 | 0.2869 | 140 |
| Vegetables | 0.426 | 0.000 | 140 | 0.1812 | 140 |
| Pulses | -0.342 | 0.000 | 140 | 0.1167 | 140 |
| Cereals | -0.155 | 0.067 | 140 | 0.0240 | 140 |
| Fish and seafood | 0.460 | 0.000 | 140 | 0.2168 | 140 |
| Starchy roots | -0.257 | 0.002 | 140 | 0.0661 | 140 |
| Fruit | 0.184 | 0.029 | 140 | 0.0339 | 140 |
| Other meat | 0.074 | 0.386 | 140 | 0.0055 | 140 |
| OCI | 0.689 | 0.000 | 140 | 0.4747 | 140 |

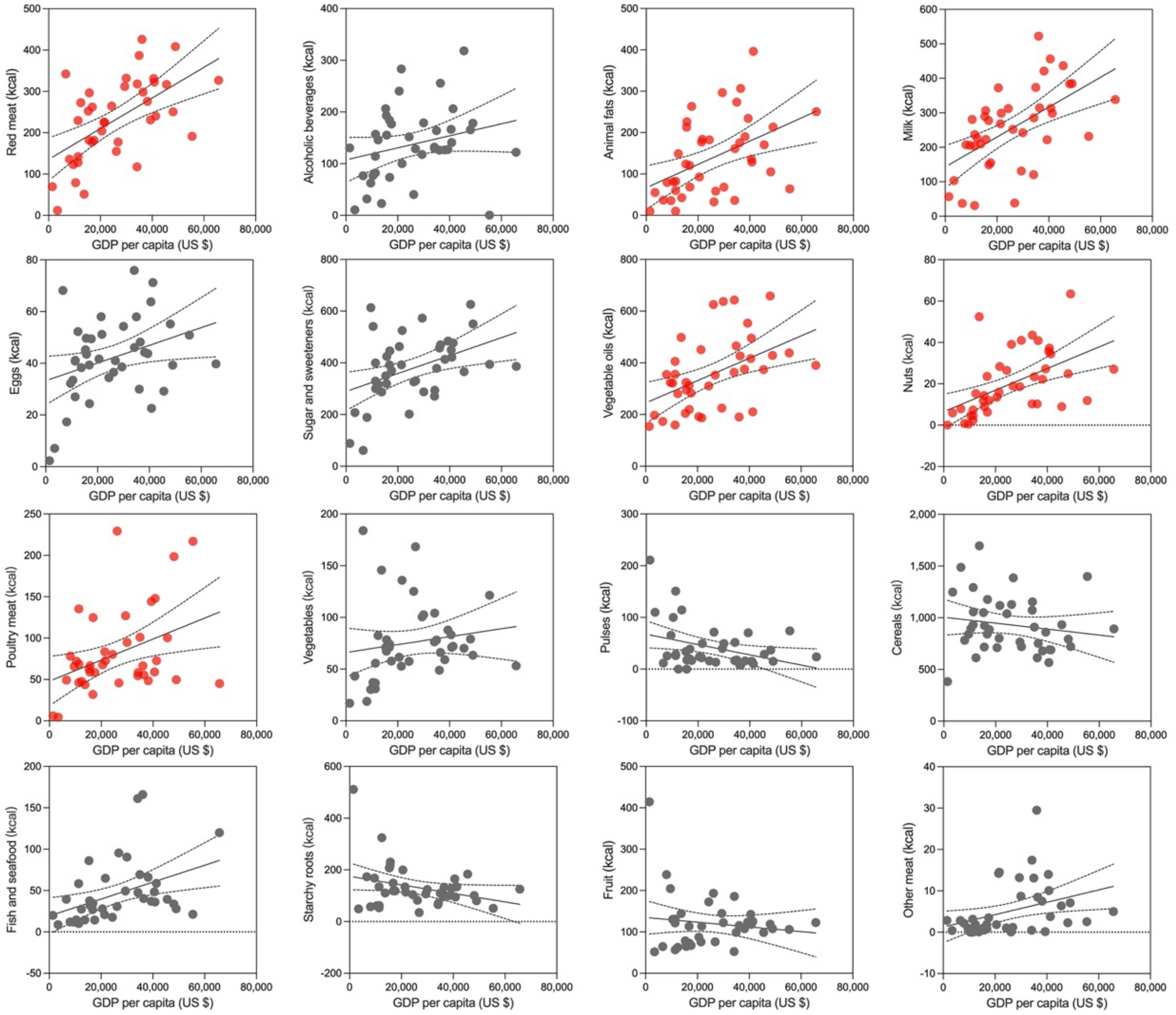


**Figure S2.** Regression analysis of association between dietary factors and GDP per capita.

**Table S3.** Association between dietary factors and GDP per capita.

| Food Type (n=40) | Correlation Coefficient | | Linear Regression |
|---|---|---|---|
| | CC | p-value | $R^2$ |
| Red meat | 0.577 | 0.000 | 0.3335 |
| Alcoholic beverages | 0.254 | 0.114 | 0.0643 |
| Animal fats | 0.465 | 0.002 | 0.2163 |
| Milk | 0.569 | 0.000 | 0.3237 |
| Eggs | 0.339 | 0.032 | 0.1149 |
| Sugar and sweeteners | 0.420 | 0.007 | 0.1766 |
| Vegetable oils | 0.473 | 0.002 | 0.2236 |
| Nuts | 0.525 | 0.001 | 0.2754 |
| Poultry meat | 0.389 | 0.013 | 0.1510 |
| Vegetables | 0.158 | 0.331 | 0.0249 |
| Pulses | -0.352 | 0.026 | 0.1236 |
| Cereals | -0.162 | 0.319 | 0.0261 |
| Fish and seafood | 0.418 | 0.007 | 0.1749 |
| Starchy roots | -0.299 | 0.061 | 0.0895 |
| Fruit | -0.136 | 0.401 | 0.0186 |
| Other meat | 0.361 | 0.022 | 0.1304 |

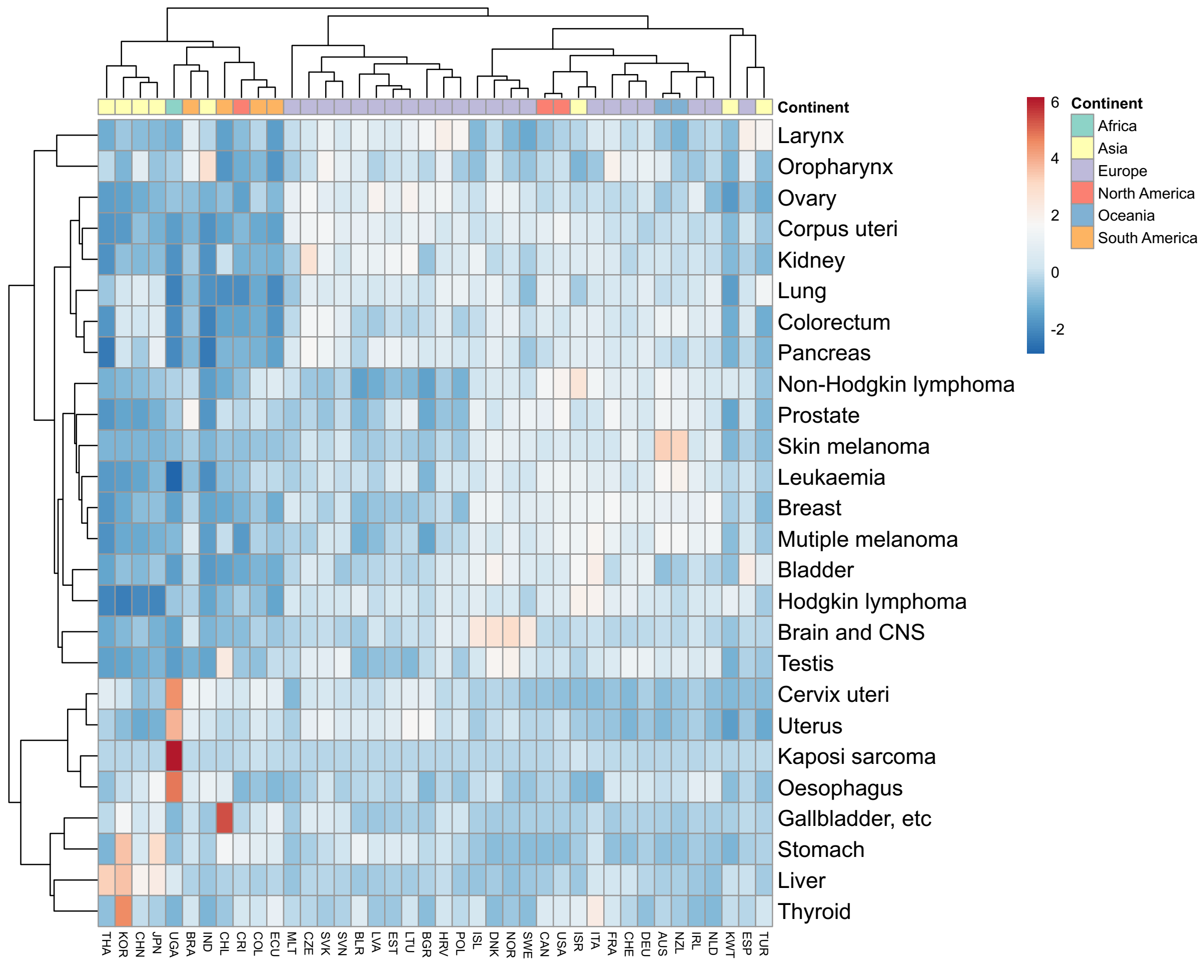


**Figure S3.** Heatmap showing the distribution of 26 cancer types across 40 countries or regions.

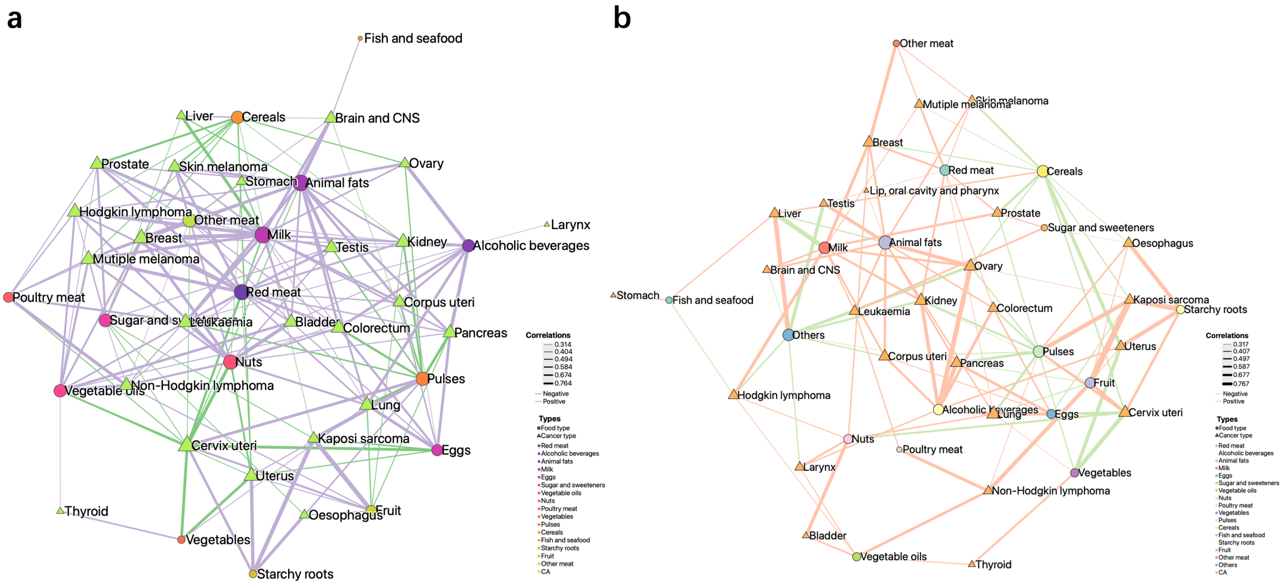


**Figure S4.** Network representations of the associations between 16 dietary components and 26 cancer types. (a) One-star association network. (b) Two-star association network constructed after adjustment for GDP per capita.

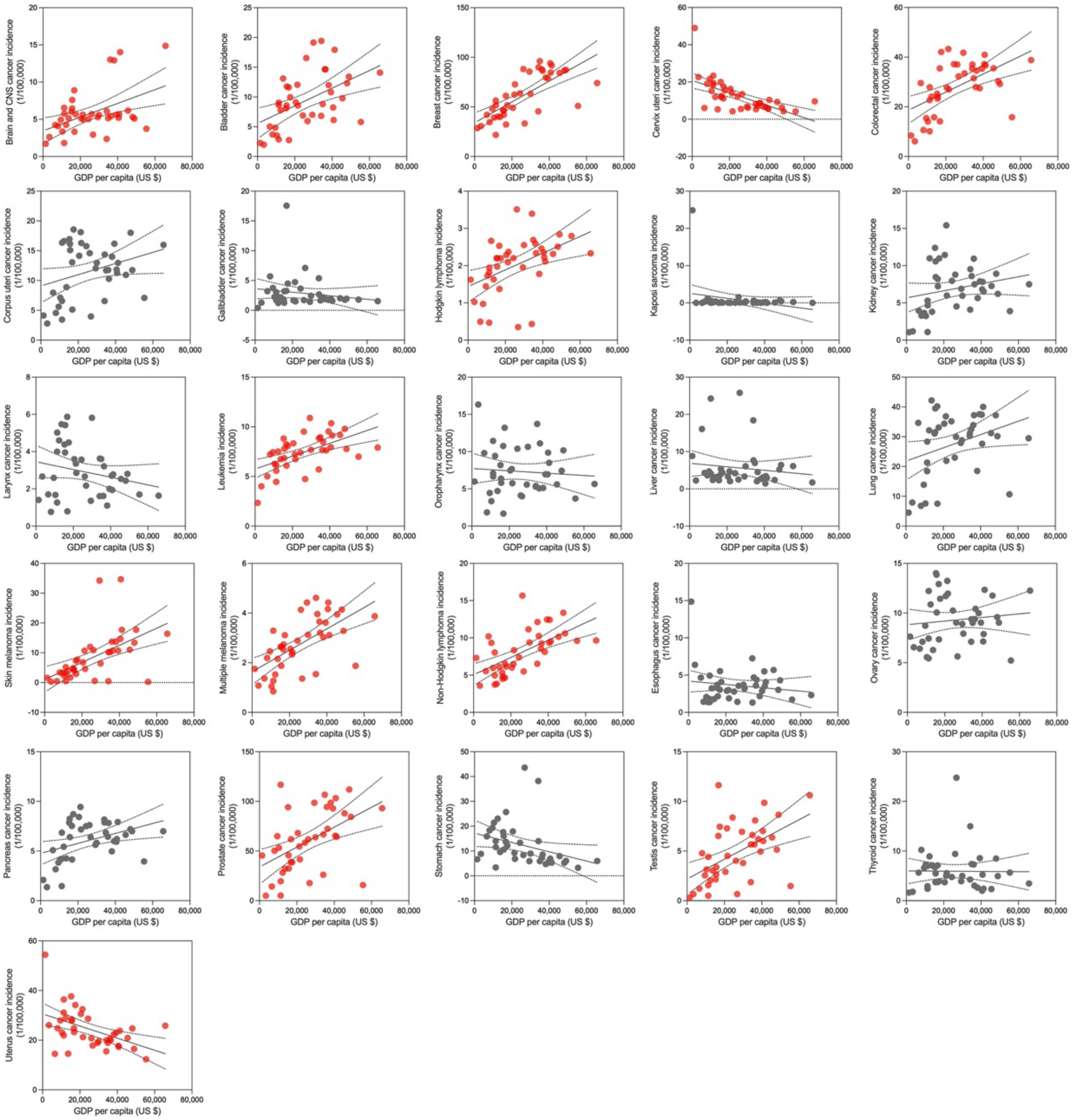


**Figure S5.** Regression analysis of association between cancer incidences and GDP per capita.

**Table S4.** Association between cancer incidences and GDP per capita.

| Cancer Type (n=40) | Correlation Coefficient | | Linear Regression |
|---|---|---|---|
| | CC | p-value | R squared |
| Brain and CNS | 0.468 | 0.002 | 0.2192 |
| Bladder | 0.496 | 0.001 | 0.2460 |
| Breast | 0.724 | 0.000 | 0.5248 |
| Cervix uteri | -0.623 | 0.000 | 0.3881 |
| Colorectum | 0.548 | 0.000 | 0.3001 |
| Corpus uteri | 0.308 | 0.053 | 0.0950 |
| Esophagus | -0.147 | 0.364 | 0.0218 |
| Gallbladder, etc. | -0.167 | 0.304 | 0.0277 |
| Hodgkin lymphoma | 0.463 | 0.003 | 0.2142 |
| Kaposi sarcoma | -0.259 | 0.106 | 0.0673 |
| Kidney | 0.226 | 0.160 | 0.0513 |
| Larynx | -0.224 | 0.164 | 0.0504 |
| Leukemia | 0.560 | 0.000 | 0.3135 |
| Liver | -0.129 | 0.428 | 0.0166 |
| Lung | 0.323 | 0.042 | 0.1046 |
| Skin melanoma | 0.553 | 0.000 | 0.3055 |
| Multiple melanoma | 0.625 | 0.000 | 0.3908 |
| Non-Hodgkin lymphoma | 0.616 | 0.000 | 0.3790 |
| Oropharynx | -0.074 | 0.650 | 0.0055 |
| Ovary | 0.112 | 0.490 | 0.0126 |
| Pancreas | 0.389 | 0.013 | 0.1514 |
| Prostate | 0.493 | 0.001 | 0.2426 |
| Stomach | -0.329 | 0.038 | 0.1080 |
| Testis | 0.523 | 0.001 | 0.2736 |
| Thyroid | -0.009 | 0.956 | 0.0001 |
| Uterus | -0.482 | 0.002 | 0.2324 |

#### Correlation and Linear Regression Analysis of associations between 16 dietary factors and 26 types of cancer (Figure S4-Figure S29, Table S4-Table S29)

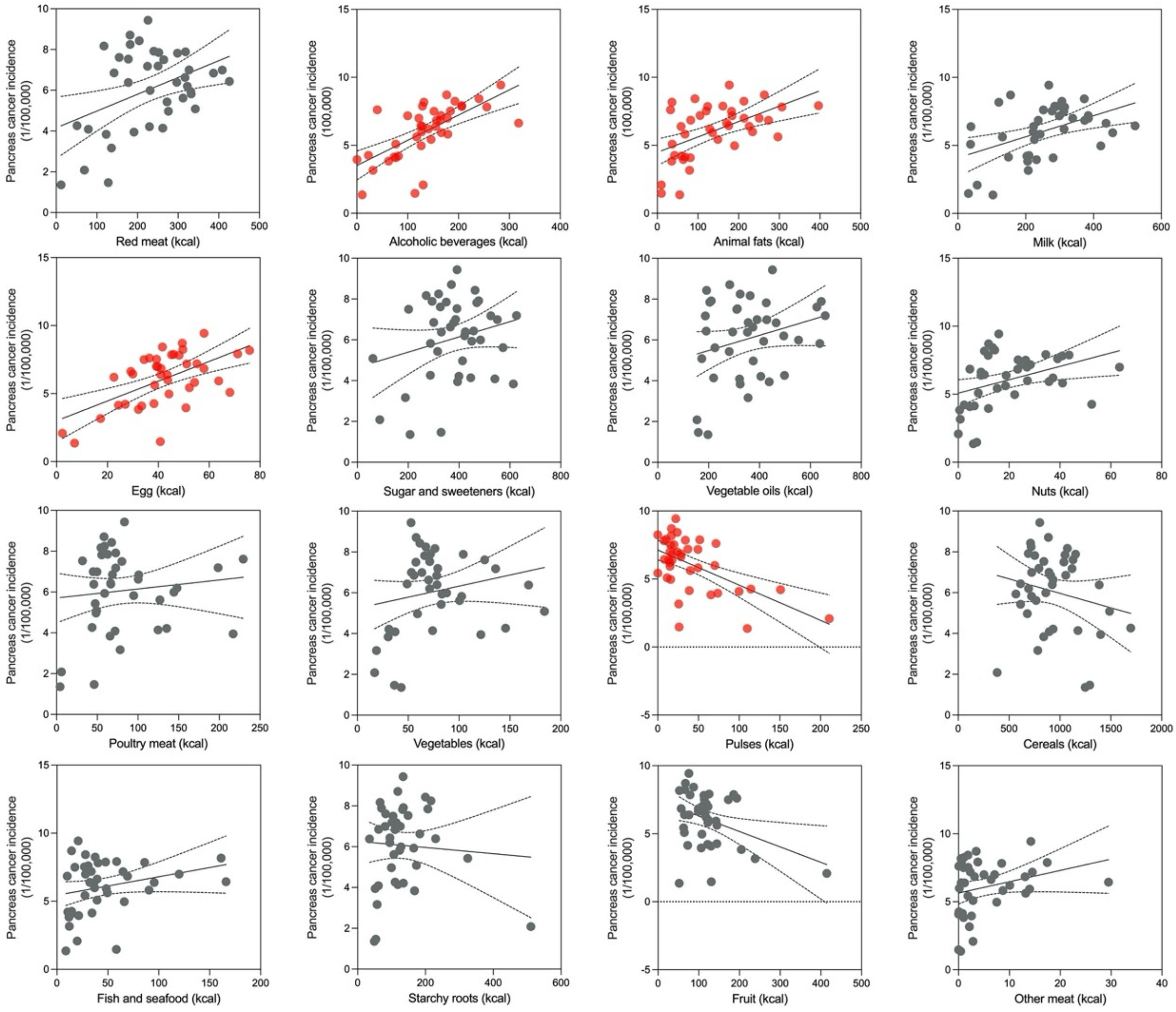


**Figure S6.** Regression analysis of association between 16 dietary factors and pancreas cancer incidence.

**Table S5.** Association between dietary factors and pancreas cancer incidence.

| Food Type (n=40) | Correlation Coefficient | | | | Linear Regression |
|---|---|---|---|---|---|
| | **CC** | **p-value** | **Partial CC** | **p-value** | $R^2$ |
| Red meat | 0.411 | 0.008 | 0.248 | 0.128 | 0.1692 |
| **Alcoholic beverages*** | **0.660** | **0.000** | **0.630** | **0.000** | **0.4352** |
| **Animal fats*** | **0.533** | **0.000** | **0.431** | **0.006** | **0.2837** |
| Milk | 0.458 | 0.003 | 0.312 | 0.053 | 0.2097 |
| **Eggs*** | **0.569** | **0.000** | **0.505** | **0.001** | **0.3243** |
| Sugar and sweeteners | 0.240 | 0.136 | 0.091 | 0.581 | 0.0574 |
| Vegetable oils | 0.264 | 0.099 | 0.099 | 0.549 | 0.0699 |
| Nuts | 0.381 | 0.015 | 0.225 | 0.169 | 0.1448 |
| Poultry meat | 0.113 | 0.489 | -0.046 | 0.783 | 0.0127 |
| Vegetables | 0.205 | 0.204 | 0.158 | 0.337 | 0.0421 |
| **Pulses*** | **-0.569** | **0.000** | **-0.502** | **0.001** | **0.3242** |
| Cereals | -0.197 | 0.224 | -0.147 | 0.372 | 0.0386 |
| Fish and seafood | 0.258 | 0.109 | 0.113 | 0.492 | 0.0663 |
| Starchy roots | -0.065 | 0.689 | 0.058 | 0.724 | 0.0042 |
| **Fruit*** | **-0.368** | **0.020** | **-0.345** | **0.032** | **0.1351** |
| Other meat | 0.268 | 0.095 | 0.148 | 0.369 | 0.0716 |

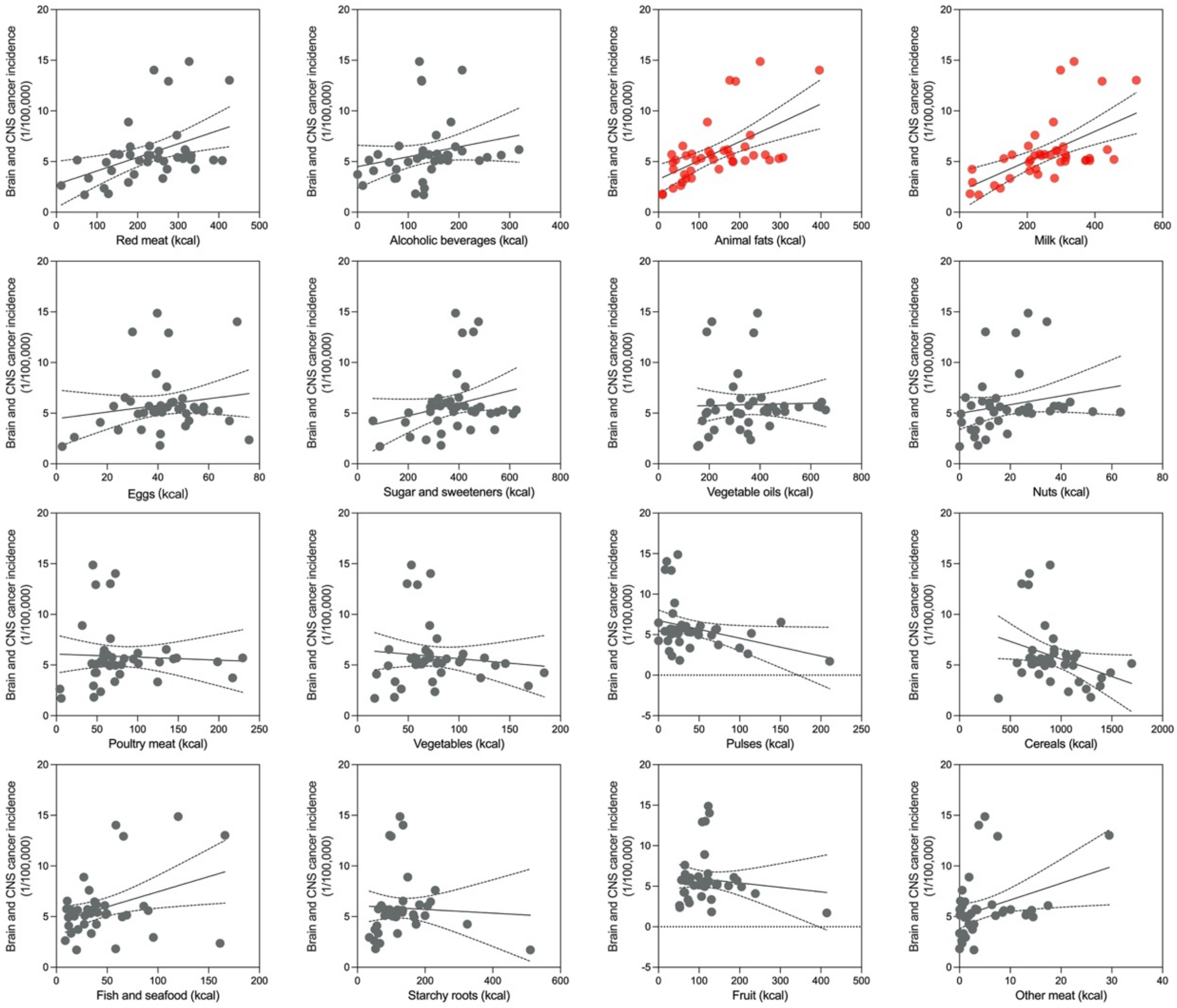


**Figure S7.** Regression analysis of association between 16 dietary factors and brain (CNS) cancer incidence.

**Table S6.** Association between dietary factors and brain (CNS) cancer incidence.

| Food Type (n=40) | Correlation Coefficient | | | | Linear Regression |
|---|---|---|---|---|---|
| | **CC** | **p-value** | **Partial CC** | **p-value** | **R squared** |
| Red meat | 0.436 | 0.005 | 0.230 | 0.160 | 0.1900 |
| Alcoholic beverages | 0.227 | 0.159 | 0.127 | 0.443 | 0.0515 |
| **Animal fats*** | **0.572** | **0.000** | **0.453** | **0.004** | **0.3273** |
| **Milk*** | **0.571** | **0.000** | **0.419** | **0.008** | **0.3256** |
| Eggs | 0.167 | 0.302 | 0.010 | 0.951 | 0.0280 |
| Sugar and sweeteners | 0.260 | 0.105 | 0.079 | 0.631 | 0.0678 |
| Vegetable oils | 0.027 | 0.869 | -0.250 | 0.125 | 0.0007 |
| Nuts | 0.217 | 0.178 | -0.038 | 0.820 | 0.0473 |
| Poultry meat | -0.049 | 0.762 | -0.284 | 0.080 | 0.0024 |
| Vegetables | -0.109 | 0.504 | -0.209 | 0.201 | 0.0118 |
| Pulses | -0.316 | 0.047 | -0.183 | 0.266 | 0.0996 |
| Cereals | -0.314 | 0.049 | -0.273 | 0.092 | 0.0985 |
| Fish and seafood | 0.370 | 0.019 | 0.217 | 0.185 | 0.1368 |
| Starchy roots | -0.052 | 0.752 | 0.105 | 0.525 | 0.0027 |
| Fruit | -0.116 | 0.476 | -0.059 | 0.719 | 0.0134 |
| Other meat | 0.346 | 0.029 | 0.215 | 0.189 | 0.1199 |

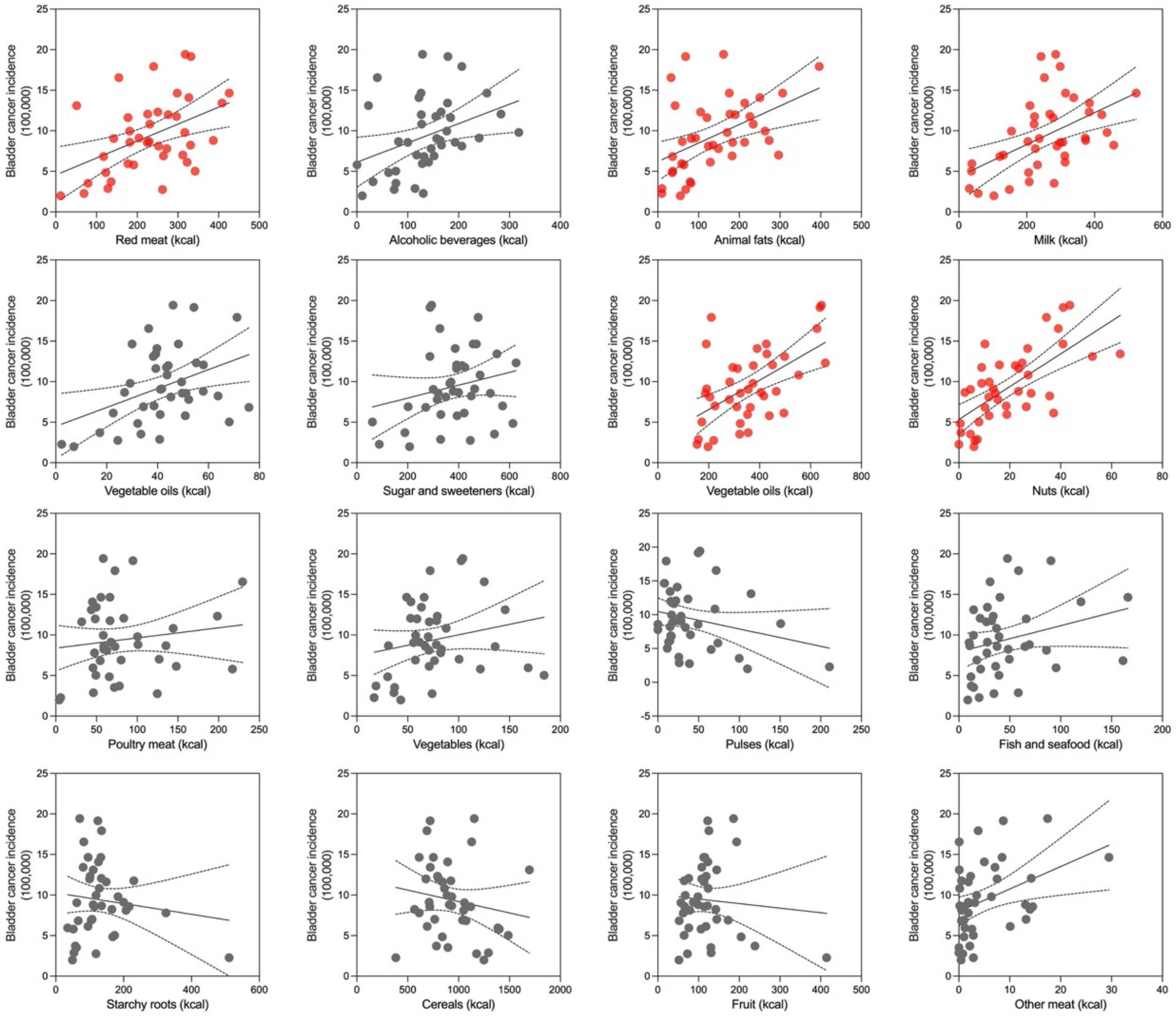


**Figure S8.** Regression analysis of association between 16 dietary factors and bladder cancer incidence.

**Table S7.** Association between dietary factors and bladder cancer incidence.

| **Food Type (n=40)** | **Correlation Coefficient** | | | | **Linear Regression** |
|---|---|---|---|---|---|
| | **CC** | **p-value** | **Partial CC** | **p-value** | **R squared** |
| Red meat | 0.450 | 0.004 | 0.231 | 0.158 | 0.2024 |
| Alcoholic beverages | 0.366 | 0.02 | 0.286 | 0.078 | 0.1337 |
| Animal fats | 0.464 | 0.003 | 0.303 | 0.061 | 0.2151 |
| Milk | 0.502 | 0.001 | 0.308 | 0.057 | 0.2518 |
| Eggs | 0.394 | 0.012 | 0.276 | 0.089 | 0.1550 |
| Sugar and sweeteners | 0.219 | 0.174 | 0.014 | 0.934 | 0.0480 |
| **Vegetable oils*** | **0.553** | **0.000** | **0.417** | **0.008** | **0.3062** |
| **Nuts*** | **0.673** | **0.000** | **0.559** | **0.000** | **0.4534** |
| Poultry meat | 0.137 | 0.399 | -0.07 | 0.674 | 0.0188 |
| Vegetables | 0.21 | 0.194 | 0.153 | 0.351 | 0.0440 |
| Pulses | -0.244 | 0.129 | -0.086 | 0.604 | 0.0595 |
| Cereals | -0.168 | 0.299 | -0.103 | 0.533 | 0.0284 |
| Fish and seafood | 0.264 | 0.100 | 0.072 | 0.664 | 0.0697 |
| Starchy roots | -0.122 | 0.453 | 0.032 | 0.848 | 0.0149 |
| Fruit | -0.079 | 0.629 | -0.013 | 0.937 | 0.0062 |
| Other meat | 0.383 | 0.015 | 0.252 | 0.121 | 0.1470 |

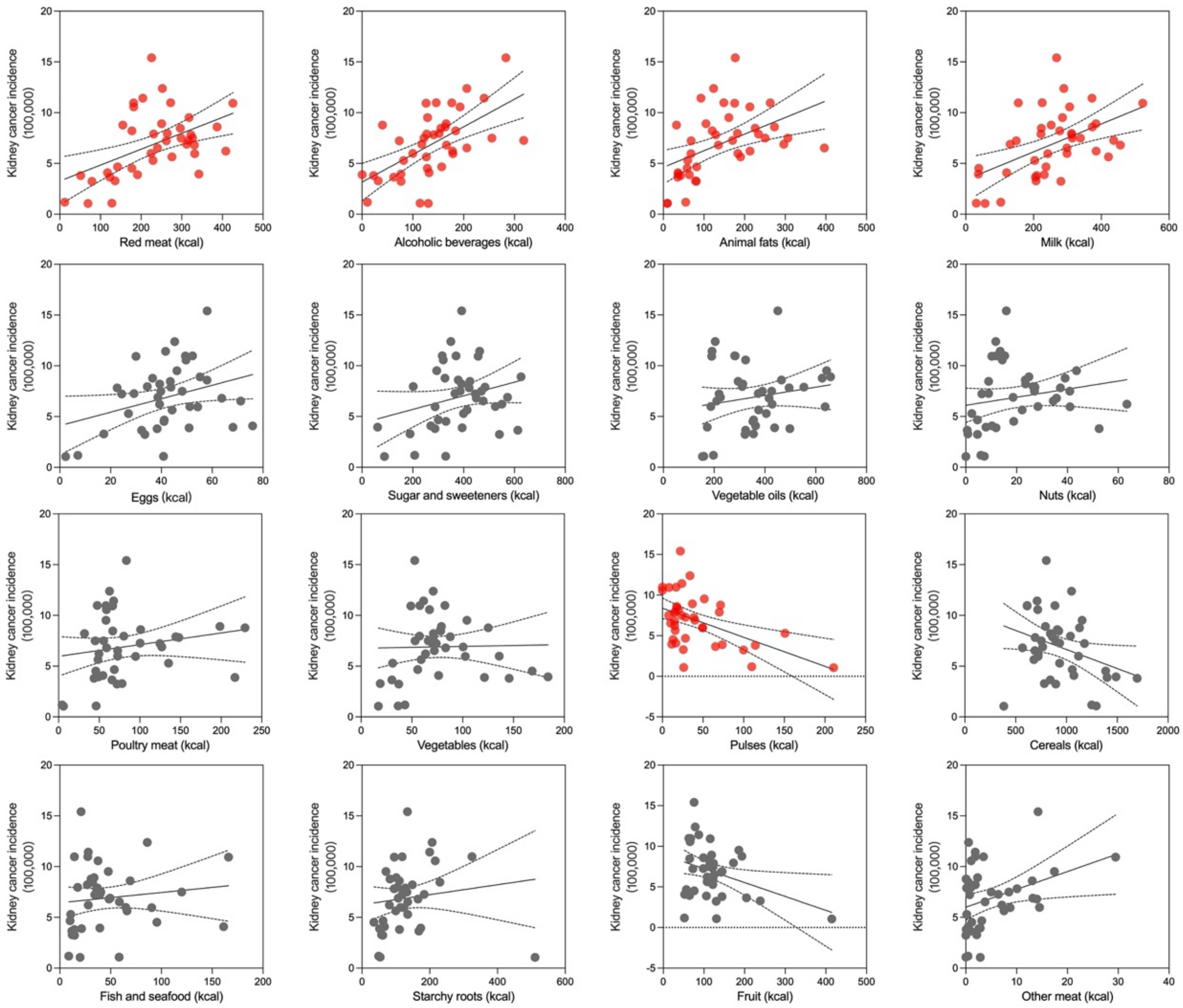


**Figure S9.** Regression analysis of association between 16 dietary factors and kidney cancer incidence.

**Table S8.** Association between dietary factors and kidney cancer incidence.

| Food Type (n=40) | Correlation Coefficient | | | | Linear Regression |
|---|---|---|---|---|---|
| | CC | p-value | Partial CC | p-value | R squared |
| **Red meat*** | **0.483** | **0.002** | **0.443** | **0.005** | **0.2330** |
| **Alcoholic beverages*** | **0.598** | **0.000** | **0.574** | **0.000** | **0.3574** |
| **Animal fats*** | **0.475** | **0.002** | **0.428** | **0.007** | **0.2254** |
| **Milk*** | **0.502** | **0.001** | **0.466** | **0.003** | **0.2520** |
| Eggs | 0.321 | 0.044 | 0.266 | 0.102 | 0.1027 |
| Sugar and sweeteners | 0.266 | 0.098 | 0.193 | 0.239 | 0.0706 |
| Vegetable oils | 0.178 | 0.271 | 0.083 | 0.616 | 0.0317 |
| Nuts | 0.190 | 0.241 | 0.086 | 0.604 | 0.0361 |
| Poultry meat | 0.180 | 0.265 | 0.103 | 0.533 | 0.0325 |
| Vegetables | 0.021 | 0.899 | -0.016 | 0.925 | 0.0004 |
| **Pulses*** | **-0.486** | **0.001** | **-0.446** | **0.004** | **0.2365** |
| Cereals | -0.321 | 0.043 | -0.296 | 0.067 | 0.1032 |
| Fish and seafood | 0.118 | 0.468 | 0.026 | 0.873 | 0.0139 |
| Starchy roots | 0.129 | 0.426 | 0.212 | 0.195 | 0.0168 |
| **Fruit*** | **-0.343** | **0.030** | **-0.323** | **0.045** | **0.1176** |
| Other meat | 0.348 | 0.028 | 0.293 | 0.07 | 0.1213 |

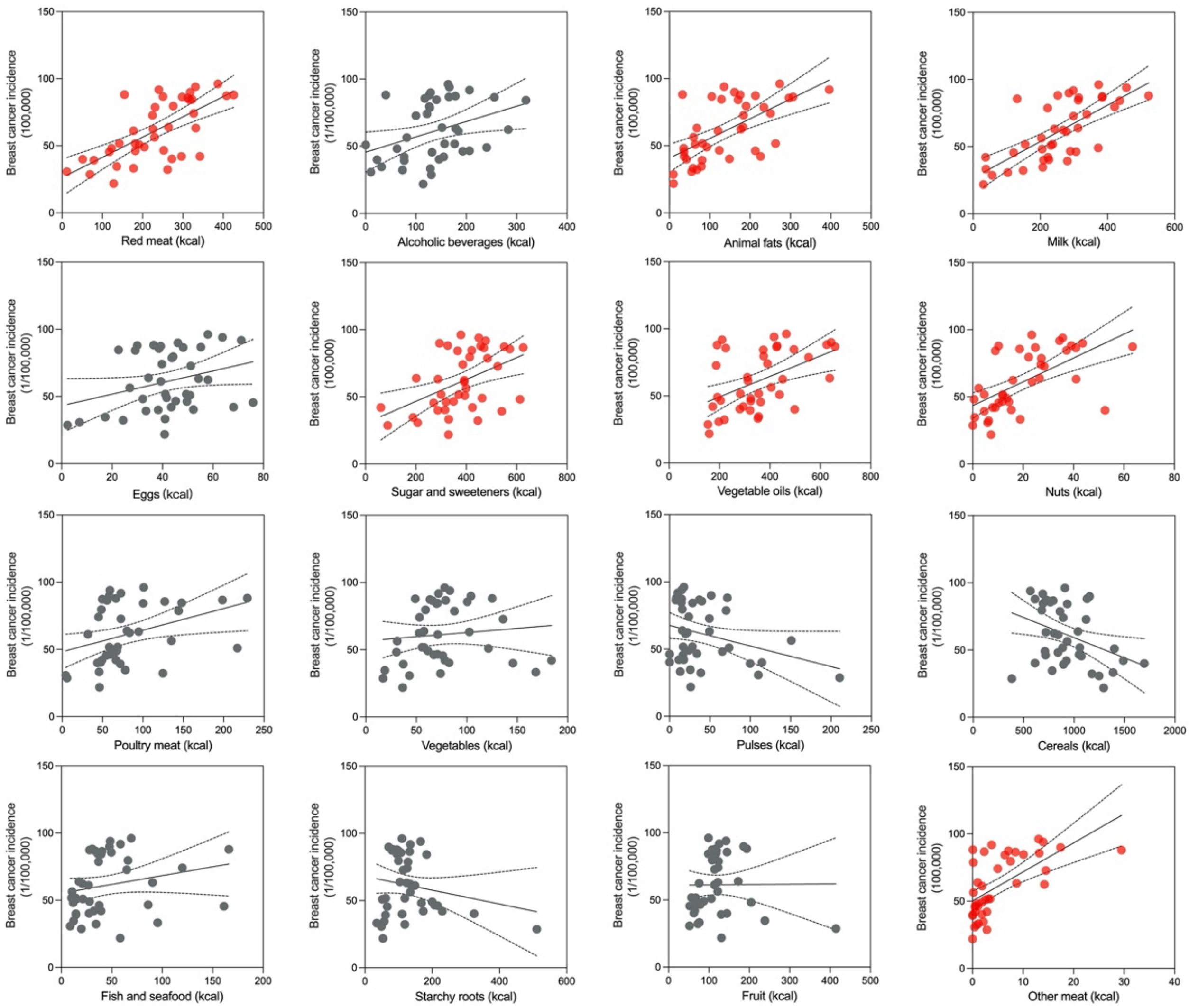


**Figure S10.** Regression analysis of association between 16 dietary factors and breast cancer incidence.

**Table S9.** Association between dietary factors and breast cancer incidence.

| Food Type (n=40) | Correlation Coefficient | | | | Linear Regression |
|---|---|---|---|---|---|
| | CC | p-value | Partial CC | p-value | R squared |
| **Red meat*** | **0.669** | **0.000** | **0.446** | **0.004** | **0.4482** |
| Alcoholic beverages | 0.36 | 0.022 | 0.265 | 0.103 | 0.1299 |
| **Animal fats*** | **0.613** | **0.000** | **0.453** | **0.004** | **0.3761** |
| **Milk*** | **0.713** | **0.000** | **0.531** | **0.001** | **0.5085** |
| Eggs | 0.300 | 0.06 | 0.084 | 0.610 | 0.0902 |
| Sugar and sweeteners | 0.464 | 0.003 | 0.255 | 0.116 | 0.2155 |
| Vegetable oils | 0.482 | 0.002 | 0.229 | 0.160 | 0.2321 |
| **Nuts*** | **0.609** | **0.000** | **0.391** | **0.014** | **0.3713** |
| Poultry meat | 0.363 | 0.021 | 0.129 | 0.434 | 0.1321 |
| Vegetables | 0.105 | 0.52 | -0.014 | 0.933 | 0.0110 |
| Pulses | -0.300 | 0.06 | -0.070 | 0.672 | 0.0898 |
| **Cereals*** | **-0.370** | **0.019** | **-0.371** | **0.020** | **0.1367** |
| Fish and seafood | 0.221 | 0.171 | -0.132 | 0.425 | 0.0487 |
| Starchy roots | -0.196 | 0.225 | 0.031 | 0.851 | 0.0385 |
| Fruit | 0.007 | 0.965 | 0.155 | 0.345 | 0.0001 |
| **Other meat*** | **0.615** | **0.000** | **0.550** | **0.000** | **0.3788** |

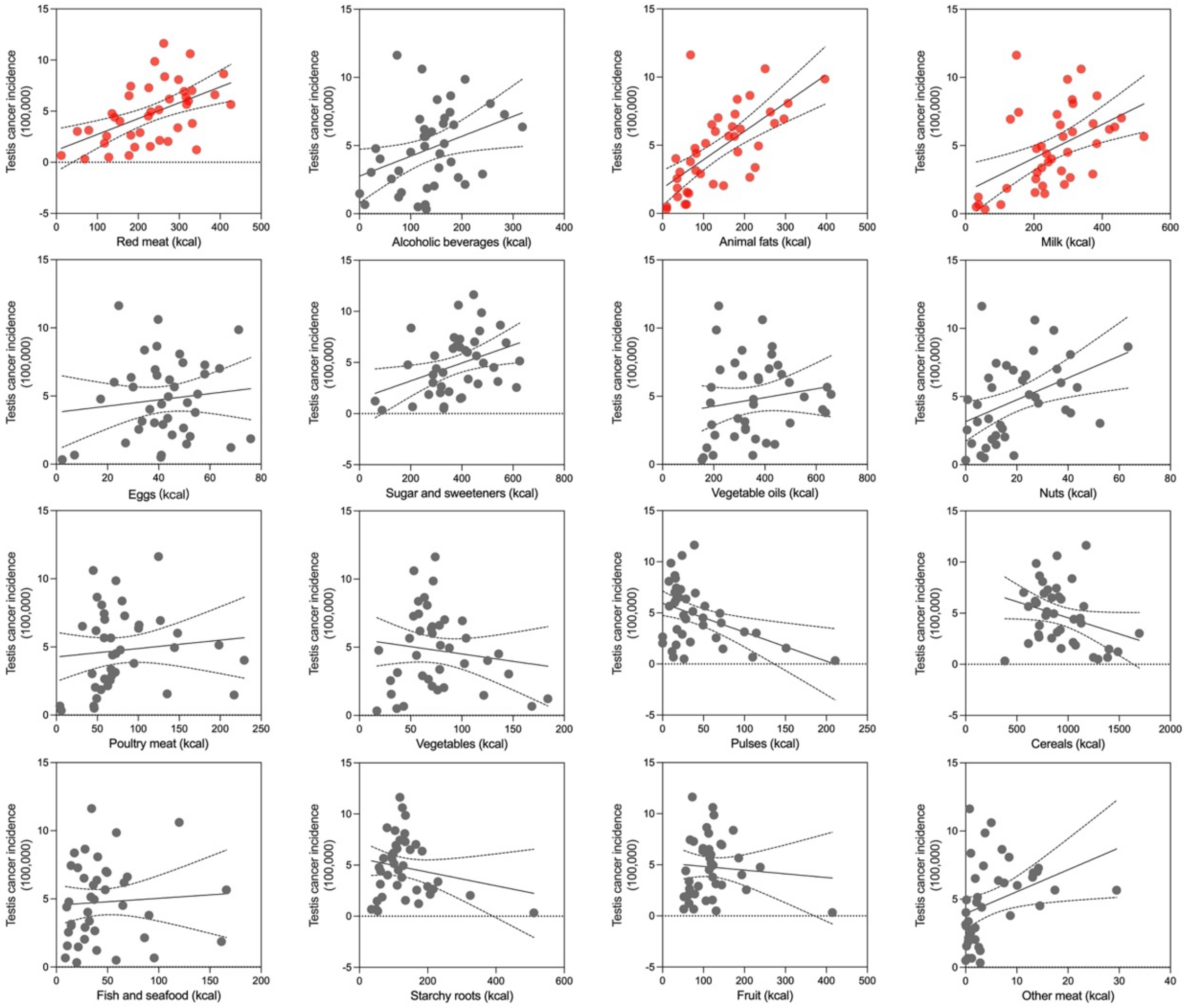


**Figure S11.** Regression analysis of association between 16 dietary factors and testis cancer incidence.

**Table S10.** Association between dietary factors and testis cancer incidence.

| Food Type (n=40) | Correlation Coefficient | | | | Linear Regression |
|---|---|---|---|---|---|
| | CC | p-value | Partial CC | p-value | R squared |
| **Red meat*** | **0.351** | **0.026** | **0.318** | **0.049** | **0.2738** |
| Alcoholic beverages | 0.523 | 0.001 | 0.265 | 0.103 | 0.1235 |
| **Animal fats*** | **0.663** | **0.000** | **0.556** | **0.000** | **0.4398** |
| Milk | 0.494 | 0.001 | 0.281 | 0.083 | 0.2445 |
| Eggs | 0.121 | 0.456 | -0.070 | 0.673 | 0.0147 |
| Sugar and sweeteners | 0.380 | 0.016 | 0.207 | 0.205 | 0.1445 |
| Vegetable oils | 0.155 | 0.340 | -0.123 | 0.455 | 0.0240 |
| Nuts | 0.419 | 0.007 | 0.199 | 0.224 | 0.1756 |
| Poultry meat | 0.106 | 0.515 | -0.124 | 0.452 | 0.0112 |
| Vegetables | -0.137 | 0.398 | -0.261 | 0.108 | 0.0188 |
| Pulses | -0.423 | 0.007 | -0.299 | 0.064 | 0.1786 |
| Cereals | -0.296 | 0.063 | -0.252 | 0.122 | 0.0879 |
| Fish and seafood | 0.065 | 0.690 | -0.199 | 0.225 | 0.0042 |
| Starchy roots | -0.193 | 0.232 | -0.045 | 0.784 | 0.0374 |
| Fruit | -0.080 | 0.625 | -0.010 | 0.953 | 0.0063 |
| Other meat | 0.350 | 0.027 | 0.203 | 0.216 | 0.1224 |

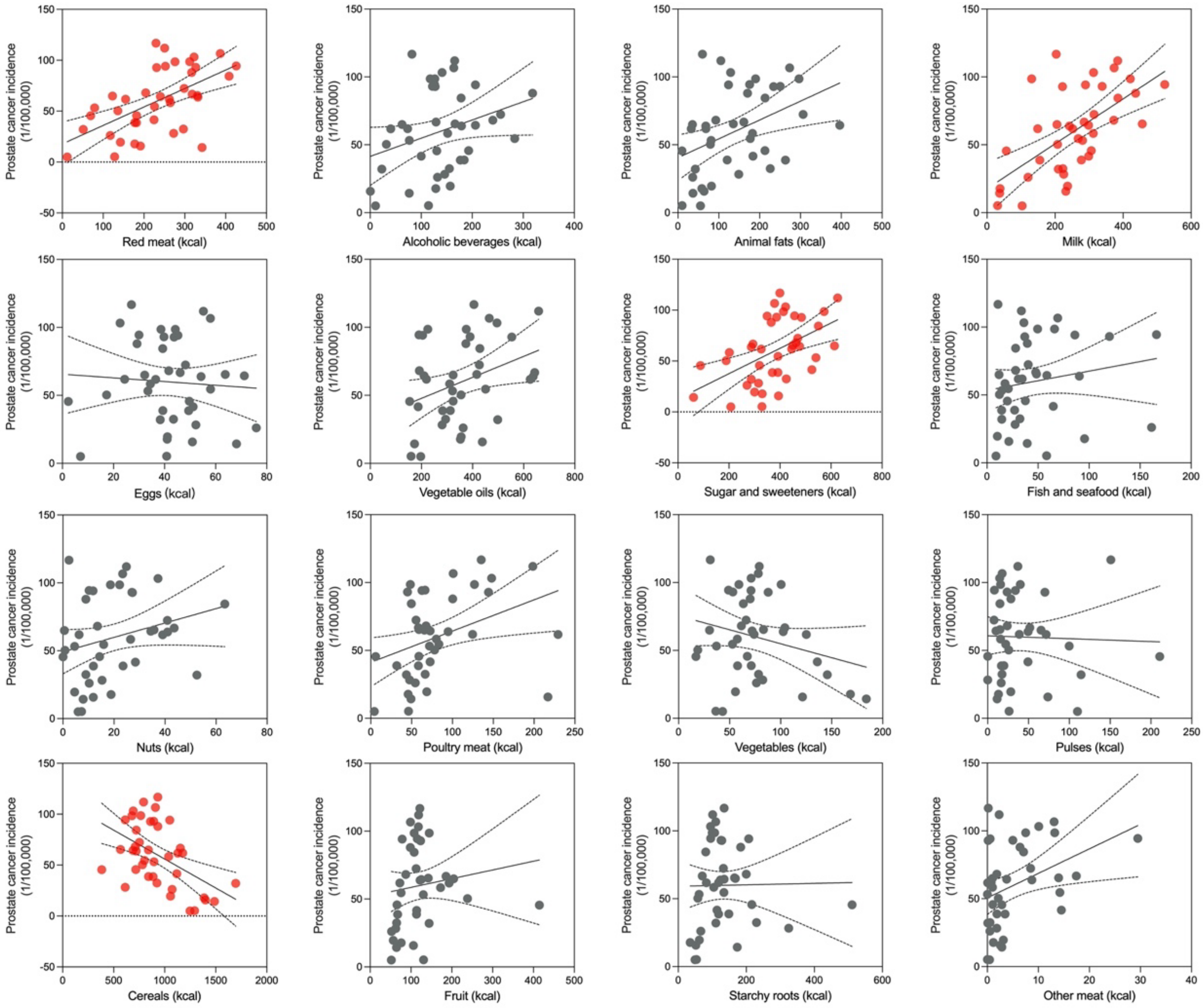


**Figure S12.** Regression analysis of association between 16 dietary factors and prostate cancer incidence.

**Table S11.** Association between dietary factors and prostate cancer incidence.

| **Food Type (n=40)** | **Correlation Coefficient** | | | | **Linear Regression** |
|---|---|---|---|---|---|
| | **CC** | **p-value** | **Partial CC** | **p-value** | **R squared** |
| **Red meat*** | **0.573** | **0.000** | **0.406** | **0.010** | **0.3282** |
| Alcoholic beverages | 0.302 | 0.058 | 0.210 | 0.199 | 0.0912 |
| Animal fats | 0.413 | 0.008 | 0.239 | 0.143 | 0.1708 |
| **Milk*** | **0.619** | **0.000** | **0.474** | **0.002** | **0.3838** |
| Eggs | -0.067 | 0.681 | -0.286 | 0.078 | 0.0045 |
| **Sugar and sweeteners*** | **0.505** | **0.001** | **0.377** | **0.018** | **0.2546** |
| Vegetable oils | 0.347 | 0.028 | 0.149 | 0.366 | 0.1205 |
| Nuts | 0.259 | 0.106 | 0.001 | 0.996 | 0.0672 |
| Poultry meat | 0.37 | 0.019 | 0.223 | 0.172 | 0.1371 |
| Vegetables | -0.245 | 0.127 | -0.376 | 0.018 | 0.0601 |
| Pulses | -0.029 | 0.857 | 0.177 | 0.282 | 0.0009 |
| **Cereals*** | **-0.498** | **0.001** | **-0.488** | **0.002** | **0.2484** |
| Fish and seafood | 0.17 | 0.296 | -0.046 | 0.781 | 0.0288 |
| Starchy roots | 0.015 | 0.929 | 0.195 | 0.234 | 0.0002 |
| Fruit | 0.132 | 0.418 | 0.231 | 0.158 | 0.0174 |
| Other meat | 0.367 | 0.02 | 0.234 | 0.153 | 0.1350 |

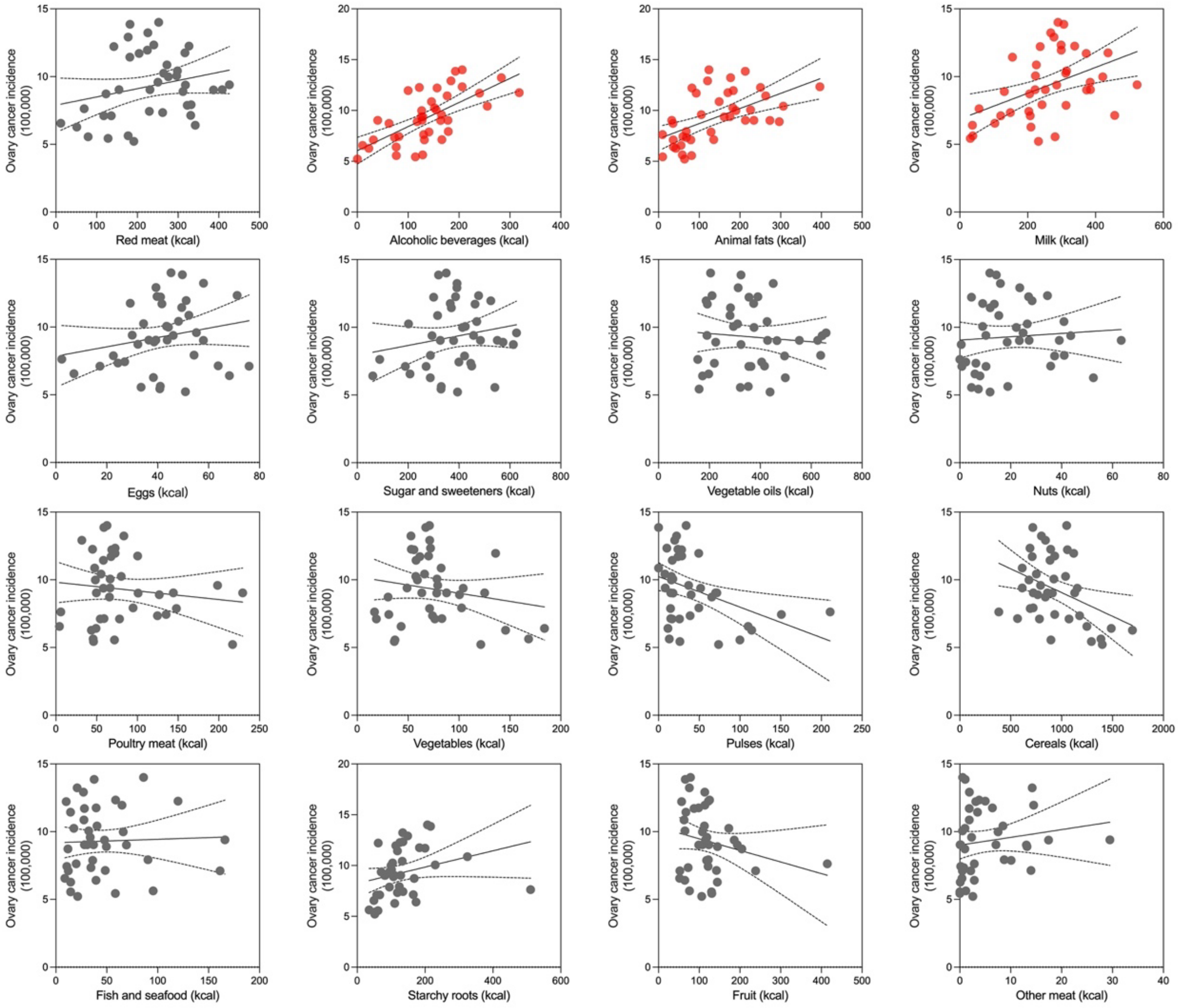


**Figure S13.** Regression analysis of association between 16 dietary factors and ovary cancer incidence.

**Table S12.** Association between dietary factors and ovary cancer incidence.

| Food Type (n=40) | Correlation Coefficient | | | | Linear Regression |
|---|---|---|---|---|---|
| | **CC** | **p-value** | **Partial CC** | **p-value** | **R squared** |
| Red meat | 0.242 | 0.132 | 0.218 | 0.182 | 0.0586 |
| **Alcoholic beverages*** | **0.673** | **0.000** | **0.670** | **0.000** | **0.4529** |
| **Animal fats*** | **0.558** | **0.000** | **0.575** | **0.000** | **0.3116** |
| **Milk*** | **0.450** | **0.004** | **0.473** | **0.002** | **0.2029** |
| Eggs | 0.215 | 0.182 | 0.190 | 0.247 | 0.0464 |
| Sugar and sweeteners | 0.184 | 0.256 | 0.152 | 0.357 | 0.0339 |
| Vegetable oils | -0.087 | 0.594 | -0.16 | 0.331 | 0.0076 |
| Nuts | 0.076 | 0.641 | 0.020 | 0.903 | 0.0058 |
| Poultry meat | -0.130 | 0.426 | -0.189 | 0.249 | 0.0168 |
| Vegetables | -0.182 | 0.262 | -0.203 | 0.215 | 0.0330 |
| **Pulses*** | **-0.396** | **0.011** | **-0.383** | **0.016** | **0.1568** |
| **Cereals*** | **-0.388** | **0.013** | **-0.377** | **0.018** | **0.1502** |
| Fish and seafood | 0.038 | 0.816 | -0.010 | 0.952 | 0.0014 |
| Starchy roots | 0.274 | 0.087 | 0.324 | 0.044 | 0.0751 |
| Fruit | -0.222 | 0.169 | -0.210 | 0.200 | 0.0491 |
| Other meat | 0.147 | 0.365 | 0.115 | 0.486 | 0.0216 |

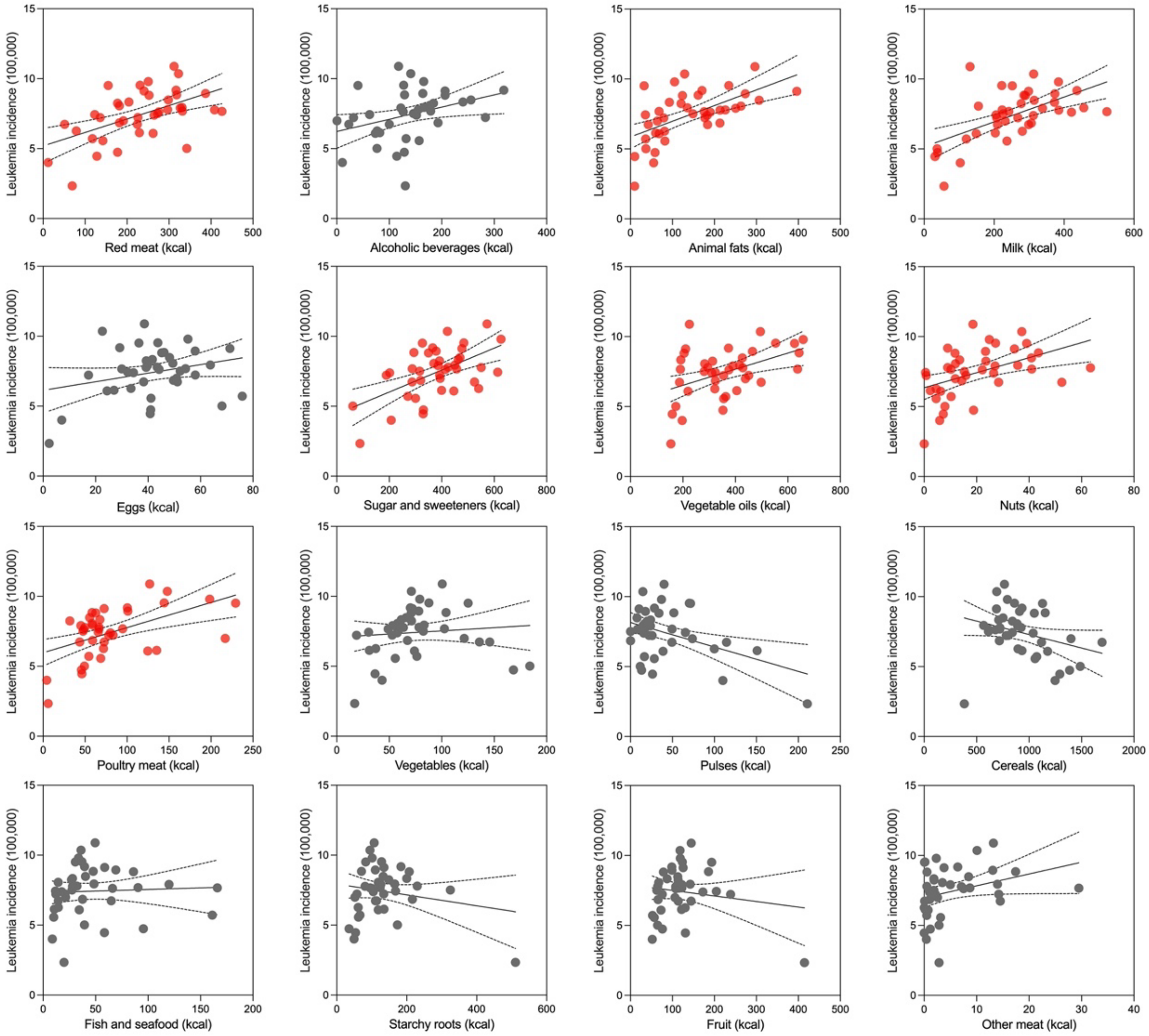


**Figure S14.** Regression analysis of association between 16 dietary factors and Leukemia incidence.

**Table S13.** Association between dietary factors and leukemia cancer incidence.

| Food Type (n=40) | Correlation Coefficient | | | | Linear Regression |
|---|---|---|---|---|---|
| | CC | p-value | Partial CC | p-value | R squared |
| Alcoholic beverages | 0.346 | 0.029 | 0.314 | 0.052 | 0.1198 |
| Red meat | 0.535 | 0.000 | 0.255 | 0.118 | 0.2867 |
| **Animal fats*** | **0.587** | **0.000** | **0.445** | **0.005** | **0.3445** |
| **Milk*** | **0.585** | **0.000** | **0.391** | **0.014** | **0.3425** |
| Eggs | 0.267 | 0.096 | 0.099 | 0.549 | 0.0712 |
| **Sugar and sweeteners*** | **0.561** | **0.000** | **0.434** | **0.006** | **0.3152** |
| Vegetable oils | 0.456 | 0.003 | 0.262 | 0.107 | 0.2078 |
| Nuts | 0.462 | 0.003 | 0.239 | 0.143 | 0.2139 |
| **Poultry meat*** | **0.509** | **0.001** | **0.381** | **0.017** | **0.2589** |
| Vegetables | 0.096 | 0.557 | 0.009 | 0.956 | 0.0092 |
| Pulses | -0.429 | 0.006 | -0.299 | 0.064 | 0.1839 |
| Cereals | -0.297 | 0.062 | -0.253 | 0.120 | 0.0884 |
| Fish and seafood | 0.049 | 0.766 | -0.247 | 0.130 | 0.0024 |
| Starchy roots | -0.185 | 0.253 | -0.022 | 0.893 | 0.0343 |
| Fruit | -0.144 | 0.376 | -0.082 | 0.620 | 0.0206 |
| Other meat | 0.302 | 0.058 | 0.129 | 0.434 | 0.0912 |

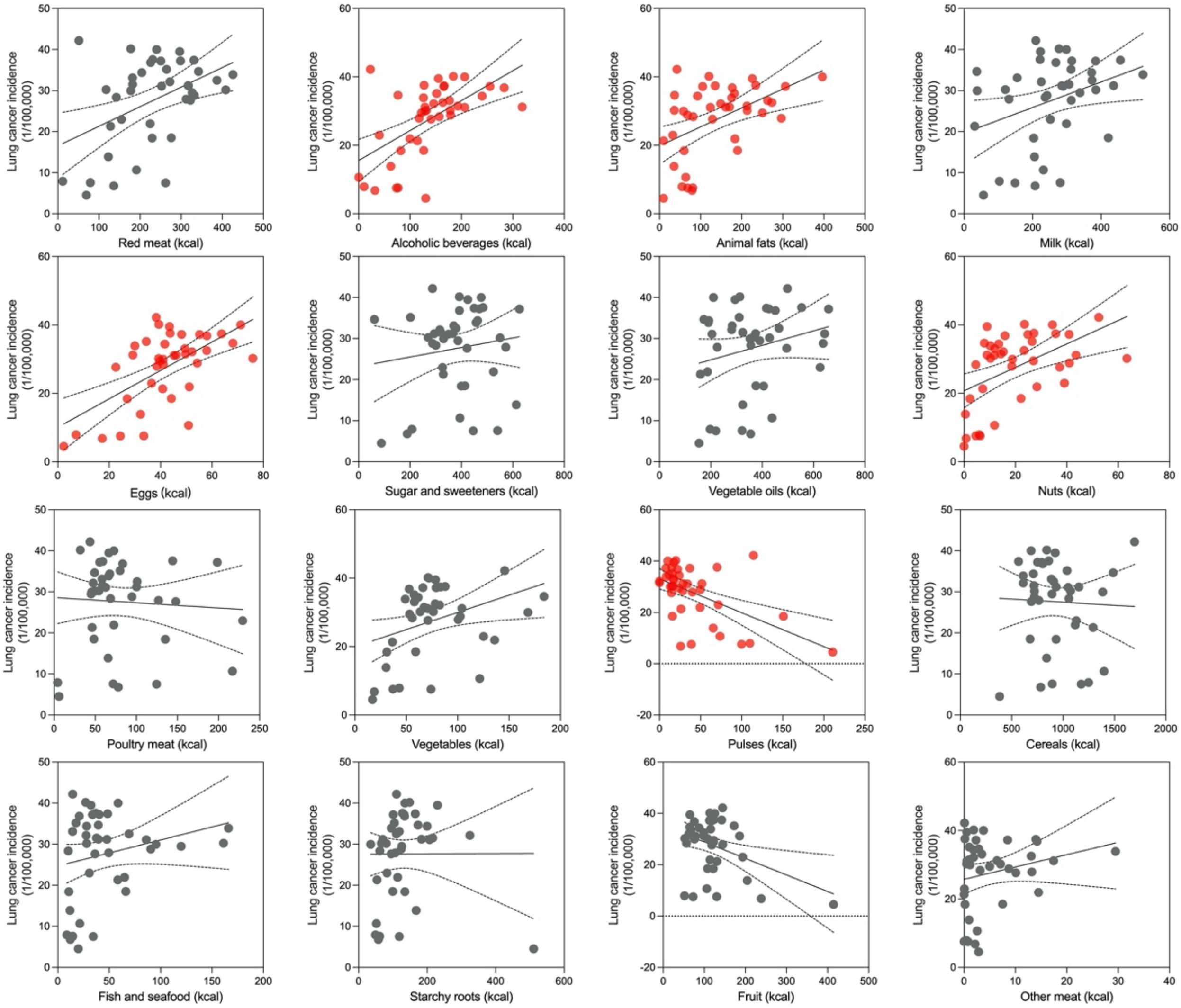


**Figure S15.** Regression analysis of association between 16 dietary factors and lung cancer incidence.

**Table S14.** Association between dietary factors and lung cancer incidence.

| Food Type (n=40) | Correlation Coefficient | | | | Linear Regression |
|---|---|---|---|---|---|
| | CC | p-value | Partial CC | p-value | R squared |
| **Red meat*** | **0.445** | **0.004** | **0.334** | **0.037** | **0.1981** |
| **Alcoholic beverages*** | **0.582** | **0.000** | **0.546** | **0.000** | **0.3383** |
| **Animal fats*** | **0.488** | **0.001** | **0.403** | **0.011** | **0.2380** |
| Milk | 0.344 | 0.03 | 0.206 | 0.209 | 0.1184 |
| **Eggs*** | **0.608** | **0.000** | **0.56** | **0.000** | **0.3696** |
| Sugar and sweeteners | 0.139 | 0.393 | 0.003 | 0.983 | 0.0193 |
| Vegetable oils | 0.234 | 0.145 | 0.098 | 0.554 | 0.0549 |
| **Nuts*** | **0.495** | **0.001** | **0.404** | **0.011** | **0.2452** |
| Poultry meat | -0.061 | 0.711 | -0.214 | 0.192 | 0.0037 |
| **Vegetables*** | **0.354** | **0.025** | **0.325** | **0.044** | **0.1255** |
| **Pulses*** | **-0.544** | **0.000** | **-0.485** | **0.002** | **0.2957** |
| Cereals | -0.04 | 0.809 | 0.014 | 0.934 | 0.0016 |
| Fish and seafood | 0.225 | 0.162 | 0.105 | 0.526 | 0.0508 |
| Starchy roots | 0.004 | 0.983 | 0.111 | 0.501 | 0.0000 |
| **Fruit*** | **-0.391** | **0.013** | **-0.37** | **0.020** | **0.1530** |
| Other meat | 0.216 | 0.180 | 0.113 | 0.494 | 0.0468 |

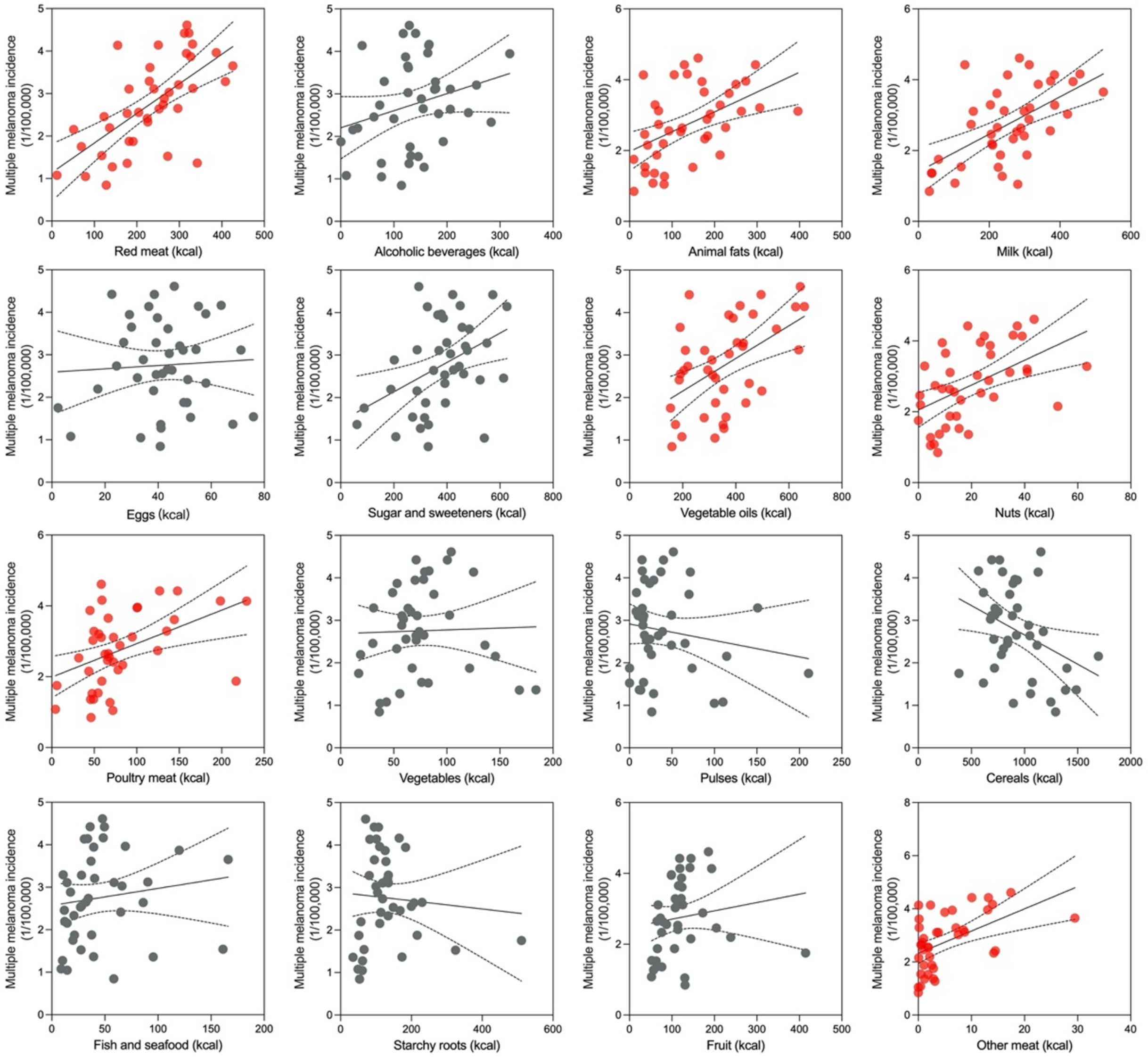


**Figure S16.** Regression analysis of association between 16 dietary factors and multiple melanoma incidence.

**Table S15.** Association between dietary factors and multiple melanoma incidence.

| Food Type (n=40) | Correlation Coefficient | | | | Linear Regression |
|---|---|---|---|---|---|
| | CC | p-value | Partial CC | p-value | R squared |
| **Red meat*** | **0.649** | **0.000** | **0.452** | **0.004** | **0.4210** |
| Alcoholic beverages | 0.267 | 0.096 | 0.143 | 0.384 | 0.0711 |
| Animal fats | 0.491 | 0.001 | 0.290 | 0.073 | 0.2413 |
| **Milk*** | **0.584** | **0.000** | **0.355** | **0.027** | **0.3405** |
| Eggs | 0.057 | 0.729 | -0.212 | 0.196 | 0.0032 |
| Sugar and sweeteners | 0.414 | 0.008 | 0.213 | 0.192 | 0.1713 |
| Vegetable oils | 0.510 | 0.001 | 0.312 | 0.053 | 0.2600 |
| Nuts | 0.506 | 0.001 | 0.267 | 0.100 | 0.2558 |
| Poultry meat | 0.449 | 0.004 | 0.286 | 0.077 | 0.2013 |
| Vegetables | 0.030 | 0.853 | -0.088 | 0.592 | 0.0009 |
| Pulses | -0.159 | 0.326 | 0.083 | 0.616 | 0.0254 |
| **Cereals*** | **-0.357** | **0.024** | **-0.332** | **0.039** | **0.1273** |
| Fish and seafood | 0.142 | 0.381 | -0.168 | 0.307 | 0.0203 |
| Starchy roots | -0.077 | 0.636 | 0.147 | 0.371 | 0.0060 |
| Fruit | 0.142 | 0.381 | 0.294 | 0.069 | 0.0203 |
| **Other meat*** | **0.502** | **0.001** | **0.379** | **0.017** | **0.2518** |

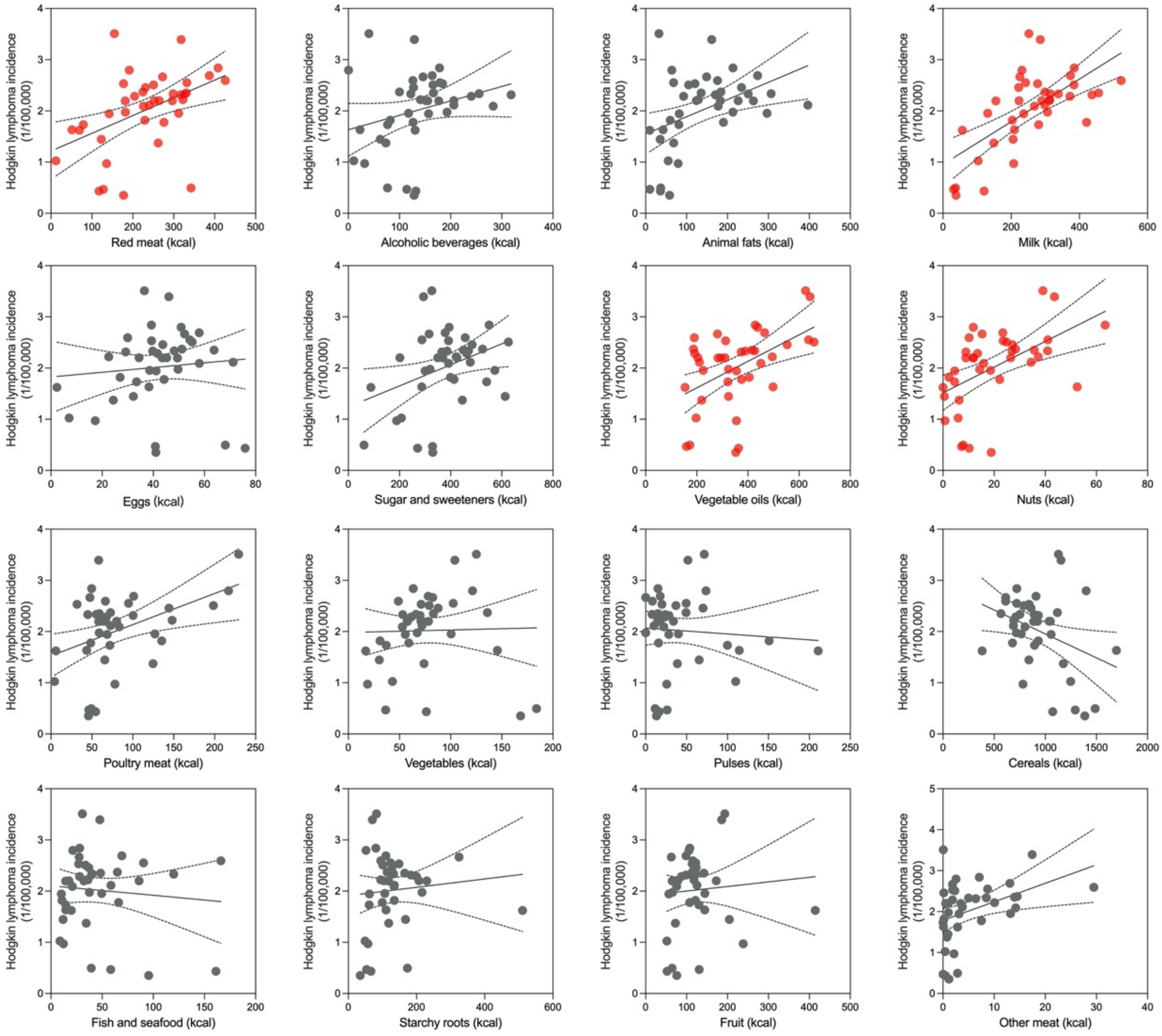


**Figure S17.** Regression analysis of association between 16 dietary factors and Hodgkin lymphoma incidence.

**Table S16.** Association between dietary factors and Hodgkin lymphoma incidence.

| **Food Type (n=40)** | **Correlation Coefficient** | | | | **Linear Regression** |
|---|---|---|---|---|---|
| | **CC** | **p-value** | **Partial CC** | **p-value** | **R squared** |
| Red meat | 0.46 | 0.003 | 0.266 | 0.102 | 0.2113 |
| Alcoholic beverages | 0.266 | 0.097 | 0.173 | 0.291 | 0.0708 |
| Animal fats | 0.421 | 0.007 | 0.262 | 0.108 | 0.1769 |
| **Milk*** | **0.655** | **0.000** | **0.537** | **0.000** | **0.4287** |
| Eggs | 0.096 | 0.557 | -0.073 | 0.657 | 0.0092 |
| Sugar and sweeteners | 0.350 | 0.027 | 0.193 | 0.238 | 0.1225 |
| **Vegetable oils*** | **0.489** | **0.001** | **0.345** | **0.031** | **0.2388** |
| **Nuts*** | **0.515** | **0.001** | **0.360** | **0.024** | **0.2651** |
| Poultry meat | 0.411 | 0.008 | 0.283 | 0.081 | 0.1689 |
| Vegetables | 0.024 | 0.881 | -0.055 | 0.738 | 0.0006 |
| Pulses | -0.067 | 0.682 | 0.115 | 0.484 | 0.0045 |
| Cereals | -0.344 | 0.030 | -0.308 | 0.056 | 0.1185 |
| Fish and seafood | -0.094 | 0.565 | -0.357 | 0.026 | 0.0088 |
| Starchy roots | 0.093 | 0.570 | 0.273 | 0.093 | 0.0086 |
| Fruit | 0.077 | 0.636 | 0.160 | 0.332 | 0.0059 |
| Other meat | 0.386 | 0.014 | 0.264 | 0.104 | 0.1487 |

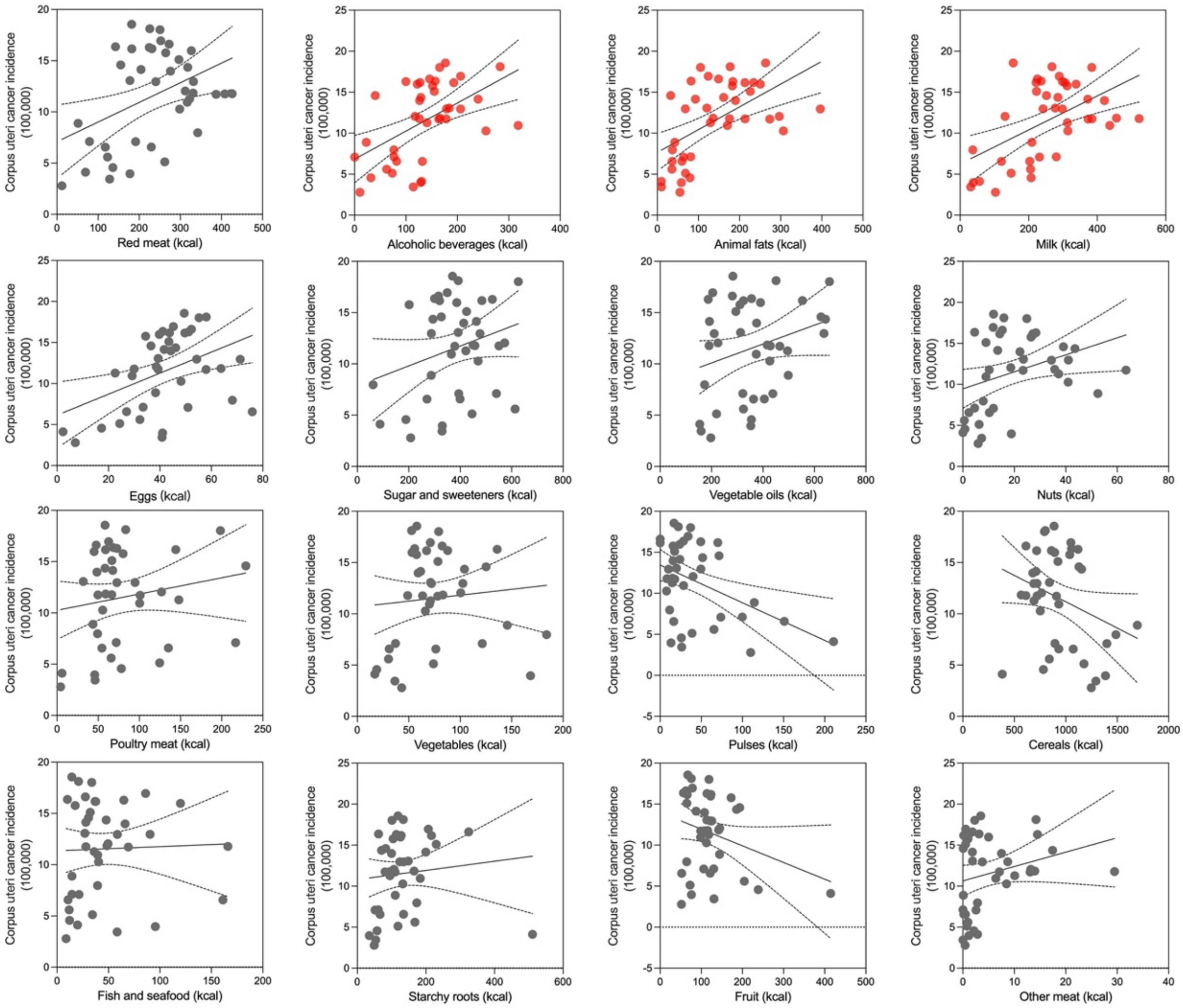


**Figure S18.** Regression analysis of association between 16 dietary factors and corpus uteri cancer incidence.

**Table S17.** Association between dietary factors and corpus uteri incidence.

| Food Type (n=40) | Correlation Coefficient | | | | Linear Regression |
|---|---|---|---|---|---|
| | CC | p-value | Partial CC | p-value | R squared |
| Red meat | 0.405 | 0.010 | -0.255 | 0.117 | 0.1637 |
| Alcoholic beverages | 0.515 | 0.001 | 0.124 | 0.452 | 0.2653 |
| Animal fats | 0.552 | 0.000 | -0.107 | 0.515 | 0.3049 |
| Milk | 0.519 | 0.001 | -0.118 | 0.474 | 0.2691 |
| **Eggs*** | **0.424** | **0.006** | **-0.514** | **0.001** | **0.1800** |
| Sugar and sweeteners | 0.262 | 0.103 | -0.123 | 0.454 | 0.0686 |
| Vegetable oils | 0.277 | 0.084 | -0.268 | 0.099 | 0.0767 |
| **Nuts*** | **0.338** | **0.033** | **-0.445** | **0.005** | **0.1144** |
| Poultry meat | 0.170 | 0.294 | -0.175 | 0.287 | 0.0289 |
| Vegetables | 0.092 | 0.573 | -0.624 | 0.000 | 0.0084 |
| **Pulses*** | **-0.427** | **0.006** | **0.501** | **0.001** | **0.1826** |
| Cereals | -0.301 | 0.059 | -0.404 | 0.011 | 0.0906 |
| Fish and seafood | 0.033 | 0.841 | -0.020 | 0.905 | 0.0011 |
| Starchy roots | 0.101 | 0.535 | 0.493 | 0.001 | 0.0102 |
| Fruit | -0.279 | 0.081 | 0.525 | 0.001 | 0.0780 |
| Other meat | 0.238 | 0.139 | -0.152 | 0.356 | 0.0566 |

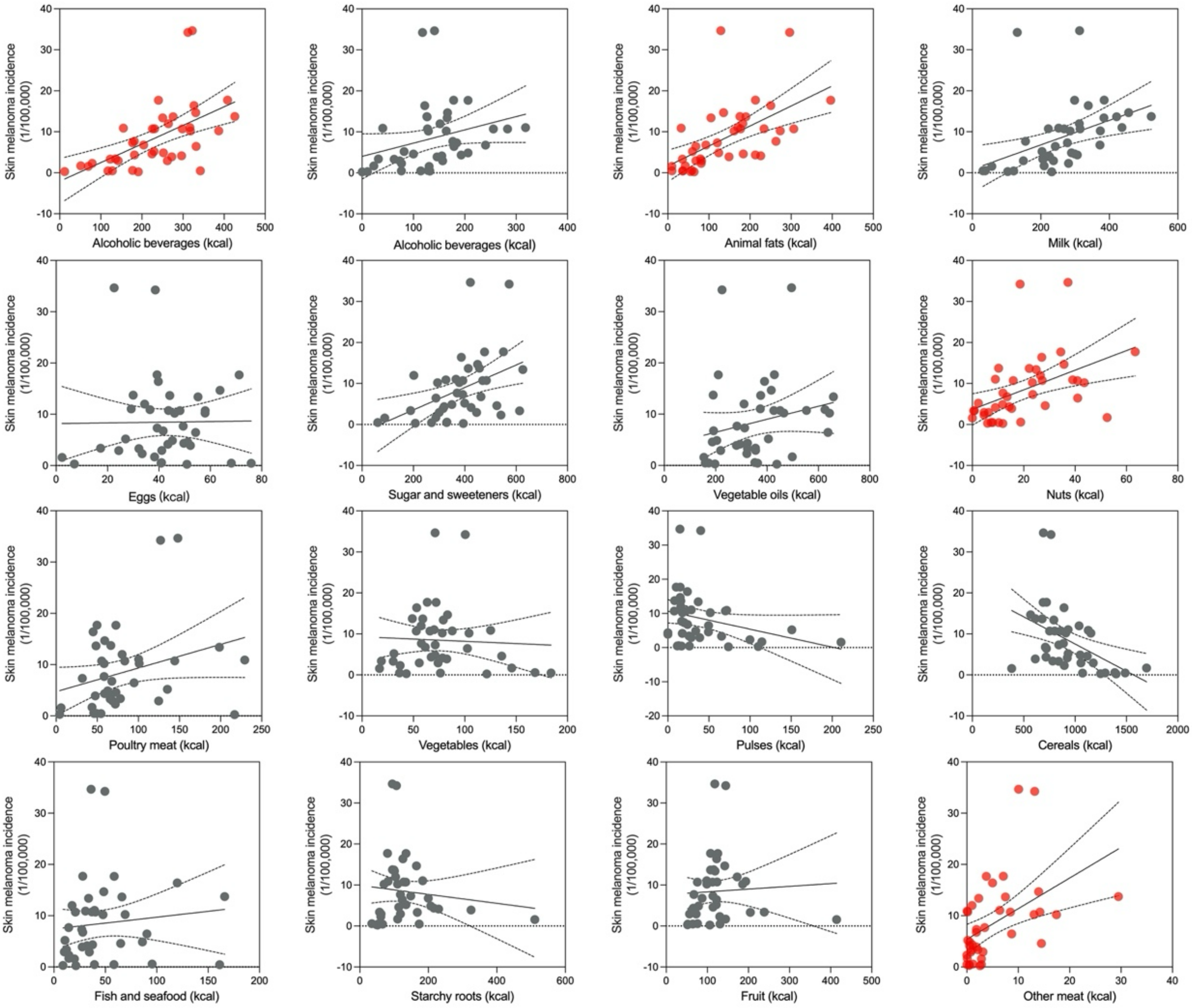


**Figure S19.** Regression analysis of association between 16 dietary factors and skin melanoma Incidence.

**Table S18.** Association between dietary factors and skin melanoma incidence.

| Food Type (n=40) | Correlation Coefficient | | | | Linear Regression |
|---|---|---|---|---|---|
| | CC | p-value | Partial CC | p-value | R squared |
| **Red meat*** | **0.562** | **0.000** | **0.357** | **0.026** | **0.3158** |
| Alcoholic beverages | 0.286 | 0.074 | 0.180 | 0.272 | 0.0815 |
| **Animal fats*** | **0.570** | **0.000** | **0.424** | **0.007** | **0.3250** |
| Milk | 0.439 | 0.005 | 0.181 | 0.269 | 0.1925 |
| Eggs | 0.013 | 0.939 | -0.223 | 0.173 | 0.0002 |
| Sugar and sweeteners | 0.435 | 0.005 | 0.269 | 0.098 | 0.1895 |
| Vegetable oils | 0.223 | 0.166 | -0.052 | 0.755 | 0.0499 |
| Nuts | 0.459 | 0.003 | 0.238 | 0.145 | 0.2105 |
| Poultry meat | 0.290 | 0.069 | 0.098 | 0.552 | 0.0843 |
| Vegetables | -0.052 | 0.752 | -0.168 | 0.305 | 0.0027 |
| Pulses | -0.287 | 0.073 | -0.119 | 0.472 | 0.0822 |
| **Cereals*** | **-0.456** | **0.003** | **-0.446** | **0.004** | **0.2079** |
| Fish and seafood | 0.108 | 0.507 | -0.163 | 0.323 | 0.0117 |
| Starchy roots | -0.116 | 0.475 | 0.062 | 0.709 | 0.0135 |
| Fruit | 0.054 | 0.741 | 0.157 | 0.341 | 0.0029 |
| **Other meat*** | **0.477** | **0.002** | **0.356** | **0.026** | **0.2271** |

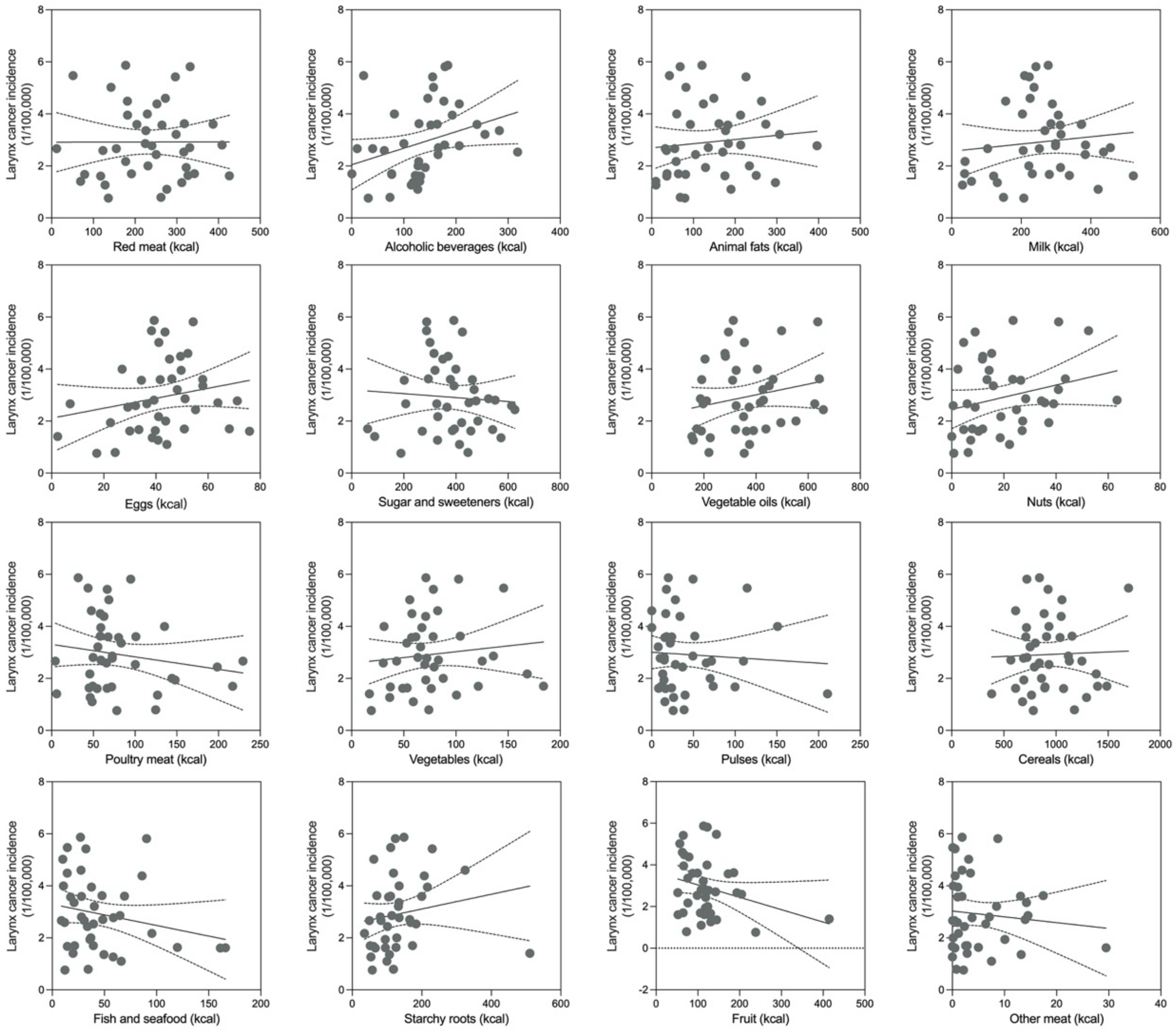


**Figure S20.** Regression analysis of association between 16 dietary factors and larynx cancer incidence.

**Table S19.** Association between dietary factors and larynx incidence.

| Food Type (n=40) | Correlation Coefficient | | | | Linear Regression |
|---|---|---|---|---|---|
| | **CC** | **p-value** | **Partial CC** | **p-value** | **R squared** |
| Red meat | 0.002 | 0.992 | 0.165 | 0.316 | 0.0000 |
| Alcoholic beverages | 0.315 | 0.048 | 0.395 | 0.013 | 0.0993 |
| Animal fats | 0.106 | 0.517 | 0.243 | 0.136 | 0.0111 |
| Milk | 0.115 | 0.481 | 0.302 | 0.061 | 0.0131 |
| Eggs | 0.209 | 0.195 | 0.311 | 0.054 | 0.0439 |
| Sugar and sweeteners | -0.067 | 0.679 | 0.030 | 0.854 | 0.0045 |
| Vegetable oils | 0.204 | 0.206 | 0.362 | 0.024 | 0.0418 |
| Nuts | 0.252 | 0.117 | 0.445 | 0.004 | 0.0634 |
| Poultry meat | -0.170 | 0.295 | -0.092 | 0.578 | 0.0288 |
| Vegetables | 0.117 | 0.471 | 0.159 | 0.335 | 0.0137 |
| Pulses | -0.064 | 0.695 | -0.157 | 0.341 | 0.0041 |
| Cereals | 0.033 | 0.840 | -0.003 | 0.984 | 0.0011 |
| Fish and seafood | -0.216 | 0.181 | -0.138 | 0.403 | 0.0465 |
| Starchy roots | 0.169 | 0.297 | 0.110 | 0.507 | 0.0286 |
| Fruit | -0.269 | 0.093 | -0.310 | 0.055 | 0.0723 |
| Other meat | -0.101 | 0.537 | -0.021 | 0.897 | 0.0101 |

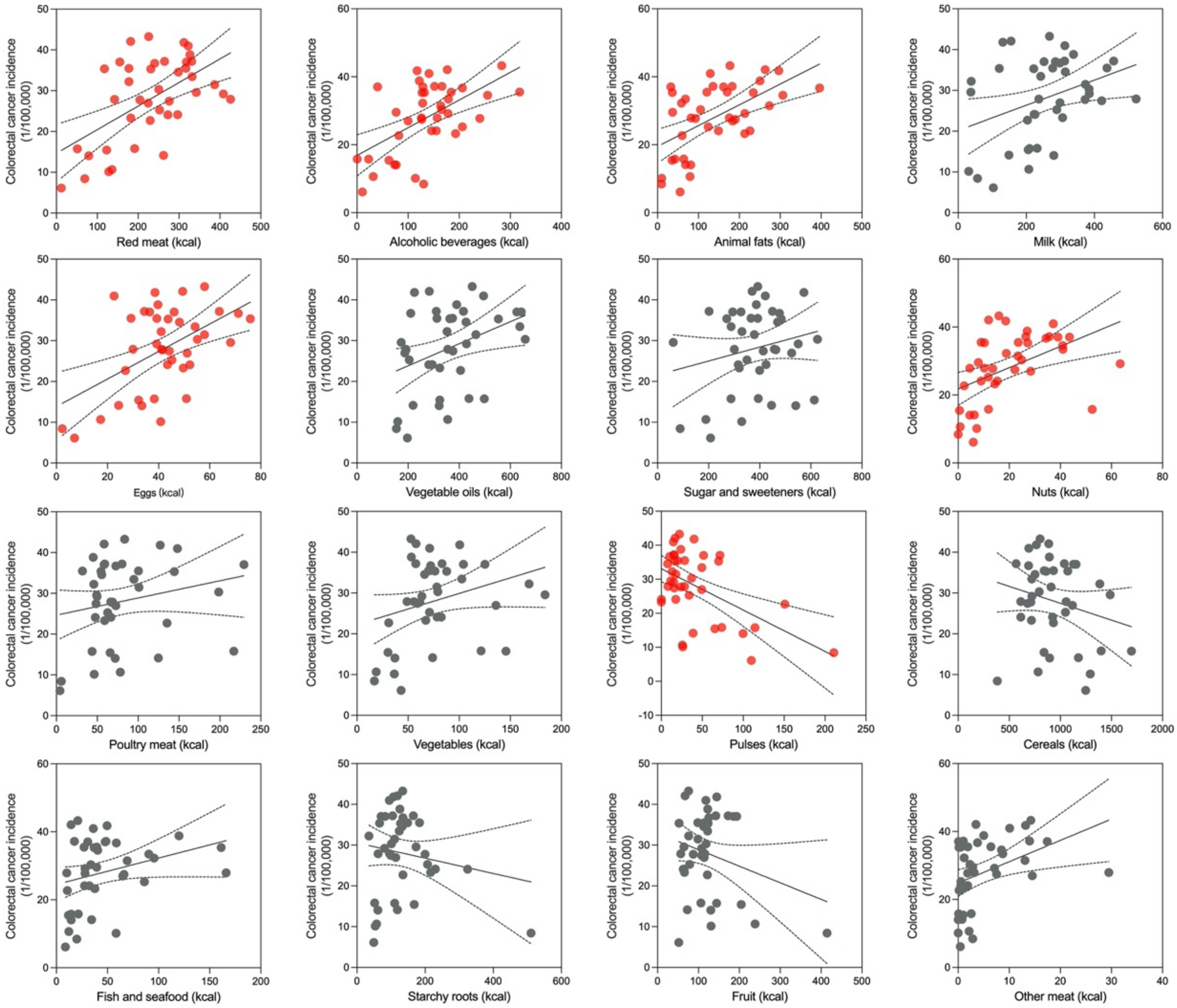


**Figure S21.** Regression analysis of association between 16 dietary factors and colorectal cancer incidence.

**Table S20.** Association between dietary factors and colorectal cancer incidence.

| **Food Type (n=40)** | **Correlation Coefficient** | | | | **Linear Regression** |
|---|---|---|---|---|---|
| | **CC** | **p-value** | **Partial CC** | **p-value** | **R squared** |
| **Red meat*** | **0.56** | **0.000** | **0.357** | **0.026** | **0.3135** |
| **Alcoholic beverages*** | **0.562** | **0.000** | **0.523** | **0.001** | **0.3162** |
| **Animal fats*** | **0.558** | **0.000** | **0.410** | **0.010** | **0.3116** |
| Milk | 0.355 | 0.025 | 0.062 | 0.706 | 0.1258 |
| **Eggs*** | **0.513** | **0.001** | **0.415** | **0.009** | **0.2627** |
| Sugar and sweeteners | 0.212 | 0.188 | -0.023 | 0.888 | 0.0451 |
| Vegetable oils | 0.373 | 0.018 | 0.154 | 0.348 | 0.1390 |
| Nuts | 0.469 | 0.002 | 0.255 | 0.118 | 0.2198 |
| Poultry meat | 0.208 | 0.199 | -0.007 | 0.967 | 0.0431 |
| Vegetables | 0.279 | 0.081 | 0.234 | 0.152 | 0.0780 |
| **Pulses*** | **-0.520** | **0.001** | **-0.418** | **0.008** | **0.2701** |
| Cereals | -0.223 | 0.167 | -0.163 | 0.323 | 0.0496 |
| Fish and seafood | 0.286 | 0.073 | 0.075 | 0.650 | 0.0819 |
| Starchy roots | -0.156 | 0.337 | 0.010 | 0.950 | 0.0242 |
| Fruit | -0.256 | 0.111 | -0.218 | 0.181 | 0.0654 |
| Other meat | 0.391 | 0.013 | 0.248 | 0.129 | 0.1529 |

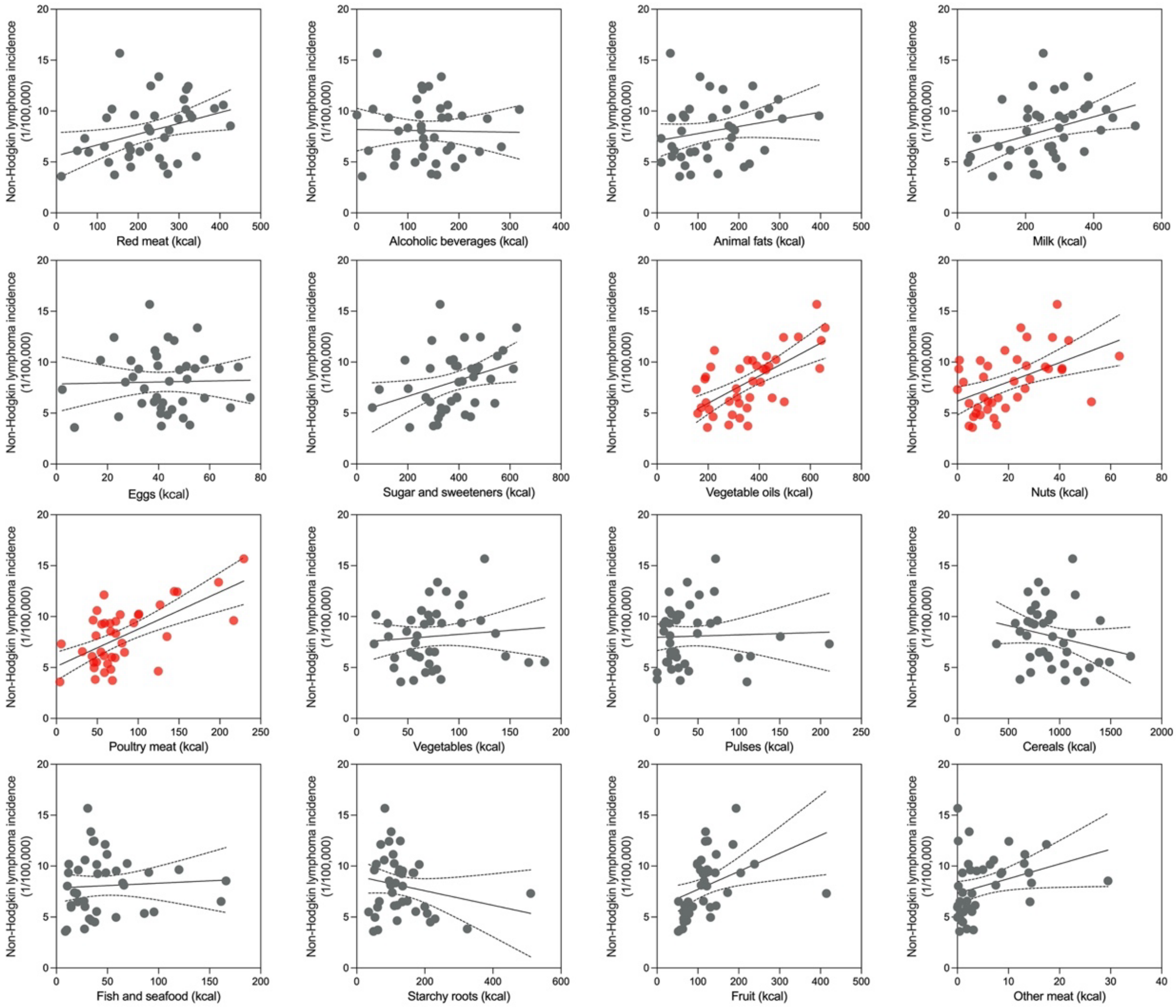


**Figure S22.** Regression analysis of association between 16 dietary factors and non-Hodgkin lymphoma incidence.

**Table S21.** Association between dietary factors and non-Hodgkin lymphoma incidence.

| Food Type (n=40) | Correlation Coefficient | | | | Linear Regression |
|---|---|---|---|---|---|
| | CC | p-value | Partial CC | p-value | R squared |
| Red meat | 0.36 | 0.022 | 0.008 | 0.963 | 0.1299 |
| Alcoholic beverages | -0.021 | 0.898 | -0.232 | 0.155 | 0.0004 |
| Animal fats | 0.224 | 0.164 | -0.089 | 0.590 | 0.0503 |
| Milk | 0.381 | 0.015 | 0.047 | 0.777 | 0.1448 |
| Eggs | 0.026 | 0.874 | -0.247 | 0.130 | 0.0007 |
| Sugar and sweeteners | 0.345 | 0.029 | 0.121 | 0.465 | 0.1189 |
| **Vegetable oils*** | **0.647** | **0.000** | **0.513** | **0.001** | **0.4192** |
| Nuts | 0.496 | 0.001 | 0.257 | 0.114 | 0.2456 |
| **Poultry meat*** | **0.632** | **0.000** | **0.541** | **0.000** | **0.3999** |
| Vegetables | 0.101 | 0.535 | 0.005 | 0.975 | 0.0102 |
| **Pulses*** | **0.036** | **0.825** | **0.342** | **0.033** | **0.0013** |
| Cereals | -0.227 | 0.158 | -0.164 | 0.317 | 0.0517 |
| Fish and seafood | 0.061 | 0.709 | -0.275 | 0.091 | 0.0037 |
| Starchy roots | -0.208 | 0.197 | -0.032 | 0.847 | 0.0433 |
| **Fruit*** | **0.392** | **0.012** | **0.609** | **0.000** | **0.1534** |
| Other meat | 0.315 | 0.047 | 0.127 | 0.442 | 0.0995 |

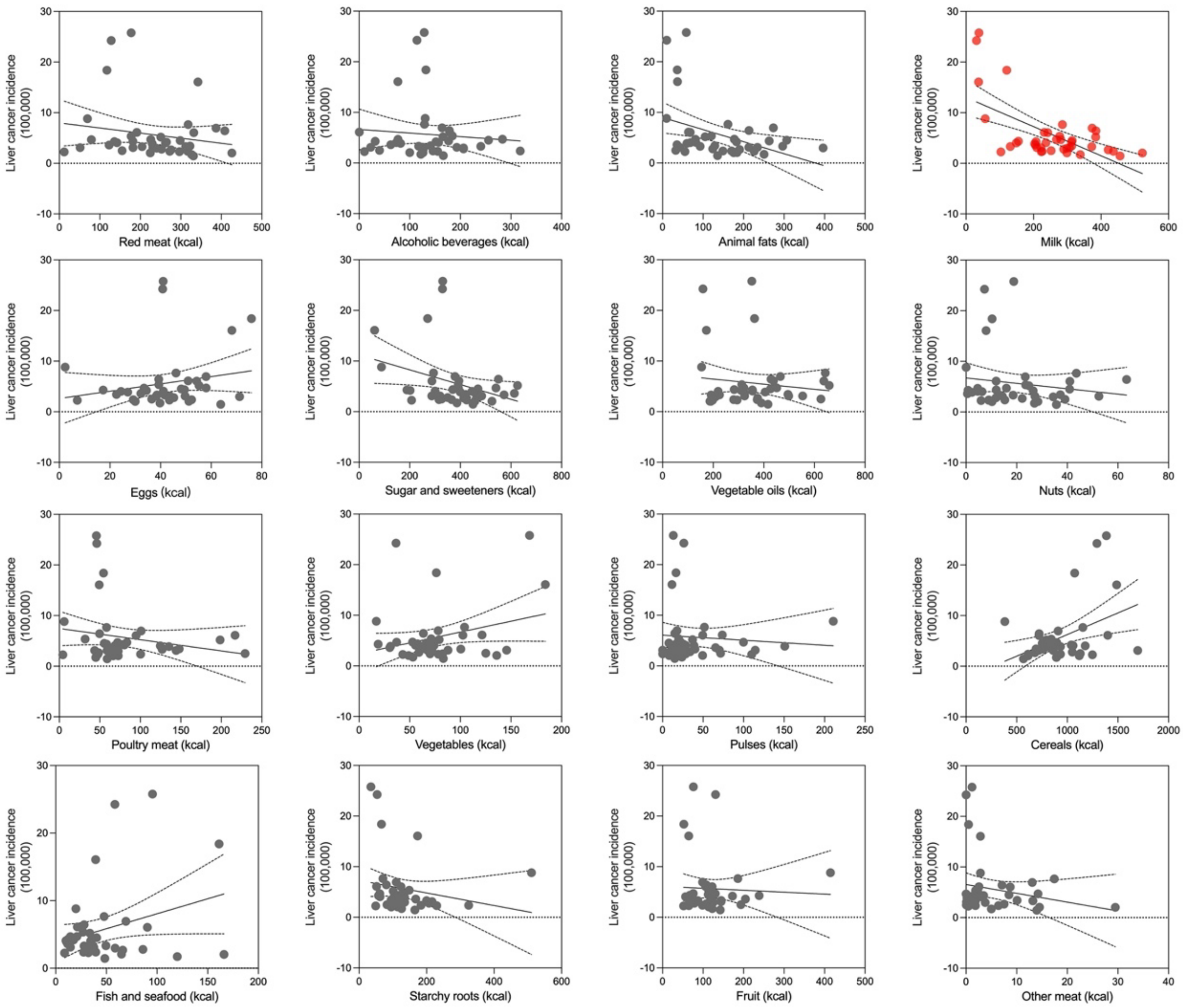


**Figure S23.** Regression analysis of association between 16 dietary factors and liver cancer incidence.

**Table S22.** Association between dietary factors and Liver cancer incidence.

| Food Type (n=40) | Correlation Coefficient | | | | Linear Regression |
|---|---|---|---|---|---|
| | CC | p-value | Partial CC | p-value | R squared |
| Red meat | -0.176 | 0.276 | -0.126 | 0.445 | 0.0311 |
| Alcoholic beverages | -0.09 | 0.583 | -0.059 | 0.720 | 0.0080 |
| **Animal fats*** | **-0.395** | **0.012** | **-0.381** | **0.017** | **0.1559** |
| **Milk*** | **-0.600** | **0.000** | **-0.646** | **0.000** | **0.3603** |
| Eggs | 0.199 | 0.219 | 0.260 | 0.110 | 0.0395 |
| Sugar and sweeteners | -0.329 | 0.038 | -0.305 | 0.059 | 0.1079 |
| Vegetable oils | -0.126 | 0.438 | -0.075 | 0.651 | 0.0159 |
| Nuts | -0.145 | 0.372 | -0.092 | 0.579 | 0.0211 |
| Poultry meat | -0.198 | 0.220 | -0.162 | 0.325 | 0.0393 |
| Vegetables | 0.284 | 0.076 | 0.310 | 0.054 | 0.0805 |
| Pulses | -0.077 | 0.636 | -0.132 | 0.423 | 0.0060 |
| **Cereals*** | **0.417** | **0.007** | **0.405** | **0.011** | **0.1737** |
| Fish and seafood | 0.297 | 0.063 | 0.389 | 0.014 | 0.0881 |
| Starchy roots | -0.187 | 0.249 | -0.238 | 0.144 | 0.0349 |
| Fruit | -0.044 | 0.789 | -0.062 | 0.706 | 0.0019 |
| Other meat | -0.197 | 0.223 | -0.163 | 0.322 | 0.0389 |

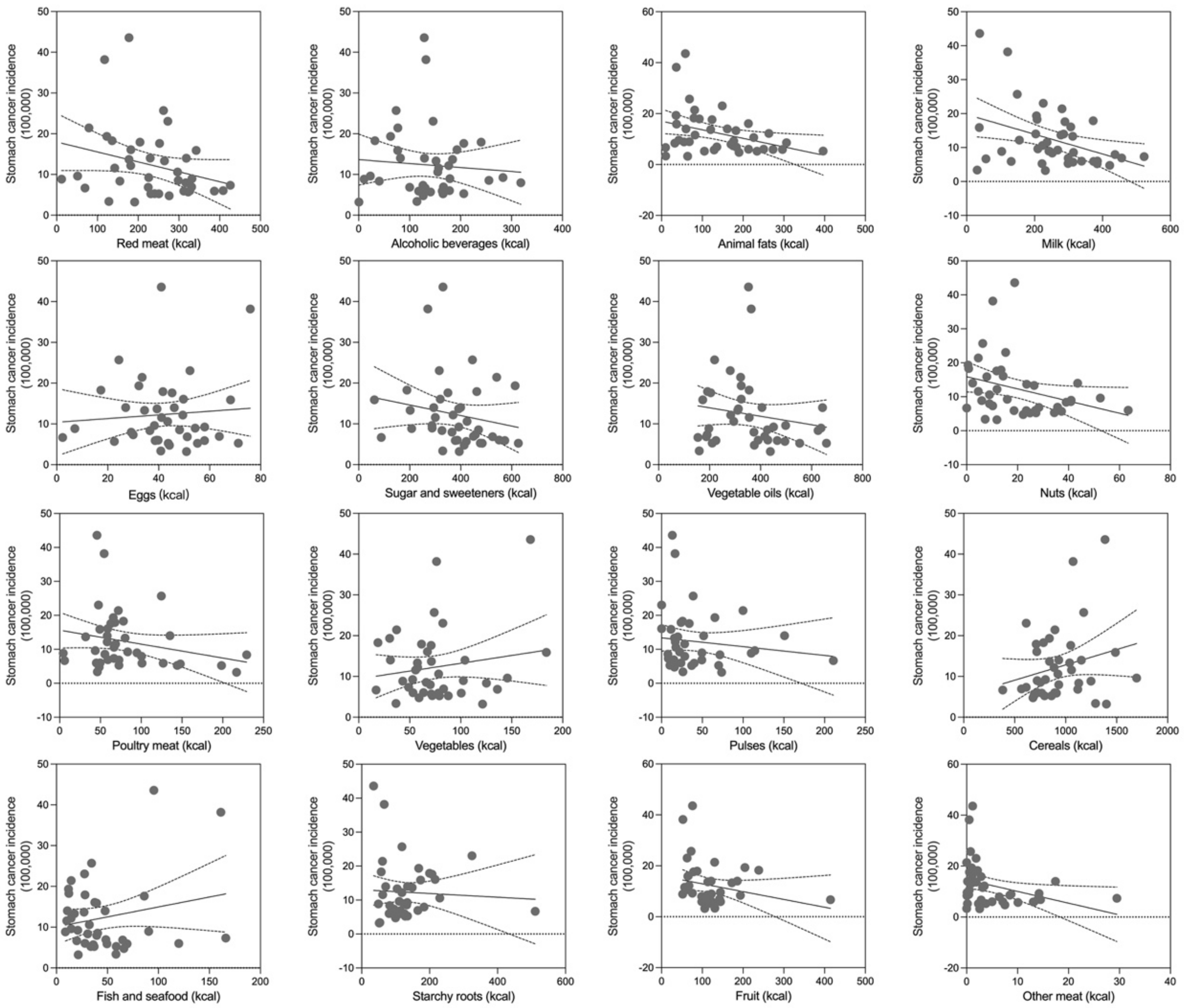


**Figure S24.** Regression analysis of association between 16 dietary factors and stomach cancer incidence.

**Table S23.** Association between dietary factors and stomach cancer incidence.

| Food Type (n=40) | Correlation Coefficient | | | | Linear Regression |
|---|---|---|---|---|---|
| | CC | p-value | Partial CC | p-value | R squared |
| Red meat | -0.275 | 0.085 | -0.111 | 0.501 | 0.0759 |
| Alcoholic beverages | -0.078 | 0.631 | 0.005 | 0.974 | 0.0061 |
| Animal fats | -0.358 | 0.023 | -0.245 | 0.133 | 0.1279 |
| Milk | -0.390 | 0.013 | -0.261 | 0.109 | 0.1518 |
| Eggs | 0.080 | 0.623 | 0.215 | 0.188 | 0.0064 |
| Sugar and sweeteners | -0.188 | 0.246 | -0.058 | 0.726 | 0.0352 |
| Vegetable oils | -0.167 | 0.303 | -0.014 | 0.932 | 0.0279 |
| Nuts | -0.314 | 0.049 | -0.176 | 0.285 | 0.0984 |
| Poultry meat | -0.236 | 0.143 | -0.124 | 0.451 | 0.0556 |
| Vegetables | 0.163 | 0.314 | 0.231 | 0.157 | 0.0267 |
| Pulses | -0.130 | 0.426 | -0.277 | 0.088 | 0.0168 |
| Cereals | 0.236 | 0.143 | 0.196 | 0.231 | 0.0557 |
| Fish and seafood | 0.210 | 0.194 | 0.405 | 0.011 | 0.0440 |
| Starchy roots | -0.053 | 0.744 | -0.168 | 0.306 | 0.0028 |
| Fruit | -0.225 | 0.162 | -0.289 | 0.075 | 0.0508 |
| Other meat | -0.336 | 0.034 | -0.247 | 0.130 | 0.1128 |

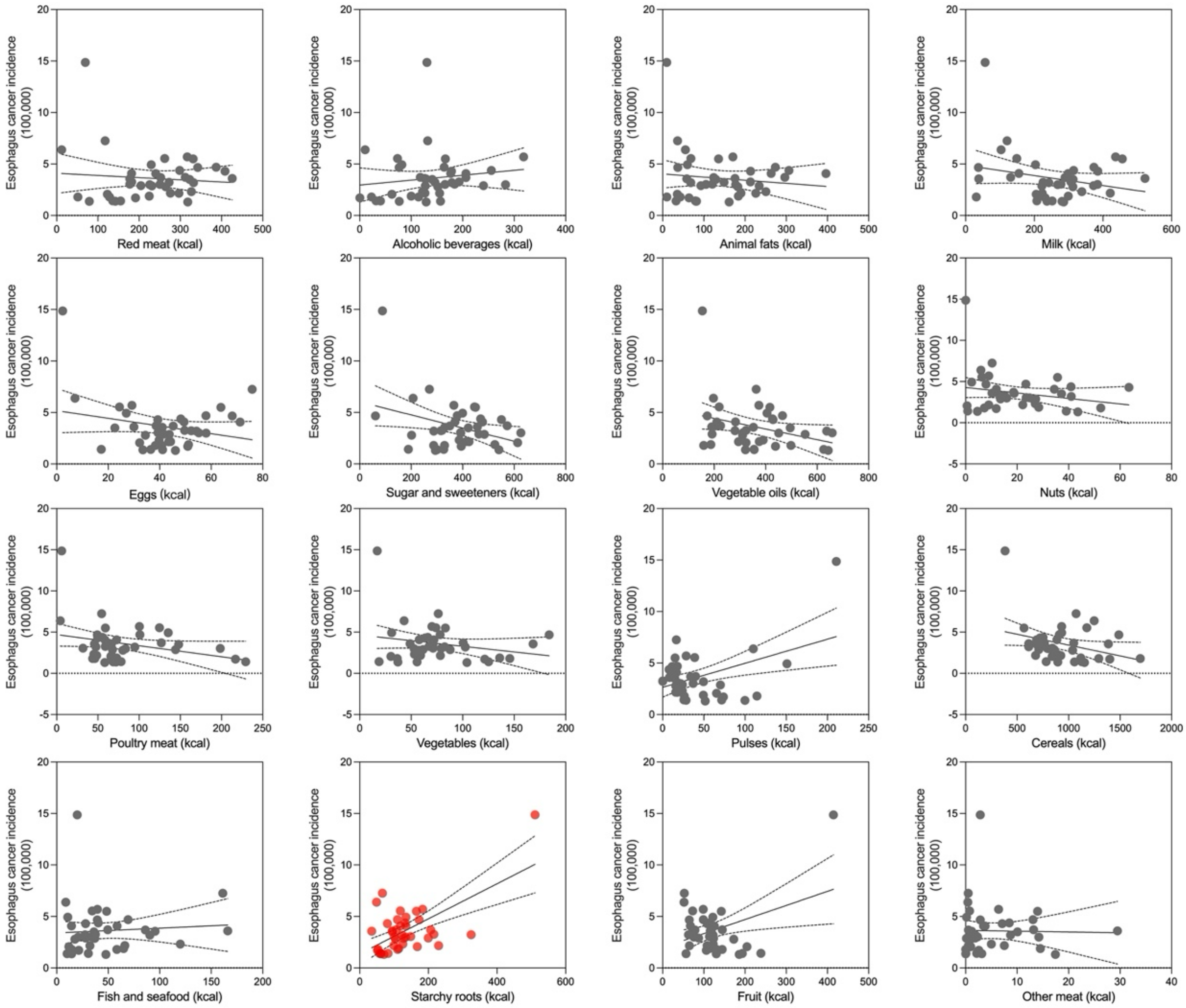


**Figure S25.** Regression analysis of association between 16 dietary factors and esophagus cancer incidence.

**Table S24.** Association between dietary factors and esophagus cancer incidence.

| Food Type (n=40) | Correlation Coefficient | | | | Linear Regression |
|---|---|---|---|---|---|
| | **CC** | **p-value** | **Partial CC** | **p-value** | **R squared** |
| Red meat | -0.089 | 0.583 | -0.005 | 0.975 | 0.0080 |
| Alcoholic beverages | 0.144 | 0.377 | 0.189 | 0.249 | 0.0206 |
| Animal fats | -0.122 | 0.454 | -0.061 | 0.713 | 0.0148 |
| Milk | -0.244 | 0.130 | -0.196 | 0.231 | 0.0594 |
| Eggs | -0.246 | 0.126 | -0.211 | 0.198 | 0.0605 |
| **Sugar and sweeteners*** | **-0.346** | **0.029** | **-0.317** | **0.049** | **0.1200** |
| Vegetable oils | -0.311 | 0.051 | -0.276 | 0.089 | 0.0964 |
| Nuts | -0.218 | 0.177 | -0.167 | 0.310 | 0.0475 |
| Poultry meat | -0.289 | 0.071 | -0.254 | 0.119 | 0.0834 |
| Vegetables | -0.218 | 0.176 | -0.200 | 0.222 | 0.0477 |
| **Pulses*** | **0.435** | **0.005** | **0.413** | **0.009** | **0.1888** |
| Cereals | -0.308 | 0.053 | -0.340 | 0.034 | 0.0947 |
| Fish and seafood | 0.074 | 0.650 | 0.151 | 0.359 | 0.0055 |
| **Starchy roots*** | **0.616** | **0.000** | **0.606** | **0.000** | **0.3790** |
| **Fruit*** | **0.375** | **0.017** | **0.362** | **0.024** | **0.1405** |
| Other meat | -0.021 | 0.897 | 0.035 | 0.833 | 0.0004 |

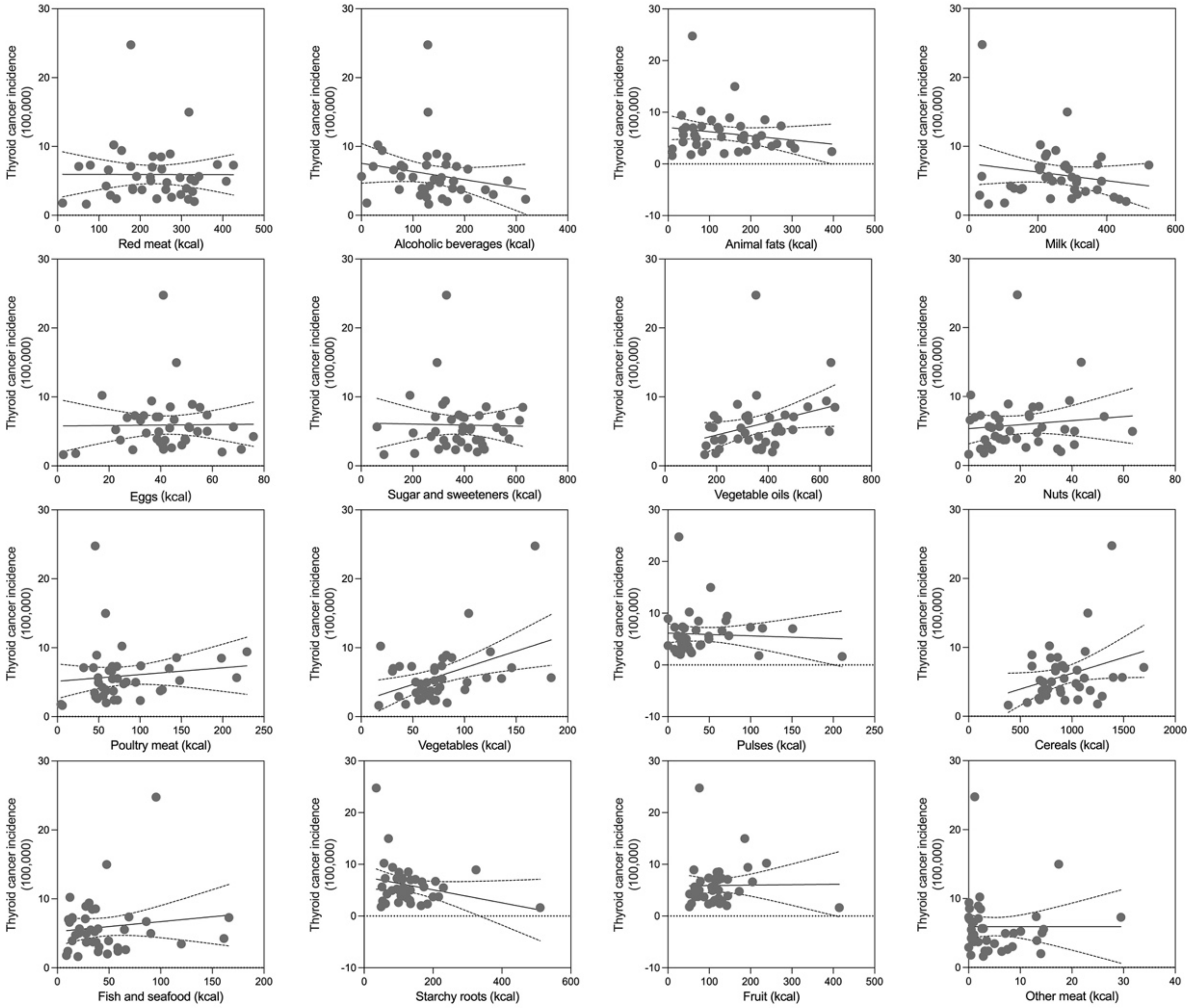


**Figure S26.** Regression analysis of association between 16 dietary factors and thyroid cancer incidence.

**Table S25.** Association between dietary factors and thyroid cancer incidence.

| **Food Type (n=40)** | **Correlation Coefficient** | | | | **Linear Regression** |
|---|---|---|---|---|---|
| | **CC** | **p-value** | **Partial CC** | **p-value** | **R squared** |
| Red meat | -0.003 | 0.984 | 0.002 | 0.989 | 0.0000 |
| Alcoholic beverages | -0.204 | 0.208 | -0.208 | 0.204 | 0.0415 |
| Animal fats | -0.183 | 0.258 | -0.202 | 0.217 | 0.0336 |
| Milk | -0.178 | 0.273 | -0.210 | 0.200 | 0.0315 |
| Eggs | 0.013 | 0.938 | 0.017 | 0.920 | 0.0002 |
| Sugar and sweeteners | -0.023 | 0.887 | -0.021 | 0.897 | 0.0005 |
| **Vegetable oils*** | **0.321** | **0.044** | **0.369** | **0.021** | **0.1029** |
| Nuts | 0.109 | 0.502 | 0.134 | 0.416 | 0.0120 |
| Poultry meat | 0.120 | 0.459 | 0.134 | 0.415 | 0.0145 |
| **Vegetables*** | **0.440** | **0.004** | **0.447** | **0.004** | **0.1938** |
| Pulses | -0.055 | 0.735 | -0.062 | 0.706 | 0.0030 |
| Cereals | 0.308 | 0.053 | 0.310 | 0.054 | 0.0947 |
| Fish and seafood | 0.131 | 0.421 | 0.148 | 0.368 | 0.0171 |
| Starchy roots | -0.260 | 0.105 | -0.275 | 0.090 | 0.0676 |
| Fruit | 0.013 | 0.938 | 0.012 | 0.944 | 0.0002 |
| Other meat | 0.001 | 0.997 | 0.004 | 0.980 | 0.0000 |

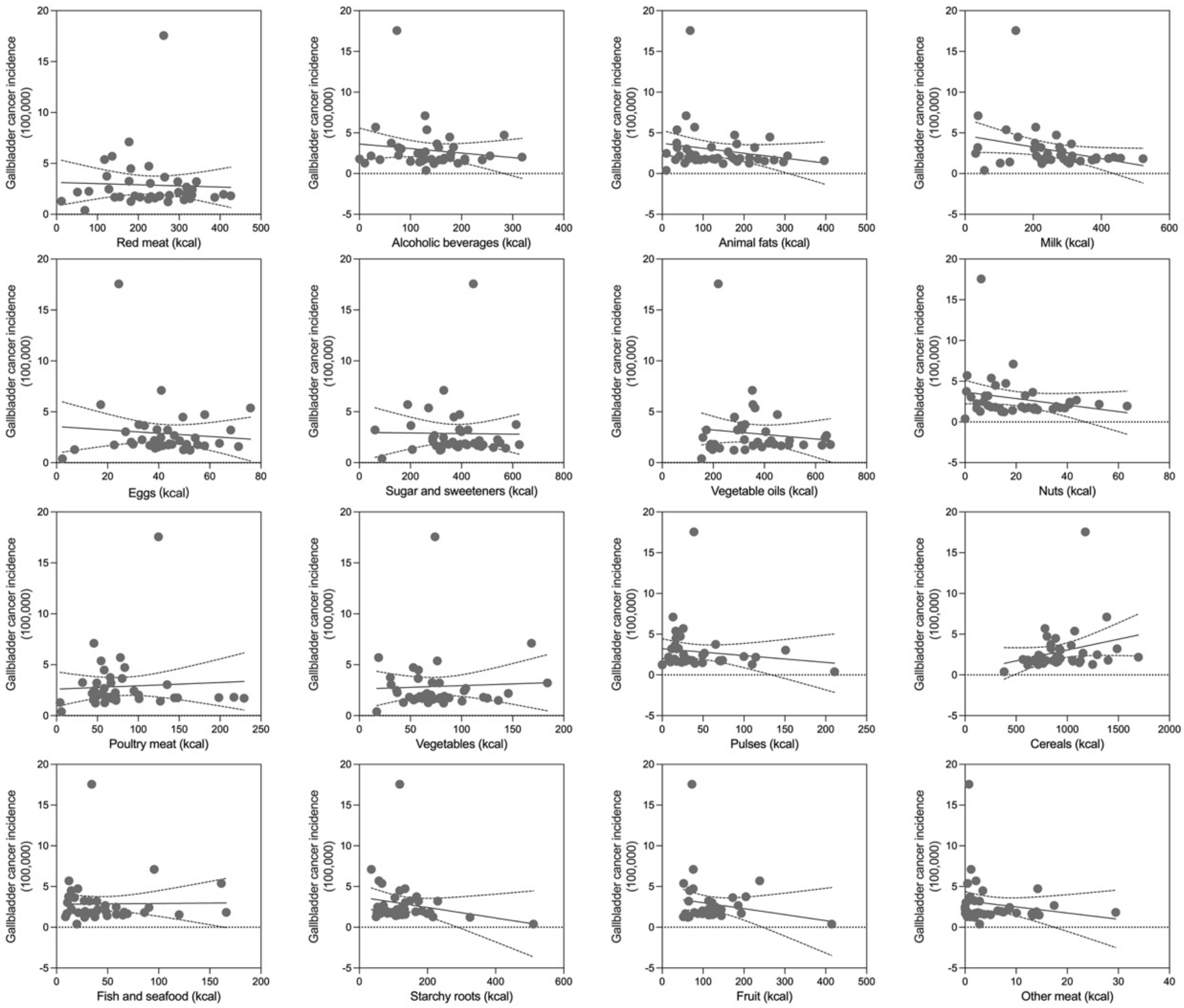


**Figure S27.** Regression analysis of association between 16 dietary factors and gallbladder cancer incidence.

**Table S26.** Association between dietary factors and gallbladder cancer incidence.

| Food Type (n=40) | Correlation Coefficient | | | | Linear Regression |
|---|---|---|---|---|---|
| | CC | p-value | Partial CC | p-value | R squared |
| Red meat | -0.041 | 0.804 | 0.069 | 0.676 | 0.0016 |
| Alcoholic beverages | -0.142 | 0.382 | -0.105 | 0.526 | 0.0202 |
| Animal fats | -0.206 | 0.202 | -0.147 | 0.371 | 0.0425 |
| Milk | -0.302 | 0.058 | -0.256 | 0.116 | 0.0914 |
| Eggs | -0.091 | 0.577 | -0.037 | 0.823 | 0.0083 |
| Sugar and sweeteners | -0.013 | 0.934 | 0.063 | 0.703 | 0.0002 |
| Vegetable oils | -0.113 | 0.487 | -0.039 | 0.811 | 0.0128 |
| Nuts | -0.222 | 0.168 | -0.161 | 0.328 | 0.0494 |
| Poultry meat | 0.06 | 0.713 | 0.137 | 0.404 | 0.0036 |
| Vegetables | 0.046 | 0.779 | 0.074 | 0.654 | 0.0021 |
| Pulses | -0.133 | 0.413 | -0.207 | 0.205 | 0.0177 |
| Cereals | 0.265 | 0.098 | 0.245 | 0.133 | 0.0703 |
| Fish and seafood | 0.013 | 0.937 | 0.092 | 0.577 | 0.0002 |
| Starchy roots | -0.199 | 0.218 | -0.265 | 0.103 | 0.0397 |
| Fruit | -0.171 | 0.291 | -0.198 | 0.226 | 0.0293 |
| Other meat | -0.174 | 0.283 | -0.124 | 0.452 | 0.0303 |

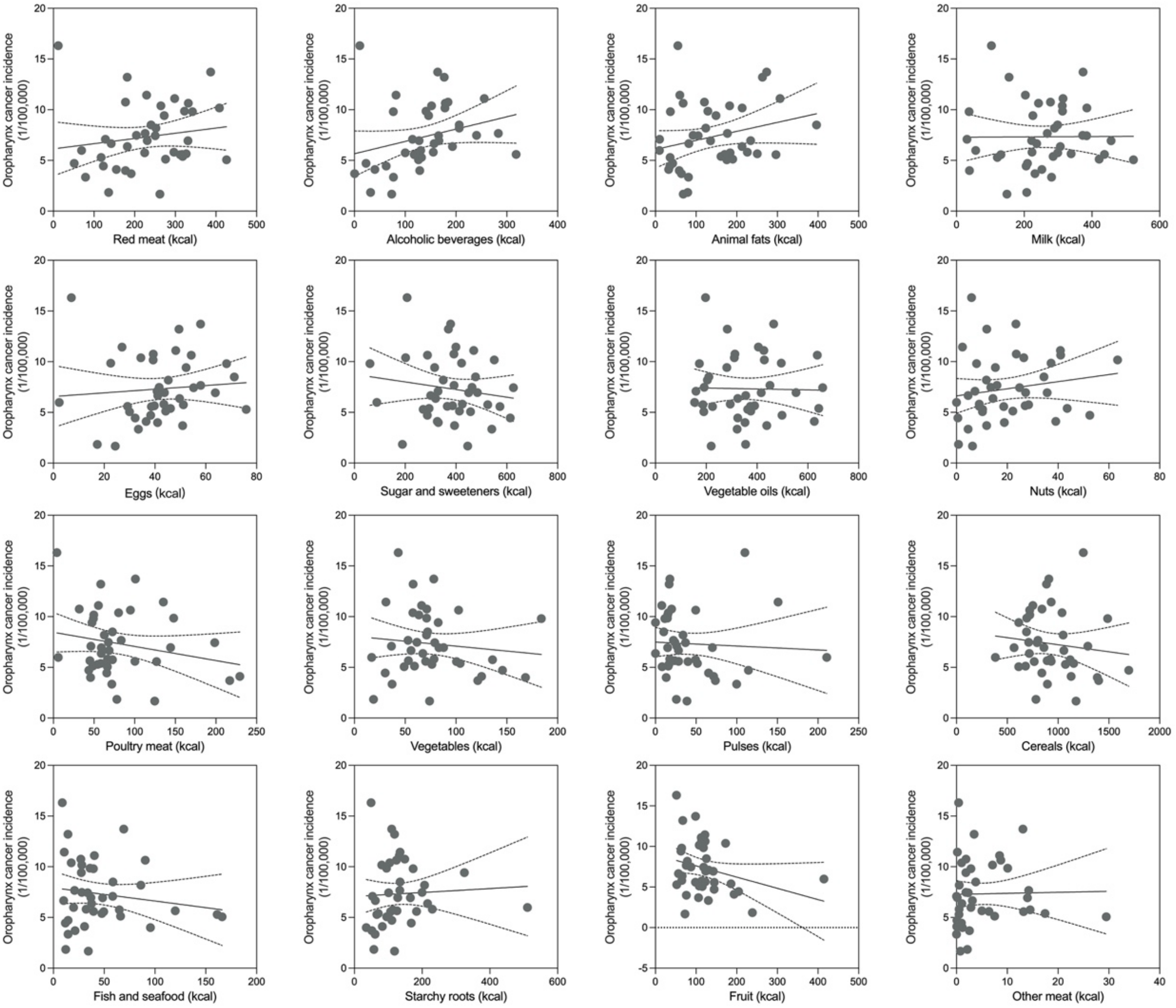


**Figure S28.** Regression analysis of association between 16 dietary factors and oropharynx cancer incidence.

**Table S27.** Association between dietary factors and oropharynx cancer incidence.

| Food Type (n=40) | Correlation Coefficient | | | | Linear Regression |
|---|---|---|---|---|---|
| | **CC** | **p-value** | **Partial CC** | **p-value** | **R squared** |
| Red meat | 0.156 | 0.335 | 0.244 | 0.134 | 0.0244 |
| Alcoholic beverages | 0.264 | 0.100 | 0.293 | 0.070 | 0.0698 |
| Animal fats | 0.255 | 0.112 | 0.328 | 0.042 | 0.0650 |
| Milk | 0.006 | 0.970 | 0.059 | 0.722 | 0.0000 |
| Eggs | 0.086 | 0.598 | 0.118 | 0.473 | 0.0074 |
| Sugar and sweeteners | -0.147 | 0.364 | -0.128 | 0.436 | 0.0217 |
| Vegetable oils | -0.020 | 0.903 | 0.017 | 0.918 | 0.0004 |
| Nuts | 0.165 | 0.308 | 0.241 | 0.140 | 0.0274 |
| Poultry meat | -0.216 | 0.180 | -0.204 | 0.212 | 0.0468 |
| Vegetables | -0.112 | 0.490 | -0.102 | 0.535 | 0.0126 |
| Pulses | -0.052 | 0.751 | -0.083 | 0.615 | 0.0027 |
| Cereals | -0.118 | 0.468 | -0.132 | 0.423 | 0.0139 |
| Fish and seafood | -0.152 | 0.350 | -0.133 | 0.419 | 0.0230 |
| Starchy roots | 0.051 | 0.755 | 0.030 | 0.855 | 0.0026 |
| Fruit | -0.275 | 0.086 | -0.288 | 0.075 | 0.0755 |
| Other meat | 0.020 | 0.902 | 0.050 | 0.761 | 0.0004 |

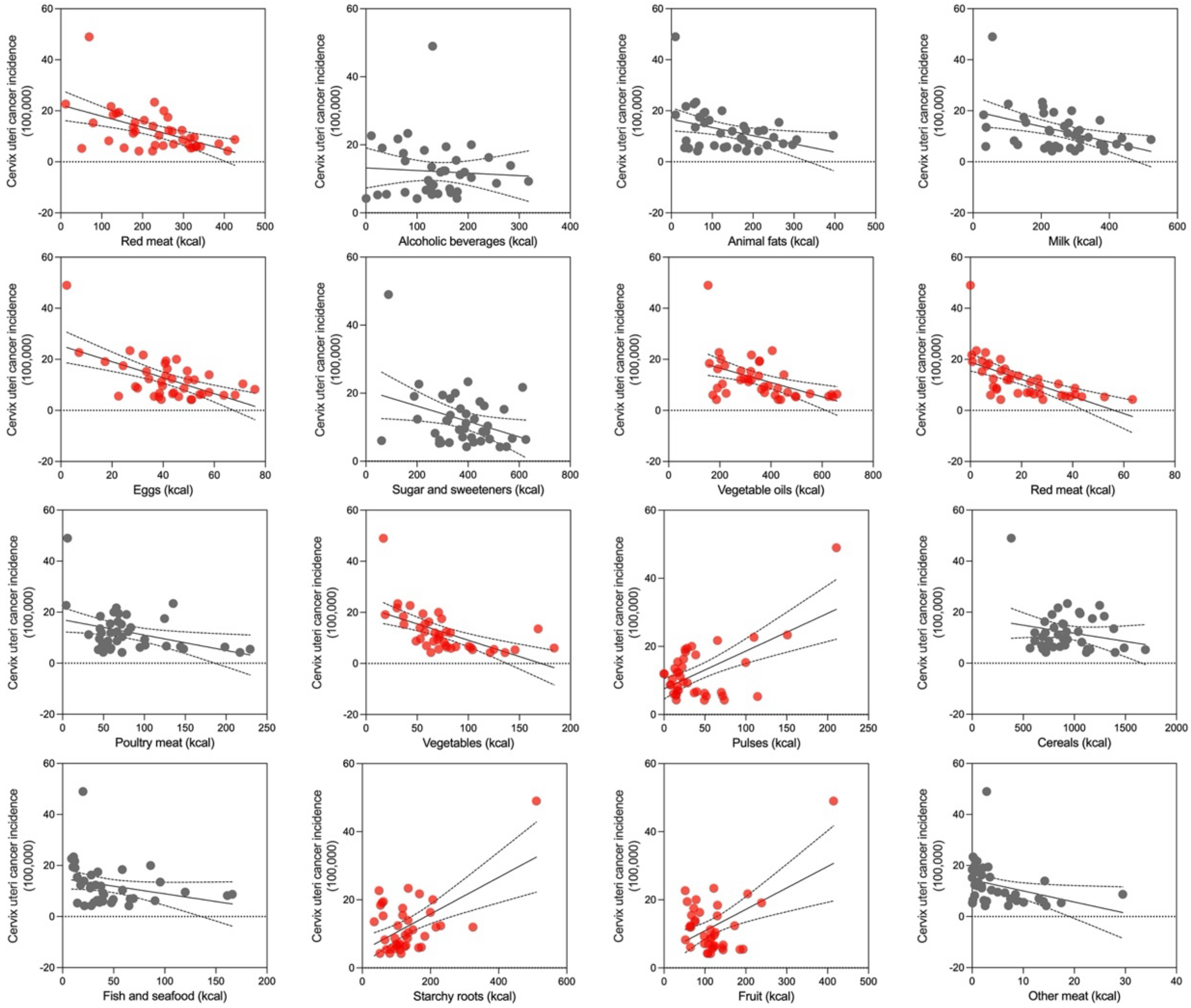


**Figure S29.** Regression analysis of association between 16 dietary factors and cervix uteri cancer incidence.

**Table S28.** Association between dietary factors and cervix uteri cancer incidence.

| **Food Type (n=40)** | **Correlation Coefficient** | | | | **Linear Regression** |
|---|---|---|---|---|---|
| | **CC** | **p-value** | **Partial CC** | **p-value** | **R squared** |
| Red meat | -0.523 | 0.001 | -0.255 | 0.117 | 0.2732 |
| Alcoholic beverages | -0.064 | 0.694 | 0.124 | 0.452 | 0.0041 |
| Animal fats | -0.364 | 0.021 | -0.107 | 0.515 | 0.1326 |
| Milk | -0.43 | 0.006 | -0.118 | 0.474 | 0.1853 |
| **Eggs*** | **-0.589** | **0.000** | **-0.514** | **0.001** | **0.3474** |
| Sugar and sweeteners | -0.349 | 0.027 | -0.123 | 0.454 | 0.1221 |
| Vegetable oils | -0.479 | 0.002 | -0.268 | 0.099 | 0.2296 |
| **Nuts*** | **-0.623** | **0.000** | **-0.445** | **0.005** | **0.3885** |
| Poultry meat | -0.368 | 0.019 | -0.175 | 0.287 | 0.1355 |
| **Vegetables*** | **-0.58** | **0.000** | **-0.624** | **0.000** | **0.3364** |
| **Pulses*** | **0.586** | **0.000** | **0.501** | **0.001** | **0.3437** |
| Cereals | -0.211 | 0.191 | -0.404 | 0.011 | 0.0446 |
| Fish and seafood | -0.275 | 0.086 | -0.020 | 0.905 | 0.0754 |
| **Starchy roots*** | **0.554** | **0.000** | **0.493** | **0.001** | **0.3069** |
| **Fruit*** | **0.492** | **0.001** | **0.525** | **0.001** | **0.2416** |
| Other meat | -0.336 | 0.034 | -0.152 | 0.356 | 0.1128 |

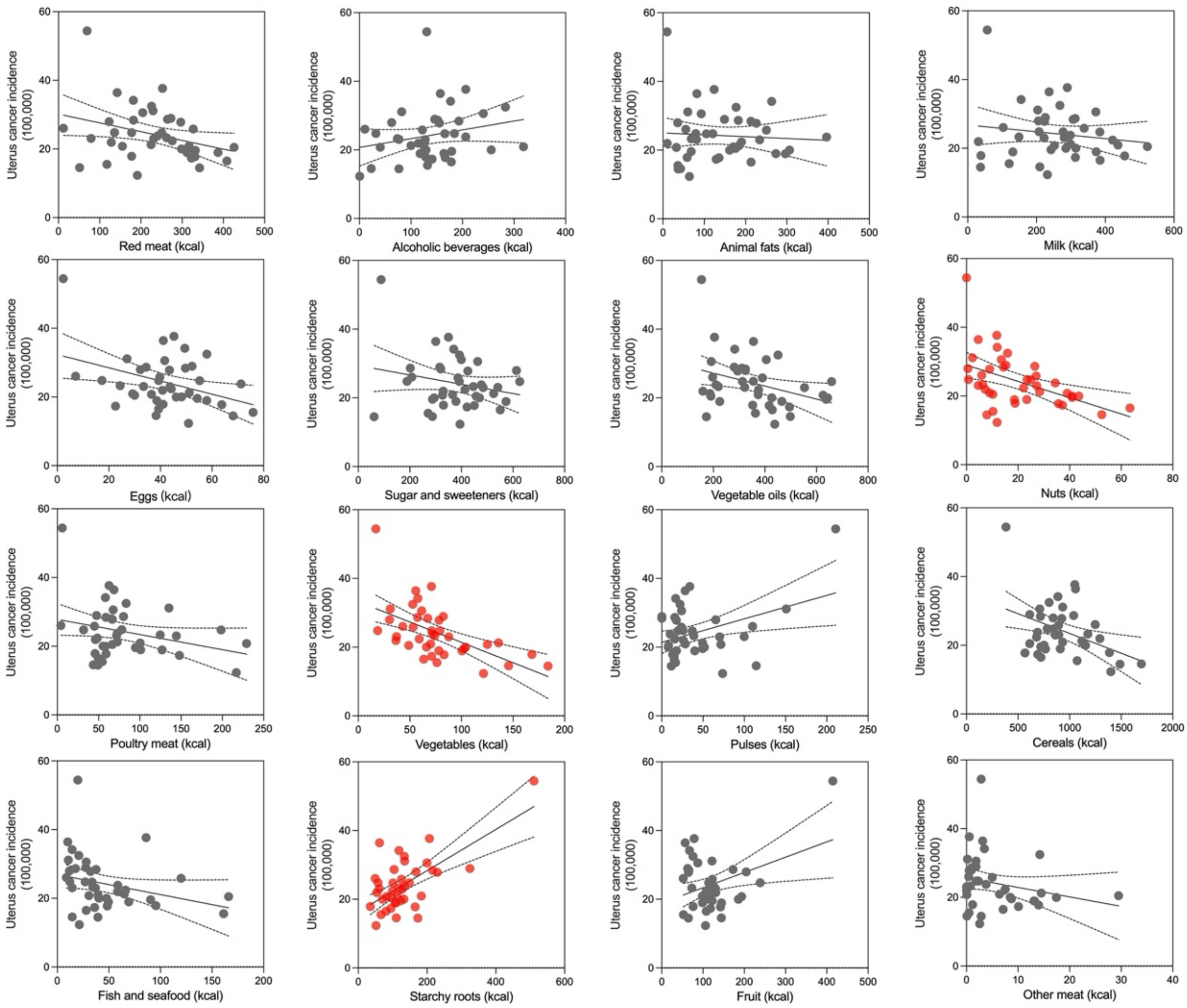


**Figure S30.** Regression analysis of association between 16 dietary factors and uterus cancer incidence.

**Table S29.** Association between dietary factors and uterus cancer incidence.

| **Food Type (n=40)** | **Correlation Coefficient** | | | | **Linear Regression** |
|---|---|---|---|---|---|
| | **CC** | **p-value** | **Partial CC** | **p-value** | **R squared** |
| Red meat | -0.325 | 0.041 | -0.065 | 0.696 | 0.1054 |
| Alcoholic beverages | 0.232 | 0.149 | 0.418 | 0.008 | 0.0539 |
| Animal fats | -0.065 | 0.692 | 0.206 | 0.209 | 0.0042 |
| Milk | -0.150 | 0.355 | 0.172 | 0.294 | 0.0225 |
| Eggs | -0.384 | 0.015 | -0.267 | 0.100 | 0.1472 |
| Sugar and sweeteners | -0.222 | 0.169 | -0.024 | 0.884 | 0.0492 |
| Vegetable oils | -0.342 | 0.031 | -0.148 | 0.368 | 0.1173 |
| Nuts | -0.467 | 0.002 | -0.287 | 0.077 | 0.2179 |
| Poultry meat | -0.285 | 0.074 | -0.121 | 0.462 | 0.0814 |
| **Vegetables*** | **-0.569** | **0.000** | **-0.570** | **0.000** | **0.3236** |
| Pulses | 0.384 | 0.015 | 0.261 | 0.108 | 0.1471 |
| **Cereals*** | **-0.407** | **0.009** | **-0.560** | **0.000** | **0.1653** |
| Fish and seafood | -0.282 | 0.078 | -0.101 | 0.539 | 0.0797 |
| **Starchy roots*** | **0.654** | **0.000** | **0.610** | **0.000** | **0.4282** |
| **Fruit*** | **0.368** | **0.019** | **0.348** | **0.030** | **0.1356** |
| Other meat | -0.226 | 0.161 | -0.063 | 0.701 | 0.0510 |

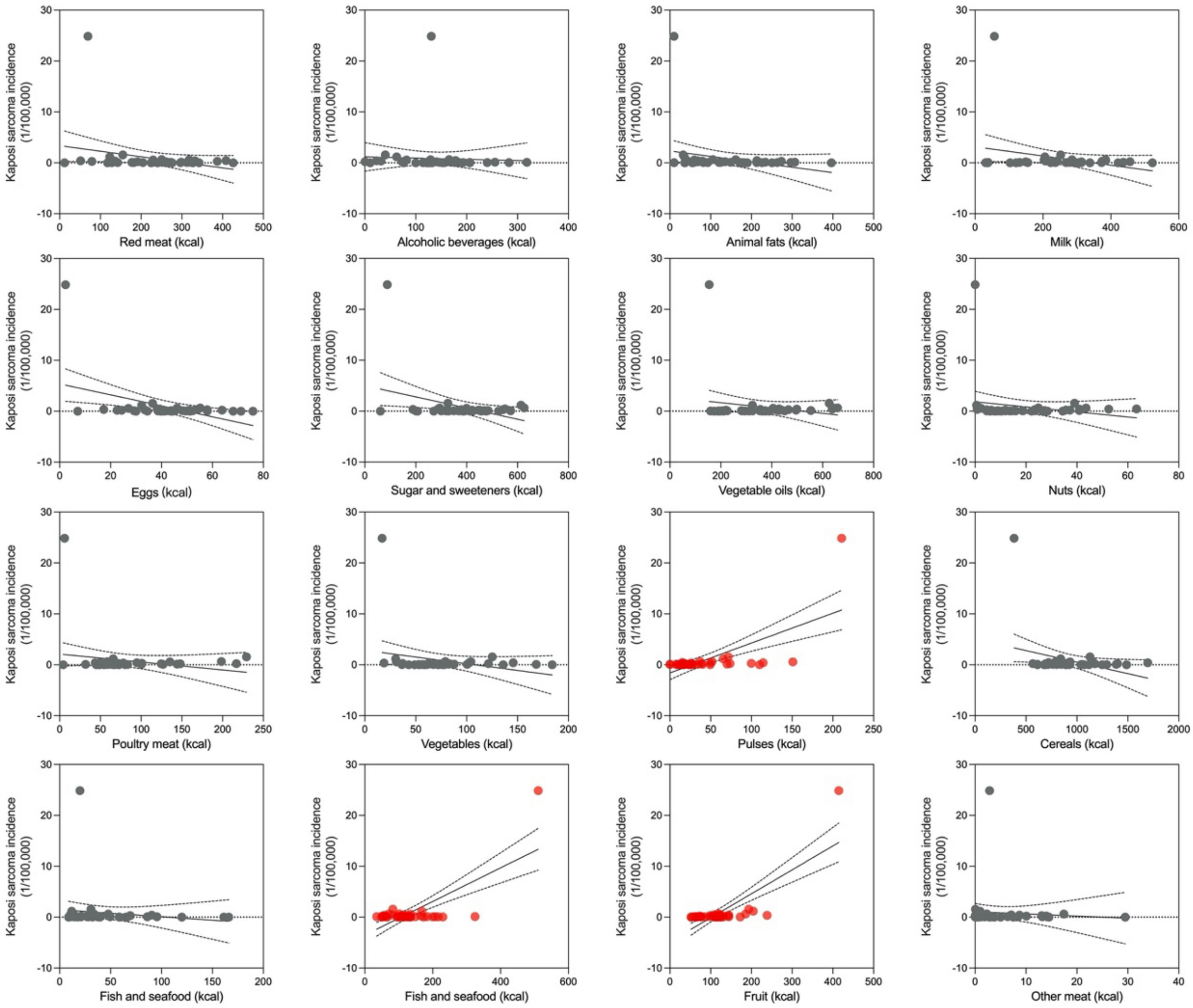


**Figure S31.** Regression analysis of association between 16 dietary factors and Kaposi sarcoma incidence.

**Table S30.** Association between dietary factors and Kaposi sarcoma incidence.

| **Food Type (n=40)** | **Correlation Coefficient** | | | | **Linear Regression** |
|---|---|---|---|---|---|
| | **CC** | **p-value** | **Partial CC** | **p-value** | **R squared** |
| Red meat | -0.276 | 0.084 | -0.160 | 0.330 | 0.0763 |
| Alcoholic beverages | -0.044 | 0.789 | 0.024 | 0.886 | 0.0019 |
| Animal fats | -0.253 | 0.115 | -0.155 | 0.346 | 0.0641 |
| Milk | -0.272 | 0.090 | -0.156 | 0.343 | 0.0738 |
| **Eggs*** | **-0.428** | **0.006** | **-0.374** | **0.019** | **0.1828** |
| Sugar and sweeteners | -0.357 | 0.024 | -0.282 | 0.082 | 0.1271 |
| Vegetable oils | -0.188 | 0.245 | -0.077 | 0.642 | 0.0354 |
| Nuts | -0.194 | 0.231 | -0.070 | 0.671 | 0.0376 |
| Poultry meat | -0.201 | 0.214 | -0.113 | 0.495 | 0.0404 |
| Vegetables | -0.252 | 0.117 | -0.221 | 0.176 | 0.0634 |
| **Pulses*** | **0.654** | **0.000** | **0.622** | **0.000** | **0.4277** |
| **Cereals*** | **-0.318** | **0.046** | **-0.378** | **0.018** | **0.1012** |
| Fish and seafood | -0.133 | 0.413 | -0.028 | 0.866 | 0.0177 |
| **Starchy roots*** | **0.716** | **0.000** | **0.692** | **0.000** | **0.5123** |
| **Fruit*** | **0.773** | **0.000** | **0.771** | **0.000** | **0.5980** |
| Other meat | -0.069 | 0.674 | 0.028 | 0.866 | 0.0047 |

### Multiple Linear Regression Analysis (Table S31-S81)

#### OCI

**Table S31.** Model summary of the multiple linear regression.

| R | $R^2$ | Adjusted $R^2$ | Std. Error of the Estimate | Durbin-Watson |
|---|---|---|---|---|
| 0.883 | 0.780 | 0.763 | 42.937 | 1.520 |

**Table S32.** ANOVA of the multiple linear regression.

| | Sum of Squares | df | MS | F | P |
|---|---|---|---|---|---|
| Regression | 837794.626 | 10 | 83779.463 | 45.444 | 0.000 |
| Residual | 235977.717 | 128 | 1843.576 | | |
| Total | 1073772.340 | 138 | | | |

**Table S33.** Multiple linear regression.

| Items | B | SE | Beta | t | P | Tolerance | VIF | N |
|---|---|---|---|---|---|---|---|---|
| Red meat | 0.250 | 0.059 | 0.304 | 4.232 | 0.000 | 0.333 | 3.002 | 139 |
| Alcoholic Beverages | 0.444 | 0.085 | 0.335 | 5.189 | 0.000 | 0.413 | 2.424 | 139 |
| Animal fats group | 0.075 | 0.083 | 0.065 | 0.903 | 0.368 | 0.332 | 3.016 | 139 |
| Milk | 0.115 | 0.050 | 0.165 | 2.322 | 0.022 | 0.339 | 2.951 | 139 |
| Eggs | 0.131 | 0.354 | 0.028 | 0.370 | 0.712 | 0.309 | 3.236 | 139 |
| Sugar and Sweeteners | 0.080 | 0.041 | 0.128 | 1.957 | 0.053 | 0.399 | 2.507 | 139 |
| GDP per capita | 0.001 | 0.001 | 0.067 | 0.932 | 0.353 | 0.333 | 2.999 | 139 |
| BMI>25 | 0.055 | 0.402 | 0.010 | 0.137 | 0.892 | 0.347 | 2.883 | 139 |
| Smoking prevalence | 0.358 | 0.562 | 0.035 | 0.638 | 0.525 | 0.582 | 1.717 | 139 |
| Total calories | -0.012 | 0.014 | -0.070 | -0.869 | 0.386 | 0.267 | 3.744 | 139 |

**Brain and CNS**

**Table S34.** Model summary of the multiple linear regression.

| R | $R^2$ | Adjusted $R^2$ | Std. Error of the Estimate | Durbin-Watson |
|---|---|---|---|---|
| 0.724 | 0.524 | 0.337 | 2.470 | 2.719 |

**Table S35.** ANOVA of the multiple linear regression.

| | Sum of Squares | df | MS | F | P |
|---|---|---|---|---|---|
| Regression | 187.849 | 11 | 17.077 | 2.800 | 0.014 |
| Residual | 170.792 | 28 | 6.100 | | |
| Total | 358.641 | 39 | | | |

**Table S36.** Multiple linear regression.

| Items (n=40) | B | SE | Beta | t | P | Tolerance | VIF |
|---|---|---|---|---|---|---|---|
| Alcoholic beverages | -0.011 | 0.010 | -0.258 | -1.101 | 0.280 | 0.309 | 3.237 |
| Animal fats | 0.016 | 0.006 | 0.493 | 2.68 | 0.012 | 0.503 | 1.990 |
| Red meat | 0.002 | 0.007 | 0.061 | 0.267 | 0.791 | 0.327 | 3.054 |
| Milk | 0.014 | 0.006 | 0.522 | 2.35 | 0.026 | 0.345 | 2.900 |
| Nuts | -0.021 | 0.036 | -0.108 | -0.59 | 0.560 | 0.503 | 1.990 |
| Pulses | 0.005 | 0.015 | 0.078 | 0.371 | 0.713 | 0.383 | 2.611 |
| Cereals | 0.000 | 0.003 | 0.033 | 0.112 | 0.911 | 0.196 | 5.090 |
| GDP per capita | 0.000 | 0.000 | 0.217 | 0.93 | 0.361 | 0.313 | 3.195 |
| BMI>25 | -0.013 | 0.055 | -0.055 | -0.233 | 0.818 | 0.309 | 3.232 |
| Smoking prevalence | 0.073 | 0.096 | 0.151 | 0.765 | 0.450 | 0.435 | 2.298 |
| Total calories | -0.002 | 0.003 | -0.216 | -0.557 | 0.582 | 0.113 | 8.849 |

**Bladder**

**Table S37.** Model summary of the multiple linear regression.

| R | $R^2$ | Adjusted $R^2$ | Std. Error of the Estimate | Durbin-Watson |
|---|---|---|---|---|
| 0.791 | 0.626 | 0.480 | 3.314 | 1.708 |

**Table S38.** ANOVA of the multiple linear regression.

| | Sum of Squares | df | MS | F | P |
|---|---|---|---|---|---|
| Regression | 515.411 | 11 | 46.856 | 4.267 | 0.001 |
| Residual | 307.478 | 28 | 10.981 | | |
| Total | 822.888 | 39 | | | |

**Table S39.** Multiple linear regression.

| Items (n=40) | B | SE | Beta | t | P | Tolerance | VIF |
|---|---|---|---|---|---|---|---|
| Alcoholic beverages | 0.000 | 0.014 | -0.003 | -0.016 | 0.988 | 0.309 | 3.237 |
| Animal fats | 0.006 | 0.008 | 0.119 | 0.732 | 0.470 | 0.503 | 1.990 |
| Red meat | -0.005 | 0.009 | -0.101 | -0.502 | 0.620 | 0.327 | 3.054 |
| Milk | 0.004 | 0.008 | 0.102 | 0.519 | 0.608 | 0.345 | 2.900 |
| Nuts | 0.111 | 0.049 | 0.37 | 2.272 | 0.031 | 0.503 | 1.990 |
| Pulses | 0.002 | 0.020 | 0.016 | 0.086 | 0.932 | 0.383 | 2.611 |
| Cereals | -0.004 | 0.004 | -0.267 | -1.023 | 0.315 | 0.196 | 5.090 |
| GDP per capita | 0.000 | 0.000 | -0.12 | -0.582 | 0.565 | 0.313 | 3.195 |
| BMI>25 | -0.019 | 0.073 | -0.053 | -0.254 | 0.801 | 0.309 | 3.232 |
| Smoking prevalence | 0.140 | 0.128 | 0.191 | 1.089 | 0.286 | 0.435 | 2.298 |
| Total calories | 0.006 | 0.004 | 0.468 | 1.363 | 0.184 | 0.113 | 8.849 |

**Breast**

**Table S40.** Model summary of the multiple linear regression.

| R | $R^2$ | Adjusted $R^2$ | Std. Error of the Estimate | Durbin-Watson |
|---|---|---|---|---|
| 0.918 | 0.842 | 0.780 | 10.445 | 2.049 |

**Table S41.** ANOVA of the multiple linear regression.

| | Sum of Squares | df | MS | F | P |
|---|---|---|---|---|---|
| Regression | 16318.716 | 11 | 1483.520 | 13.597 | 0.000 |
| Residual | 3055.034 | 28 | 109.108 | | |
| Total | 19373.750 | 39 | | | |

**Table S42.** Multiple linear regression.

| Items (n=40) | B | SE | Beta | t | P | Tolerance | VIF |
|---|---|---|---|---|---|---|---|
| Alcoholic beverages | -0.042 | 0.043 | -0.134 | -0.992 | 0.33 | 0.309 | 3.237 |
| Animal fats | 0.052 | 0.025 | 0.218 | 2.059 | 0.049 | 0.503 | 1.990 |
| Red meat | 0.013 | 0.03 | 0.059 | 0.447 | 0.658 | 0.327 | 3.054 |
| Milk | 0.045 | 0.024 | 0.235 | 1.842 | 0.076 | 0.345 | 2.900 |
| Nuts | 0.184 | 0.154 | 0.126 | 1.193 | 0.243 | 0.503 | 1.99 |
| Pulses | -0.027 | 0.062 | -0.053 | -0.435 | 0.667 | 0.383 | 2.611 |
| Cereals | -0.026 | 0.014 | -0.318 | -1.879 | 0.071 | 0.196 | 5.090 |
| GDP per capita | 0.000 | 0.000 | 0.017 | 0.126 | 0.901 | 0.313 | 3.195 |
| BMI>25 | -0.303 | 0.231 | -0.177 | -1.313 | 0.200 | 0.309 | 3.232 |
| Smoking prevalence | -0.318 | 0.405 | -0.09 | -0.787 | 0.438 | 0.435 | 2.298 |
| Total calories | 0.035 | 0.013 | 0.597 | 2.673 | 0.012 | 0.113 | 8.849 |

#### Corpus uteri

**Table S43.** Model summary of the multiple linear regression.

| R | $R^2$ | Adjusted $R^2$ | Std. Error of the Estimate | Durbin-Watson |
|---|---|---|---|---|
| 0.790 | 0.624 | 0.477 | 3.388 | 1.887 |

**Table S44.** ANOVA of the multiple linear regression.

| | Sum of Squares | df | MS | F | P |
|---|---|---|---|---|---|
| Regression | 534.115 | 11 | 48.556 | 4.230 | 0.001 |
| Residual | 321.421 | 28 | 11.479 | | |
| Total | 855.536 | 39 | | | |

**Table S45.** Multiple linear regression.

| Items (n=40) | B | SE | Beta | t | P | Tolerance | VIF |
|---|---|---|---|---|---|---|---|
| Alcoholic beverages | 0.010 | 0.014 | 0.157 | 0.756 | 0.456 | 0.309 | 3.237 |
| Animal fats | 0.010 | 0.008 | 0.193 | 1.183 | 0.247 | 0.503 | 1.990 |
| Red meat | -0.007 | 0.010 | -0.157 | -0.773 | 0.446 | 0.327 | 3.054 |
| Milk | 0.002 | 0.008 | 0.044 | 0.224 | 0.825 | 0.345 | 2.900 |
| Nuts | 0.014 | 0.050 | 0.045 | 0.278 | 0.783 | 0.503 | 1.990 |
| Pulses | -0.018 | 0.020 | -0.171 | -0.915 | 0.368 | 0.383 | 2.611 |
| Cereals | -0.004 | 0.004 | -0.223 | -0.853 | 0.401 | 0.196 | 5.090 |
| GDP per capita | 0.000 | 0.000 | -0.214 | -1.032 | 0.311 | 0.313 | 3.195 |
| BMI>25 | 0.124 | 0.075 | 0.345 | 1.657 | 0.109 | 0.309 | 3.232 |
| Smoking prevalence | 0.123 | 0.131 | 0.165 | 0.94 | 0.355 | 0.435 | 2.298 |
| Total calories | 0.003 | 0.004 | 0.255 | 0.74 | 0.465 | 0.113 | 8.849 |

#### Hodgkin lymphoma

**Table S46.** Model summary of the multiple linear regression.

| R | $R^2$ | Adjusted $R^2$ | Std. Error of the Estimate | Durbin-Watson |
|---|---|---|---|---|
| 0.861 | 0.742 | 0.640 | 0.447 | 1.706 |

**Table S47.** ANOVA of the multiple linear regression.

| | Sum of Squares | df | MS | F | P |
|---|---|---|---|---|---|
| Regression | 16.089 | 11 | 1.463 | 7.310 | 0.000 |
| Residual | 5.602 | 28 | 0.200 | | |
| Total | 21.691 | 39 | | | |

**Table S48.** Multiple linear regression.

| Items (n=40) | B | SE | Beta | t | P | Tolerance | VIF |
|---|---|---|---|---|---|---|---|
| Alcoholic beverages | 0.000 | 0.002 | -0.036 | -0.206 | 0.838 | 0.309 | 3.237 |
| Animal fats | 0.000 | 0.001 | 0.051 | 0.38 | 0.707 | 0.503 | 1.990 |
| Red meat | 0.001 | 0.001 | 0.11 | 0.655 | 0.518 | 0.327 | 3.054 |
| Milk | 0.001 | 0.001 | 0.212 | 1.299 | 0.205 | 0.345 | 2.900 |
| Nuts | 0.010 | 0.007 | 0.209 | 1.545 | 0.133 | 0.503 | 1.99 |
| Pulses | 0.005 | 0.003 | 0.318 | 2.051 | 0.05 | 0.383 | 2.611 |
| Cereals | -0.001 | 0.001 | -0.23 | -1.06 | 0.298 | 0.196 | 5.090 |
| GDP per capita | 0.000 | 0.000 | -0.027 | -0.157 | 0.876 | 0.313 | 3.195 |
| BMI>25 | 0.024 | 0.01 | 0.418 | 2.422 | 0.022 | 0.309 | 3.232 |
| Smoking prevalence | 0.019 | 0.017 | 0.157 | 1.081 | 0.289 | 0.435 | 2.298 |
| Total calories | 0.000 | 0.001 | 0.098 | 0.343 | 0.734 | 0.113 | 8.849 |

**Kidney**

**Table S49.** Model summary of the multiple linear regression.

| R | $R^2$ | Adjusted $R^2$ | Std. Error of the Estimate | Durbin-Watson |
|---|---|---|---|---|
| 0.804 | 0.647 | 0.509 | 2.249 | 2.230 |

**Table S50.** ANOVA of the multiple linear regression.

| | Sum of Squares | df | MS | F | P |
|---|---|---|---|---|---|
| Regression | 259.670 | 11 | 23.606 | 4.668 | 0.000 |
| Residual | 141.588 | 28 | 5.057 | | |
| Total | 401.258 | 39 | | | |

**Table S51.** Multiple linear regression.

| Items (n=40) | B | SE | Beta | t | P | Tolerance | VIF |
|---|---|---|---|---|---|---|---|
| Alcoholic beverages | 0.014 | 0.009 | 0.315 | 1.561 | 0.130 | 0.309 | 3.237 |
| Animal fats | 0.001 | 0.005 | 0.035 | 0.224 | 0.824 | 0.503 | 1.990 |
| Red meat | 0.002 | 0.006 | 0.066 | 0.337 | 0.738 | 0.327 | 3.054 |
| Milk | 0.001 | 0.005 | 0.022 | 0.116 | 0.908 | 0.345 | 2.90 |
| Nuts | -0.021 | 0.033 | -0.100 | -0.629 | 0.535 | 0.503 | 1.990 |
| Pulses | -0.017 | 0.013 | -0.226 | -1.243 | 0.224 | 0.383 | 2.611 |
| Cereals | -0.002 | 0.003 | -0.162 | -0.640 | 0.527 | 0.196 | 5.090 |
| GDP per capita | 0.000 | 0.000 | -0.317 | -1.579 | 0.126 | 0.313 | 3.195 |
| BMI>25 | 0.085 | 0.050 | 0.346 | 1.716 | 0.097 | 0.309 | 3.232 |
| Smoking prevalence | 0.042 | 0.087 | 0.082 | 0.483 | 0.633 | 0.435 | 2.298 |
| Total calories | 0.002 | 0.003 | 0.277 | 0.829 | 0.414 | 0.113 | 8.849 |

**Larynx**

**Table S52.** Model summary of the multiple linear regression.

| R | $R^2$ | Adjusted $R^2$ | Std. Error of the Estimate | Durbin-Watson |
|---|---|---|---|---|
| 0.795 | 0.632 | 0.487 | 1.017 | 1.720 |

**Table S53.** ANOVA of the multiple linear regression.

| | Sum of Squares | df | MS | F | P |
|---|---|---|---|---|---|
| Regression | 49.702 | 11 | 4.518 | 4.372 | 0.001 |
| Residual | 28.938 | 28 | 1.034 | | |
| Total | 78.640 | 39 | | | |

**Table S54.** Multiple linear regression.

| Items (n=40) | B | SE | Beta | t | P | Tolerance | VIF |
|---|---|---|---|---|---|---|---|
| Alcoholic beverages | 0.005 | 0.004 | 0.242 | 1.173 | 0.251 | 0.309 | 3.237 |
| Animal fats | 0.001 | 0.002 | 0.033 | 0.207 | 0.838 | 0.503 | 1.99 |
| Red meat | -0.001 | 0.003 | -0.086 | -0.43 | 0.670 | 0.327 | 3.054 |
| Milk | 0.002 | 0.002 | 0.152 | 0.779 | 0.443 | 0.345 | 2.9 |
| Nuts | 0.031 | 0.015 | 0.334 | 2.065 | 0.048 | 0.503 | 1.99 |
| Pulses | 0.007 | 0.006 | 0.219 | 1.18 | 0.248 | 0.383 | 2.611 |
| Cereals | 0.000 | 0.001 | 0.007 | 0.028 | 0.978 | 0.196 | 5.09 |
| GDP per capita | 0.000 | 0.000 | -0.574 | -2.799 | 0.009 | 0.313 | 3.195 |
| BMI>25 | 0.027 | 0.022 | 0.248 | 1.202 | 0.239 | 0.309 | 3.232 |
| Smoking prevalence | 0.113 | 0.039 | 0.498 | 2.866 | 0.008 | 0.435 | 2.298 |
| Total calories | 0.000 | 0.001 | -0.032 | -0.093 | 0.927 | 0.113 | 8.849 |

**Leukemia**

**Table S55.** Model summary of the multiple linear regression.

| R | $R^2$ | Adjusted $R^2$ | Std. Error of the Estimate | Durbin-Watson |
|---|---|---|---|---|
| 0.888 | 0.789 | 0.707 | 0.963 | 1.596 |

**Table S56.** ANOVA of the multiple linear regression.

| | Sum of Squares | df | MS | F | P |
|---|---|---|---|---|---|
| Regression | 97.223 | 11 | 8.838 | 9.535 | 0.000 |
| Residual | 25.954 | 28 | 0.927 | | |
| Total | 123.176 | 39 | | | |

**Table S57.** Multiple linear regression.

| Items (n=40) | B | SE | Beta | t | P | Tolerance | VIF |
|---|---|---|---|---|---|---|---|
| Alcoholic beverages | 0.002 | 0.004 | 0.064 | 0.409 | 0.686 | 0.309 | 3.237 |
| Animal fats | 0.002 | 0.002 | 0.124 | 1.015 | 0.319 | 0.503 | 1.99 |
| Red meat | -0.001 | 0.003 | -0.075 | -0.495 | 0.624 | 0.327 | 3.054 |
| Milk | -0.003 | 0.002 | -0.176 | -1.193 | 0.243 | 0.345 | 2.900 |
| Nuts | 0.008 | 0.014 | 0.067 | 0.547 | 0.589 | 0.503 | 1.990 |
| Pulses | -0.015 | 0.006 | -0.362 | -2.582 | 0.015 | 0.383 | 2.611 |
| Cereals | -0.001 | 0.001 | -0.218 | -1.113 | 0.275 | 0.196 | 5.090 |
| GDP per capita | 0.000 | 0.000 | -0.047 | -0.300 | 0.766 | 0.313 | 3.195 |
| BMI>25 | 0.064 | 0.021 | 0.467 | 2.996 | 0.006 | 0.309 | 3.232 |
| Smoking prevalence | -0.080 | 0.037 | -0.283 | -2.153 | 0.040 | 0.435 | 2.298 |
| Total calories | 0.002 | 0.001 | 0.466 | 1.804 | 0.082 | 0.113 | 8.849 |

**Oropharynx**

**Table S58.** Model summary of the multiple linear regression.

| R | $R^2$ | Adjusted $R^2$ | Std. Error of the Estimate | Durbin-Watson |
|---|---|---|---|---|
| 0.587 | 0.345 | 0.088 | 3.094 | 1.702 |

**Table S59.** ANOVA of the multiple linear regression.

| | Sum of Squares | df | MS | F | P |
|---|---|---|---|---|---|
| Regression | 141.251 | 11 | 12.841 | 1.341 | 0.254 |
| Residual | 268.085 | 28 | 9.574 | | |
| Total | 409.336 | 39 | | | |

**Table S60.** Multiple linear regression.

| Items (n=40) | B | SE | Beta | t | P | Tolerance | VIF |
|---|---|---|---|---|---|---|---|
| Alcoholic beverages | 0.015 | 0.013 | 0.319 | 1.16 | 0.256 | 0.309 | 3.237 |
| Animal fats | 0.013 | 0.007 | 0.364 | 1.688 | 0.102 | 0.503 | 1.990 |
| Red meat | 0.012 | 0.009 | 0.374 | 1.399 | 0.173 | 0.327 | 3.054 |
| Milk | 0.004 | 0.007 | 0.154 | 0.592 | 0.559 | 0.345 | 2.9 |
| Nuts | 0.079 | 0.046 | 0.371 | 1.72 | 0.096 | 0.503 | 1.99 |
| Pulses | 0.024 | 0.018 | 0.323 | 1.308 | 0.202 | 0.383 | 2.611 |
| Cereals | 0.004 | 0.004 | 0.356 | 1.032 | 0.311 | 0.196 | 5.090 |
| GDP per capita | 0.000 | 0.000 | -0.197 | -0.722 | 0.476 | 0.313 | 3.195 |
| BMI>25 | -0.058 | 0.068 | -0.232 | -0.845 | 0.405 | 0.309 | 3.232 |
| Smoking prevalence | -0.003 | 0.120 | -0.006 | -0.026 | 0.98 | 0.435 | 2.298 |
| Total calories | -0.004 | 0.004 | -0.514 | -1.131 | 0.268 | 0.113 | 8.849 |

**Lung**

**Table S61.** Model summary of the multiple linear regression.

| R | $R^2$ | Adjusted $R^2$ | Std. Error of the Estimate | Durbin-Watson |
|---|---|---|---|---|
| 0.853 | 0.727 | 0.620 | 6.526 | 2.288 |

**Table S62.** ANOVA of the multiple linear regression.

| | Sum of Squares | df | MS | F | P |
|---|---|---|---|---|---|
| Regression | 3177.640 | 11 | 288.876 | 6.783 | 0.000 |
| Residual | 1192.420 | 28 | 42.586 | | |
| Total | 4370.060 | 39 | | | |

**Table S63.** Multiple linear regression.

| Items (n=40) | B | SE | Beta | t | P | Tolerance | VIF |
|---|---|---|---|---|---|---|---|
| Alcoholic beverages | 0.022 | 0.027 | 0.147 | 0.825 | 0.416 | 0.309 | 3.237 |
| Animal fats | 0.019 | 0.016 | 0.164 | 1.176 | 0.249 | 0.503 | 1.990 |
| Red meat | -0.021 | 0.018 | -0.197 | -1.141 | 0.264 | 0.327 | 3.054 |
| Milk | 0.004 | 0.015 | 0.04 | 0.236 | 0.815 | 0.345 | 2.900 |
| Nuts | 0.118 | 0.096 | 0.17 | 1.221 | 0.232 | 0.503 | 1.99 |
| Pulses | -0.088 | 0.039 | -0.361 | -2.262 | 0.032 | 0.383 | 2.611 |
| Cereals | -0.009 | 0.009 | -0.232 | -1.042 | 0.306 | 0.196 | 5.090 |
| GDP per capita | 0.000 | 0.000 | -0.378 | -2.144 | 0.041 | 0.313 | 3.195 |
| BMI>25 | -0.263 | 0.144 | -0.324 | -1.823 | 0.079 | 0.309 | 3.232 |
| Smoking prevalence | 0.499 | 0.253 | 0.295 | 1.973 | 0.058 | 0.435 | 2.298 |
| Total calories | 0.021 | 0.008 | 0.741 | 2.523 | 0.018 | 0.113 | 8.849 |

**Skin melanoma**

**Table S64.** Model summary of the multiple linear regression.

| R | $R^2$ | Adjusted $R^2$ | Std. Error of the Estimate | Durbin-Watson |
|---|---|---|---|---|
| 0.790 | 0.625 | 0.477 | 5.774 | 1.646 |

**Table S65.** ANOVA of the multiple linear regression.

| | Sum of Squares | df | MS | F | P |
|---|---|---|---|---|---|
| Regression | 1553.292 | 11 | 141.208 | 4.235 | 0.001 |
| Residual | 933.585 | 28 | 33.342 | | |
| Total | 2486.877 | 39 | | | |

**Table S66.** Multiple linear regression.

| Items (n=40) | B | SE | Beta | t | P | Tolerance | VIF |
|---|---|---|---|---|---|---|---|
| Alcoholic beverages | 0.009 | 0.024 | 0.079 | 0.381 | 0.706 | 0.309 | 3.237 |
| Animal fats | 0.016 | 0.014 | 0.189 | 1.158 | 0.257 | 0.503 | 1.990 |
| Red meat | 0.026 | 0.016 | 0.319 | 1.574 | 0.127 | 0.327 | 3.054 |
| Milk | -0.017 | 0.013 | -0.251 | -1.271 | 0.214 | 0.345 | 2.900 |
| Nuts | 0.166 | 0.085 | 0.317 | 1.943 | 0.062 | 0.503 | 1.990 |
| Pulses | -0.002 | 0.034 | -0.012 | -0.062 | 0.951 | 0.383 | 2.611 |
| Cereals | -0.004 | 0.008 | -0.142 | -0.545 | 0.590 | 0.196 | 5.090 |
| GDP per capita | 0.000 | 0.000 | 0.295 | 1.423 | 0.166 | 0.313 | 3.195 |
| BMI>25 | 0.186 | 0.128 | 0.304 | 1.458 | 0.156 | 0.309 | 3.232 |
| Smoking prevalence | -0.352 | 0.224 | -0.276 | -1.573 | 0.127 | 0.435 | 2.298 |
| Total calories | -0.005 | 0.007 | -0.246 | -0.715 | 0.480 | 0.113 | 8.849 |

**Multiple melanoma**

**Table S67.** Model summary of the multiple linear regression.

| R | $R^2$ | Adjusted $R^2$ | Std. Error of the Estimate | Durbin-Watson |
|---|---|---|---|---|
| 0.856 | 0.732 | 0.627 | 0.648 | 1.745 |

**Table S68.** ANOVA of the multiple linear regression.

| | Sum of Squares | df | MS | F | P |
|---|---|---|---|---|---|
| Regression | 32.124 | 11 | 2.920 | 6.958 | 0.000 |
| Residual | 11.751 | 28 | 0.420 | | |
| Total | 43.875 | 39 | | | |

**Table S69.** Multiple linear regression.

| Items (n=40) | B | SE | Beta | t | P | Tolerance | VIF |
|---|---|---|---|---|---|---|---|
| Alcoholic beverages | -0.001 | 0.003 | -0.045 | -0.257 | 0.799 | 0.309 | 3.237 |
| Animal fats | 0.001 | 0.002 | 0.076 | 0.548 | 0.588 | 0.503 | 1.990 |
| Red meat | 0.003 | 0.002 | 0.302 | 1.768 | 0.088 | 0.327 | 3.054 |
| Milk | 0.000 | 0.002 | -0.049 | -0.296 | 0.769 | 0.345 | 2.900 |
| Nuts | 0.006 | 0.010 | 0.085 | 0.619 | 0.541 | 0.503 | 1.990 |
| Pulses | 0.002 | 0.004 | 0.095 | 0.598 | 0.554 | 0.383 | 2.611 |
| Cereals | -0.001 | 0.001 | -0.292 | -1.323 | 0.197 | 0.196 | 5.090 |
| GDP per capita | 0.000 | 0.000 | 0.039 | 0.221 | 0.827 | 0.313 | 3.195 |
| BMI>25 | 0.009 | 0.014 | 0.117 | 0.663 | 0.513 | 0.309 | 3.232 |
| Smoking prevalence | -0.030 | 0.025 | -0.174 | -1.177 | 0.249 | 0.435 | 2.298 |
| Total calories | 0.001 | 0.001 | 0.474 | 1.630 | 0.114 | 0.113 | 8.849 |

**Ovary**

**Table S70.** Model summary of the multiple linear regression.

| R | $R^2$ | Adjusted $R^2$ | Std. Error of the Estimate | Durbin-Watson |
|---|---|---|---|---|
| 0.835 | 0.698 | 0.579 | 1.611 | 2.378 |

**Table S71.** ANOVA of the multiple linear regression.

| | Sum of Squares | df | MS | F | P |
|---|---|---|---|---|---|
| Regression | 167.649 | 11 | 15.241 | 5.875 | 0.000 |
| Residual | 72.641 | 28 | 2.594 | | |
| Total | 240.290 | 39 | | | |

**Table S72.** Multiple linear regression.

| Items (n=40) | B | SE | Beta | t | P | Tolerance | VIF |
|---|---|---|---|---|---|---|---|
| Alcoholic beverages | 0.017 | 0.007 | 0.486 | 2.598 | 0.015 | 0.309 | 3.237 |
| Animal fats | 0.009 | 0.004 | 0.353 | 2.411 | 0.023 | 0.503 | 1.990 |
| Red meat | -0.005 | 0.005 | -0.188 | -1.037 | 0.309 | 0.327 | 3.054 |
| Milk | 0.005 | 0.004 | 0.236 | 1.332 | 0.193 | 0.345 | 2.900 |
| Nuts | -0.011 | 0.024 | -0.066 | -0.453 | 0.654 | 0.503 | 1.990 |
| Pulses | 0.000 | 0.010 | 0.004 | 0.024 | 0.981 | 0.383 | 2.611 |
| Cereals | 0.001 | 0.002 | 0.059 | 0.25 | 0.805 | 0.196 | 5.090 |
| GDP per capita | 0.000 | 0.000 | -0.149 | -0.800 | 0.430 | 0.313 | 3.195 |
| BMI>25 | 0.059 | 0.036 | 0.310 | 1.659 | 0.108 | 0.309 | 3.232 |
| Smoking prevalence | 0.064 | 0.062 | 0.162 | 1.031 | 0.311 | 0.435 | 2.298 |
| Total calories | -0.001 | 0.002 | -0.214 | -0.694 | 0.494 | 0.113 | 8.849 |

**Pancreas**

**Table S73.** Model summary of the multiple linear regression.

| R | $R^2$ | Adjusted $R^2$ | Std. Error of the Estimate | Durbin-Watson |
|---|---|---|---|---|
| 0.838 | 0.701 | 0.584 | 1.277 | 1.710 |

**Table S74.** ANOVA of the multiple linear regression.

| | Sum of Squares | df | MS | F | P |
|---|---|---|---|---|---|
| Regression | 107.319 | 11 | 9.756 | 5.981 | 0.000 |
| Residual | 45.670 | 28 | 1.631 | | |
| Total | 152.990 | 39 | | | |

**Table S75.** Multiple linear regression.

| Items (n=40) | B | SE | Beta | t | P | Tolerance | VIF |
|---|---|---|---|---|---|---|---|
| Alcoholic beverages | 0.009 | 0.005 | 0.307 | 1.650 | 0.110 | 0.309 | 3.237 |
| Animal fats | 0.002 | 0.003 | 0.107 | 0.734 | 0.469 | 0.503 | 1.990 |
| Red meat | -0.006 | 0.004 | -0.310 | -1.715 | 0.097 | 0.327 | 3.054 |
| Milk | 0.000 | 0.003 | 0.001 | 0.008 | 0.994 | 0.345 | 2.900 |
| Nuts | 0.005 | 0.019 | 0.035 | 0.242 | 0.810 | 0.503 | 1.990 |
| Pulses | -0.016 | 0.008 | -0.354 | -2.119 | 0.043 | 0.383 | 2.611 |
| Cereals | -0.002 | 0.002 | -0.218 | -0.938 | 0.356 | 0.196 | 5.090 |
| GDP per capita | 0.000 | 0.000 | -0.110 | -0.597 | 0.555 | 0.313 | 3.195 |
| BMI>25 | 0.005 | 0.028 | 0.034 | 0.182 | 0.857 | 0.309 | 3.232 |
| Smoking prevalence | 0.069 | 0.049 | 0.217 | 1.387 | 0.176 | 0.435 | 2.298 |
| Total calories | 0.002 | 0.002 | 0.473 | 1.541 | 0.135 | 0.113 | 8.849 |

#### Prostate

**Table S76.** Model summary of the multiple linear regression.

| R | $R^2$ | Adjusted $R^2$ | Std. Error of the Estimate | Durbin-Watson |
|---|---|---|---|---|
| 0.857 | 0.734 | 0.630 | 19.053 | 2.439 |

**Table S77.** ANOVA of the multiple linear regression.

| | Sum of Squares | df | MS | F | P |
|---|---|---|---|---|---|
| Regression | 28054.673 | 11 | 2550.425 | 7.026 | 0.000 |
| Residual | 10164.263 | 28 | 363.009 | | |
| Total | 38218.936 | 39 | | | |

**Table S78.** Multiple linear regression.

| Items (n=40) | B | SE | Beta | t | P | Tolerance | VIF |
|---|---|---|---|---|---|---|---|
| Alcoholic beverages | 0.047 | 0.078 | 0.107 | 0.608 | 0.548 | 0.309 | 3.237 |
| Animal fats | 0.017 | 0.046 | 0.051 | 0.368 | 0.716 | 0.503 | 1.990 |
| Red meat | 0.113 | 0.054 | 0.356 | 2.092 | 0.046 | 0.327 | 3.054 |
| Milk | 0.035 | 0.044 | 0.129 | 0.779 | 0.443 | 0.345 | 2.9 |
| Nuts | -0.248 | 0.281 | -0.121 | -0.882 | 0.385 | 0.503 | 1.99 |
| Pulses | 0.158 | 0.113 | 0.22 | 1.400 | 0.172 | 0.383 | 2.611 |
| Cereals | -0.027 | 0.025 | -0.236 | -1.072 | 0.293 | 0.196 | 5.090 |
| GDP per capita | 0.000 | 0.000 | -0.020 | -0.113 | 0.911 | 0.313 | 3.195 |
| BMI>25 | 0.303 | 0.421 | 0.126 | 0.719 | 0.478 | 0.309 | 3.232 |
| Smoking prevalence | 0.000 | 0.738 | -0.326 | -2.206 | 0.036 | 0.435 | 2.298 |
| Total calories | 0.032 | 0.024 | 0.393 | 1.354 | 0.186 | 0.113 | 8.849 |

**Testis**

**Table S79.** Model summary of the multiple linear regression.

| R | $R^2$ | Adjusted $R^2$ | Std. Error of the Estimate | Durbin-Watson |
|---|---|---|---|---|
| 0.781 | 0.610 | 0.457 | 2.157 | 1.607 |

**Table S80.** ANOVA of the multiple linear regression.

| | Sum of Squares | df | MS | F | P |
|---|---|---|---|---|---|
| Regression | 203.658 | 11 | 18.514 | 3.980 | 0.002 |
| Residual | 130.265 | 28 | 4.652 | | |
| Total | 333.923 | 39 | | | |

**Table S81.** Multiple linear regression.

| Items (n=40) | B | SE | Beta | t | P | Tolerance | VIF |
|---|---|---|---|---|---|---|---|
| Alcoholic beverages | 0.008 | 0.009 | 0.188 | 0.886 | 0.383 | 0.309 | 3.237 |
| Animal fats | 0.013 | 0.005 | 0.405 | 2.433 | 0.022 | 0.503 | 1.990 |
| Red meat | 0.008 | 0.006 | 0.284 | 1.376 | 0.180 | 0.327 | 3.054 |
| Milk | 0.001 | 0.005 | 0.046 | 0.228 | 0.822 | 0.345 | 2.90 |
| Nuts | 0.052 | 0.032 | 0.269 | 1.618 | 0.117 | 0.503 | 1.99 |
| Pulses | 0.006 | 0.013 | 0.093 | 0.487 | 0.630 | 0.383 | 2.611 |
| Cereals | 0.004 | 0.003 | 0.334 | 1.254 | 0.220 | 0.196 | 5.090 |
| GDP per capita | 0.000 | 0.000 | 0.369 | 1.751 | 0.091 | 0.313 | 3.195 |
| BMI>25 | 0.101 | 0.048 | 0.448 | 2.109 | 0.044 | 0.309 | 3.232 |
| Smoking prevalence | -0.037 | 0.084 | -0.08 | -0.449 | 0.657 | 0.435 | 2.298 |
| Total calories | -0.005 | 0.003 | -0.709 | -2.018 | 0.053 | 0.113 | 8.849 |
